# Pleiotropic heritability quantifies the shared genetic variance of common diseases

**DOI:** 10.1101/2025.06.10.25329261

**Authors:** Yujie Zhao, Benjamin Strober, Kangcheng Hou, Gaspard Kerner, John Danesh, Steven Gazal, Wei Cheng, Michael Inouye, Alkes L. Price, Xilin Jiang

## Abstract

Common diseases are highly pleiotropic, but the overall contribution of pleiotropy to a target disease’s architecture is unknown, as most studies focus on genetic correlations with each auxiliary disease in turn. Here we propose a new method, pleiotropic heritability with bias correction (PHBC), to estimate pleiotropic heritability (*h*^2^_*pleio*_), defined as the liability-scale genetic variance of a target disease that is shared with a specific set of auxiliary diseases. We estimate *h*^2^_*pleio*_ from GWAS summary statistics by estimating the proportion of variance explained from an estimated genetic correlation matrix and employing a Monte-Carlo bias correction procedure to account for sampling noise in genetic correlation estimates. Simulations showed that PHBC produces approximately unbiased estimates of pleiotropic heritability. The average ratio of pleiotropic heritability vs. total SNP-heritability (*h*^2^_*pleio*_/*h*^2^) across 15 diseases from the UK Biobank (spanning 7 disease categories) was 27% (s.e. 2%). Pleiotropic heritability was broadly distributed across disease categories, with *h*^2^_*pleio*_/*h*^2^ decreasing only slightly when removing all auxiliary diseases in the target disease category (avg = 24% (s.e. 2%)) and only moderately when further removing one other (most informative) category whose removal had the greatest impact (avg = 18% (s.e. 1%)). Average *h*^2^_*pleio*_/*h*^2^ increased to 33% (s.e. 2%) when adding 17 auxiliary quantitative traits in UK Biobank, and 49% (s.e. 4%) when further adding 30 auxiliary diseases from large GWAS meta-analyses—with several diseases dominated by pleiotropic heritability, including depression (74%, s.e. 7%) and type 2 diabetes (59%, s.e. 4%). On average, *h*^2^_*pleio*_/*h*^2^ was 1.51x (s.e. 0.14) larger than the proportion of liability-scale total phenotypic variance explained by the same set of auxiliary diseases, implying higher pleiotropy for genetic effects than the effects of non-genetic exposures. In conclusion, we have uncovered pervasive sharing of genetic aetiologies, with roughly half of common disease heritability being pleiotropic with diseases from a broad range of disease categories, which strongly motivates the importance of multi-disease approaches to risk prediction and therapeutic development.

## Introduction

Common diseases and complex traits are highly pleiotropic and extensive efforts have been made to understand the shared components of disease risk^1–6^. Previous studies have quantified and characterized the shared genetic variance between pairs of diseases^7–14^, identified latent genetic components that are shared across many diseases^15–23^, and leveraged the sharing of genetic variant associations across diseases to increase association power^17,19,24–29^. However, to our knowledge, we lack a method to estimate the genetic variance of a given target disease that is shared with other diseases and traits.

To address this gap, we introduce pleiotropic heritability (*h*^2^_*pleio*_), which quantifies the genetic variance of a target disease that is shared with a specific set of auxiliary diseases and traits. We develop a new method, pleiotropic heritability with bias correction (PHBC), to estimate *h*^2^_*pleio*_ from GWAS summary statistics. PHBC employs a Monte-Carlo bias correction procedure to correct bias arising from sampling noise in genetic correlation estimates due to finite GWAS sample size. We validate PHBC using extensive simulations then estimate *h*^2^_*pleio*_ for 15 diseases in UK Biobank and, subsequently, for 30 diseases with GWAS summary statistics available from extensive meta-analyses. Finally, we extend PHBC to estimate the liability-scale total phenotypic variance of a target disease that is shared with a set of auxiliary diseases.

## Results

### Definition of pleiotropic heritability

We define pleiotropic heritability as the liability-scale genetic variance (from SNPs) of a target disease that is shared with a specific set of auxiliary diseases. We can view the genetic value of the target disease as the sum of a disease-specific genetic value that is not shared with the auxiliary diseases and a pleiotropic genetic value that is shared with auxiliary diseases (**Figure 1A**), with the variance of latter denoted the pleiotropic heritability (*h*^2^_*pleio*_). We use the ratio of pleiotropic heritability to total SNP-heritability (*h*^2^_*pleio*_/*h*^2^) to quantify the proportion of genetic variance (from SNPs) that is pleiotropic. We note that the definition of pleiotropic heritability is dependent on both the target disease and the selected set of auxiliary diseases, and reflects specific phenotype definitions in specific cohorts.

**Figure 1.**
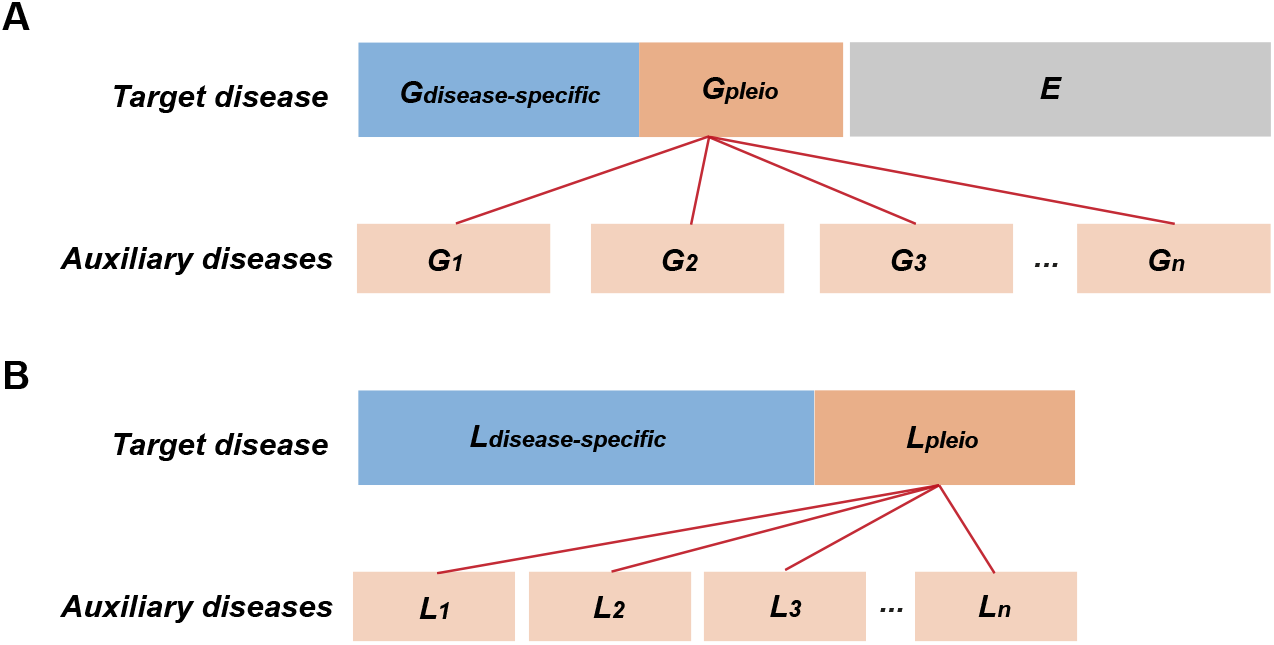
Definition of pleiotropic heritability and pleiotropic phenotypic variance. **(A)** The total phenotypic variance of the target disease consists of genetic variance (G) and environmental variance (E). The genetic variance of the target disease is partitioned into a disease-specific component and a pleiotropic component. The disease-specific component is not shared with the auxiliary diseases, and the pleiotropic component consists of a linear combination of the genetic values (*G*_1_, *G*_2_, …, *G*_*n*_) for auxiliary diseases 1 to *n*. Instances of G and E denote vectors of genetic and environmental values across individuals, and areas of rectangles denote variances. (B) The total phenotypic variance of the target disease is partitioned into a disease-specific component that is not shared with the auxiliary diseases, and a pleiotropic component that is shared with the auxiliary diseases. The pleiotropic phenotypic variance consists of a linear combination of total liabilities (*L*_1_, *L*_2_, …, *L*_*n*_) for auxiliary diseases 1 to *n*. Instances of L denote vectors of total liabilities across individuals, and areas of rectangles denote variances.

In detail, *h*^2^_*pleio*_ is the liability-scale variance explained by the weighted linear combination of auxiliary disease genetic liabilities that explains the maximum proportion of target disease heritability (in the entire population with infinite sample size):

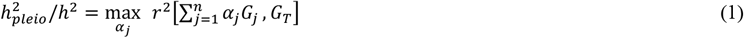

where *α*_*j*_ is the weight of the *j*^*th*^ auxiliary disease (*D*_*j*_), *r* is a correlation across individuals, *G*_*j*_ is the genetic liability of *D*_*j*_, and *G*_*T*_is the genetic liability of the target disease; *h*^2^_*pleio*_ is the corresponding *r*^2^ with the total liability *L*_*T*_instead of the genetic liability *G*_*T*_.

Alternatively, we can define *h*^2^_*pleio*_/*h*^2^ using the proportion of variance of causal SNP effect sizes on the target disease explained by the weighted linear combination of causal SNP effect sizes on the auxiliary diseases that explains the maximum proportion of variance:

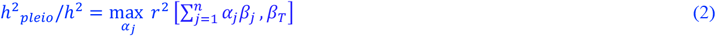

where *α*_*j*_ is the weight for the *j*^*th*^ auxiliary disease (*D*_*j*_), *r* is a correlation across SNPs, *β*_*j*_ is the vector of standardized causal SNP effect sizes on *D*_*j*_ (the number of s.d. increase in phenotype per 1 s.d. increase in genotype), and *β*_*T*_is the vector of standardized causal SNP effect sizes on the target disease. We note that under the assumption of no correlation between causal effect sizes of SNPs in LD with each other, the two definitions are equivalent (**Methods**).

We note that when an auxiliary disease *D*_*j*_ has zero genetic correlation with the target disease, its contribution to pleiotropic heritability of the target disease is zero. We emphasize that pleiotropic heritability is different from the heritability contributed by pleiotropic variants; if an auxiliary disease shares all causal variants with the target disease but with a genetic correlation of zero, its contribution to pleiotropic heritability of the target disease is zero, but 100% of the target disease heritability is contributed by pleiotropic variants. In this study, we restrict both pleiotropic heritability and total heritability to the heritability contributed by common SNPs (see below).

### Overview of estimation of pleiotropic heritability

We estimate *h*^2^_*pleio*_ from estimates of total SNP-heritability and genetic correlations between the target and auxiliary diseases, and employ a Monte-Carlo bias correction procedure to obtain unbiased estimates. We emphasize the need for a bias correction procedure: for example, if the target disease has true genetic correlation of 0 with each auxiliary disease (implying true pleiotropic heritability of 0), genetic correlation *estimates* will be nonzero due to sampling noise, implying a pleiotropic heritability *estimate* greater than 0 in the absence of a bias correction procedure. Monte-Carlo sampling is a widely used method that relies on random sampling to capture properties of a noise distribution^30^. In our application, Monte-Carlo sampling captures the noise distribution of the noisily estimated genetic correlations; when the sampled genetic correlations are used to estimate *h*^2^_*pleio*_/*h*^2^, the resulting distribution captures the bias in *h*^2^_*pleio*_/*h*^2^ arising from noisily estimated genetic correlations. Specifically, Monte-Carlo sampling is used to estimate the expected ratio between the true value (unbiased) vs. initial estimate (biased) of *h*^2^_*pleio*_/*h*^2^, so that the initial estimate can be scaled by this ratio to obtain an unbiased estimate. In this study, we apply cross-trait LD Score regression (LDSC)^8^ to summary association statistics and reference LD to estimate genetic correlations; we note that the cross-trait LDSC estimand is the genetic correlation of causal effect sizes of common SNPs (**Methods**). We obtain standard errors via genomic block-jackknife, as in cross-trait LDSC^8^. We have publicly released open-source software implementing our method (**Code Availability**).

In detail, it can be shown that Eq. 2 is equivalent to (see **Methods**):

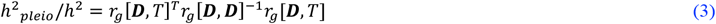

where *h*^2^ is the SNP-heritability of the target disease, *r*_*g*_[***D***, *T*] is the *n*×1 vector of genetic correlations between the target and auxiliary diseases, and *r*_*g*_[***D, D***] is the *n*×*n* genetic correlation matrix among the set of auxiliary diseases, where *n* is the number of auxiliary diseases (**Methods**). We initially estimate *h*^2^_*pleio*_/*h*^2^ by estimating the corresponding quantities in Eq. 3. However, the errors in estimates of *r*_*g*_[***D, D***] and *r*_*g*_[***D***, *T*] in finite GWAS sample size introduce an upward bias in estimates of *h*^2^_*pleio*_/*h*^2^, if *r*_*g*_[***D, D***]^− 1^ is positive definite (which is likely to be the case) (**Methods**).

To obtain an unbiased estimate, we first generate Monte-Carlo samples of estimation errors in *r*_*g*_[***D, D***] and *r*_*g*_[***D***, *T*] from the sampling covariance matrix computed from genomic block-jackknife^17^ (we use the same genomic block-jackknife for the target disease *T* and all auxiliary diseases ***D***; see **Code Availability**); these samples capture the joint distribution of estimation errors across elements in *r*_*g*_[***D, D***] and *r*_*g*_[***D***, *T*]. Then, we add the estimation errors to the point estimates of *r*_*g*_[***D, D***] and *r*_*g*_[***D***, *T*] to create Monte-Carlo samples of *h*^2^_*pleio*_/*h*^2^, which are larger than the initial estimate. We then use binary search to estimate a scaling coefficient *ξ*_*c*_ multiplying *r*_*g*_[***D***, *T*] so that the average *h*^2^_*pleio*_/*h*^2^ across Monte-Carlo samples matches the initial estimate of *h*^2^_*pleio*_/*h*^2^ (see **Methods**). Finally, we multiply the initial estimate of *h*^2^_*pleio*_/*h*^2^ (and its standard error) by 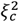 to obtain an unbiased estimate of *h*^2^_*pleio*_/*h*^2^.

In the presence of collinearity among auxiliary diseases and uncertainty in genetic correlation estimates, the estimate from Eq. 3 can be numerically unstable, which we define as s.e.(*r*_*g*_[***D***, *T*]^*T*^*r*_*g*_[***D, D***] ^−1^*r*_*g*_[***D***, *T*]) > 0.5 and/or *ξ*_*c*_ < 0.5 (see **Methods**). Whenever this occurs, we prune auxiliary traits as follows: for each pair of auxiliary traits with 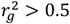, we remove the trait with lower *r*_*g*_ z-score with target trait; we repeat this procedure for 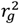 thresholds of 0.5, 0.4, 0.3, 0.2, and 0.1 until the estimate is numerically stable.

We extend our method to estimate the liability-scale total phenotypic variance of a target disease that is shared with a specific set of auxiliary diseases (*V*^2^_*pleio*_) by estimating the liability-scale phenotypic correlation between pairs of diseases using individual-level data^31^ (**Figure 1B**). (We note that the term pleiotropy generally refers to genetic effects, but we use the term pleiotropic phenotypic variance for consistency.) Further details are provided in the **Methods** section.

An overview of empirical data analyzed is as follows. We first analyzed 15 highly heritable target diseases (prevalence > 1%, heritability z-score > 6, and liability-scale heritability ranging from 0.061 to 0.21) from the UK Biobank^32^ (average N = 157K, spanning 7 PheCode disease categories^33^) (**Table 1**); the sample size reflects a restriction to unrelated individuals of British ancestry with diagnostic records from both primary care data and hospital inpatient data, which we imposed to mitigate the impact of missing diagnoses. In our UK Biobank analyses, we used the same 15 diseases as auxiliary diseases; we also performed analyses incorporating 17 quantitative traits^34,35^ from the UK Biobank as auxiliary traits (including blood biochemistry measurements, e.g. total cholesterol, blood pressure and HbA1c, and demographic traits, e.g. height, BMI and years of education; details in **Supplementary Table 2** and **Methods**). We subsequently incorporated 30 diseases with large meta-analyses available (average N = 483K) as target and/or auxiliary diseases. In all analyses, we restricted to auxiliary diseases with modest genetic correlation to the target disease (*r*_*g*_^2^ < 0.5). We have publicly released all summary association statistics analyzed in this study, along with estimates of pleiotropic heritability and related quantities (**Data Availability**). We note that while we elected to focus on target diseases, definition and estimation of pleiotropic heritability is also applicable to target quantitative traits. We recommend that users restrict to target and auxiliary traits that have heritability z-score > 6, and auxiliary traits that have *r*_*g*_^2^ < 0.5 with the target trait (**Methods**).

**Table 1.** Overview of 15 UK Biobank diseases.

| Disease | Prevalence | $h^2_{liab}$ (s.e.) | $h^2$ z-score | Category |
| --- | --- | --- | --- | --- |
| Hypothyroidism | 0.054 | 0.18 (0.024) | 7.7 | Endocrine |
| Type 2 diabetes | 0.11 | 0.19 (0.014) | 13 | Endocrine |
| Obesity | 0.20 | 0.14 (0.0087) | 16 | Endocrine |
| Depression | 0.17 | 0.087 (0.0082) | 11 | Mental disorders |
| Tobacco use disorder | 0.049 | 0.14 (0.017) | 7.8 | Mental disorders |
| Hypertension | 0.37 | 0.21 (0.011) | 19 | Circulatory system |
| Coronary atherosclerosis | 0.083 | 0.16 (0.015) | 11 | Circulatory system |
| Atrial fibrillation | 0.051 | 0.12 (0.019) | 6.2 | Circulatory system |
| Varicose veins | 0.069 | 0.12 (0.017) | 6.9 | Circulatory system |
| Respiratory diseases | 0.30 | 0.061 (0.0061) | 10 | Respiratory |
| GERD | 0.17 | 0.067 (0.0075) | 9.0 | Digestive |
| Inguinal hernia | 0.074 | 0.11 (0.015) | 7.1 | Digestive |
| Diverticulosis | 0.15 | 0.13 (0.010) | 13 | Digestive |
| Bunion | 0.048 | 0.15 (0.018) | 8.2 | Musculoskeletal |
| Radiculitis | 0.10 | 0.073 (0.010) | 7.0 | Symptoms |

### Simulations

We performed simulations to evaluate the unbiasedness of pleiotropic heritability estimates. We specified genetic architectures for 16 diseases using liability-scale heritabilities of 0.13 (median of 15 UK Biobank diseases; **Table 1**) and prevalences of 0.1 (median of 15 UK Biobank diseases; **Table 1**). True genetic correlations were set to 0.5 within disease categories and 0.1 between disease categories for the first 15 diseases (based on the 7 disease categories from **Table 1**) and 0.0 for the last disease (with each other disease), implying 5 different values of true *h*^2^_*pleio*_/*h*^2^ for each target disease (ranging from 0.00 to 0.38). The proportion of causal SNPs was set to 5%. Additional simulation settings were also evaluated. We simulated phenotypes from real UK Biobank genotypes using 1.1M HapMap3^36^ SNPs across 157,206 unrelated individuals. We applied PLINK2^37^ to compute summary association statistics and applied cross-trait LD score regression^8^ to summary association statistics for the 1.1M SNPs (MAF > 0.01) and reference LD from 1000 Genomes European individuals (9.3M SNPs) to estimate genetic correlations. We then estimated *h*^2^_*pleio*_ for each disease using the other 15 diseases as auxiliary diseases. Further details of the simulation framework are provided in the **Methods** section.

Results are reported in **Figure 2** and **Supplementary Table 4**. Before Monte-Carlo bias correction, *h*^2^_*pleio*_/*h*^2^ estimates suffered upward biases (between 0.024 (s.e. 0.0012) and 0.028 (s.e. 0.0034); average bias of 0.026 (s.e. 0.0011)), highlighting the need for a bias correction procedure. After Monte-Carlo bias correction, *h*^2^_*pleio*_/*h*^2^ estimates were approximately unbiased across the 16 simulated diseases with varying pleiotropic architectures. Most biases were small (between −0.007 (s.e. 0.003) and 0.0078 (s.e. 0.0006); average bias of −0.0020 (s.e. 0.0011)). We compared the estimated standard errors to empirical standard deviations, and determined that estimated standard errors were conservative, with the ratio of the average estimated squared jackknife standard error to the average empirical squared deviation equal to 1.63 (**Supplementary Figure 1** and **Supplementary Table 4**).

**Figure 2.**
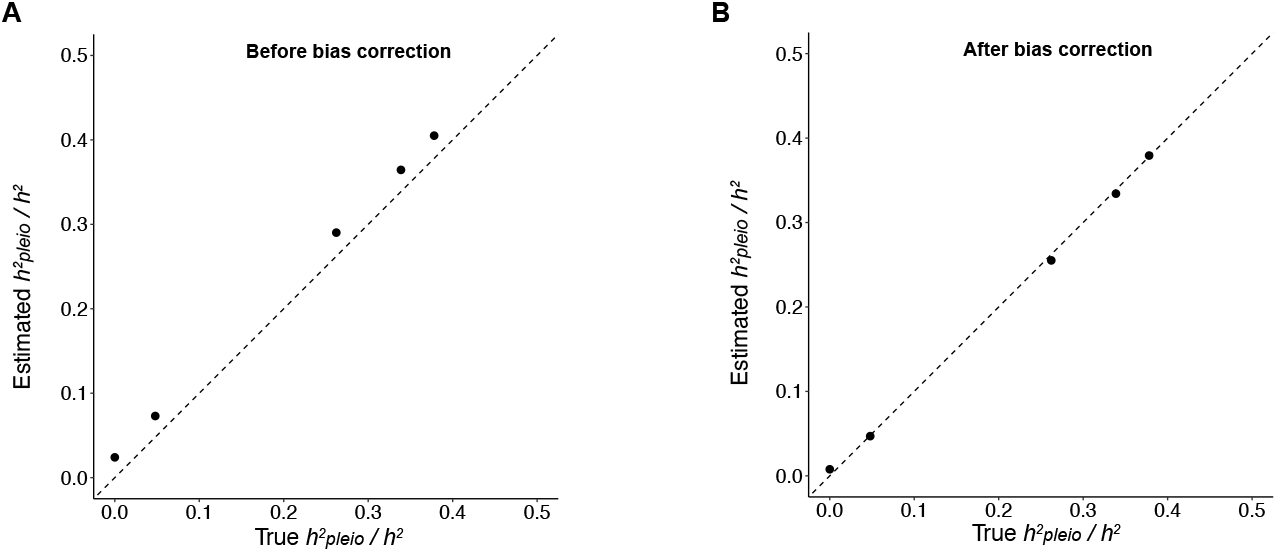
PHBC corrects upwards bias in simulations. (A) Estimates of *h*^2^_*pleio*_/*h*^2^ without the bias correction step suffer upward bias. (B) Estimates of *h*^2^_*pleio*_/*h*^2^ are approximately unbiased after the Monte-Carlo bias correction. Each dot represents the mean of all estimates for diseases with the same true value of *h*^2^_*pleio*_/*h*^2^ across 100 simulations. Error bars denote standard errors (but are generally smaller than dot size). Numerical results are reported in **Supplementary Table 4**.

We performed six secondary analyses. First, we performed simulations with the proportion of causal SNPs set to 1% (instead of 5%), and determined the results were similar to **Figure 2** (**Supplementary Figures 2-3** and **Supplementary Table 11**). Second, we simulated pleiotropic architectures with *r*_*g*_ within disease categories equal to 0.4, 0.3, or 0.2 (instead of 0.5) (and *r*_*g*_ between disease categories still equal to 0.1), and observed approximately unbiased results (**Supplementary Figures 4-5** and **Supplementary Tables 5-7**). Third, we performed simulations with liability-scale heritabilities equal to 0.25 or 0.06 (instead of 0.13). We observed approximately unbiased results with liability-scale heritability equal to 0.25 (**Supplementary Figures 6-7** and **Supplementary Tables 8**). With liability-scale heritability equal to 0.06, we observed modest downward bias for values of *h*^2^_*pleio*_/*h*^2^ above 25% and modest upward bias for values below 5%, with average bias of −0.017 (s.e. 0.0023) (**Supplementary Figures 8-9** and **Supplementary Tables 9)**. Fourth, we performed simulations with the prevalence set to 0.05 (instead of 0.1), and determined the results were similar to **Figure 2** (**Supplementary Figures 10-11** and **Supplementary Table 10**). Fifth, we estimated *h*^2^_*pleio*_/*h*^2^ without pruning highly correlated auxiliary diseases, and we observed that *h*^2^_*pleio*_/*h*^2^ estimates were approximately unbiased (**Supplementary Figures 12-13** and **Supplementary Table 12**). Across these five experiments, we determined that uncorrected estimates of *h*^2^_*pleio*_/*h*^2^ suffered upward bias (ranging from 1.1% (s.e. 0.12%) to 9.9% (s.e. 0.80%)), emphasizing the need for a bias correction procedure. Finally, we performed simulations to evaluate the genomic block-jackknife standard errors on the reduction in *h*^2^_*pleio*_/*h*^2^ in analyses with one auxiliary disease category removed, and determined that standard errors were conservative (**Supplementary Figure 14** and **Supplementary Table 13**).

### Application to 15 diseases from the UK Biobank

We estimated *h*^2^_*pleio*_/*h*^2^ for 15 highly heritable target diseases from the UK Biobank (heritability z-score > 6; average N=157K, liability-scale heritability ranging from 0.061 to 0.21, spanning 7 PheCode disease categories^33^) (**Table 1**). Genetic correlations across these 15 diseases are reported in **Figure 3A** and **Supplementary Table 14**, and corresponding liability-scale phenotypic correlations are reported in **Figure 3B** and **Supplementary Table 15**. We observed moderate genetic correlations within disease categories and between disease categories, and smaller phenotypic correlations.

**Figure 3.**
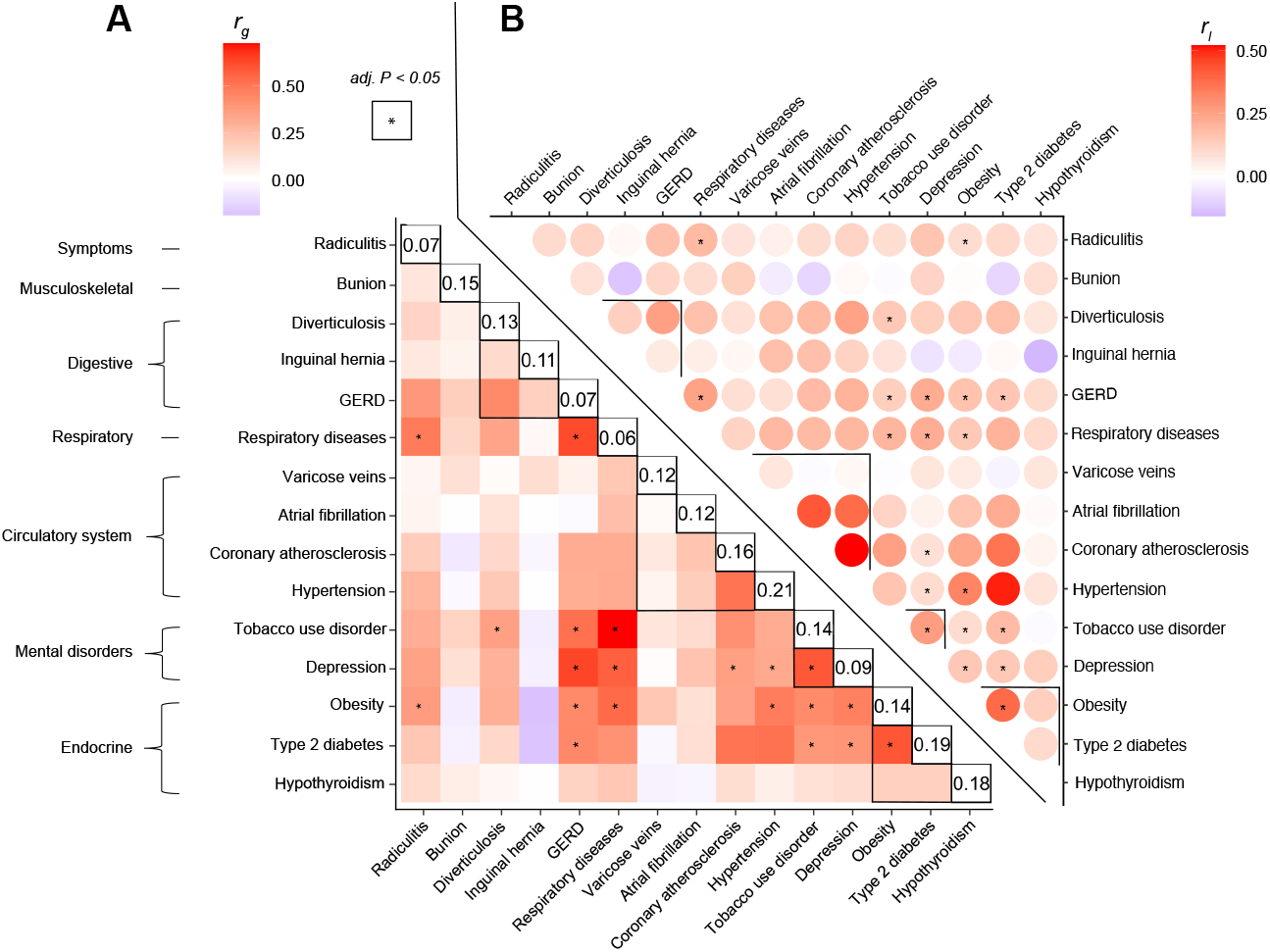
Estimates of genetic correlation and liability-scale phenotypic correlation. (A) Estimates of genetic correlation across 15 UK Biobank diseases. (B) Estimates of liability-scale phenotypic correlation across 15 UK Biobank diseases. Asterisks denote pairs of diseases for which the genetic correlation is significantly larger than the liability-scale phenotypic correlation with Bonferroni-corrected *P* < 4.76 × 10^−4^ (= 0.05/105); no pairs have liability-scale phenotypic correlation significantly larger than the genetic correlation. Black boundaries demarcate correlations between diseases within the same disease category. Numerical results are reported in **Supplementary Table 15**.

Estimates of *h*^2^_*pleio*_/*h*^2^ for three representative diseases (type 2 diabetes (T2D), depression (MDD), and hypertension (HTN)), and the average across 15 diseases, are reported in **Figure 4, Supplementary Figure 15** and **Supplementary Table 17**. We determined that 27% of SNP-heritability is pleiotropic on average (jackknife s.e.(avg) = 2%, s.d. = 19% across diseases); the Monte-Carlo bias correction procedure had a substantial impact on estimates of *h*^2^_*pleio*_/*h*^2^ (pre-correction average = 38%, jackknife s.e.(avg) = 4%, **Supplementary Figure 16, Supplementary Table 17**), consistent with simulations (**Figure 2**) and underscoring the importance of correcting upward bias. Several diseases had fairly high *h*^2^_*pleio*_/*h*^2^ estimates, including 44% (s.e. 7%) for MDD and 45% (s.e. 5%) for T2D.

**Figure 4.**
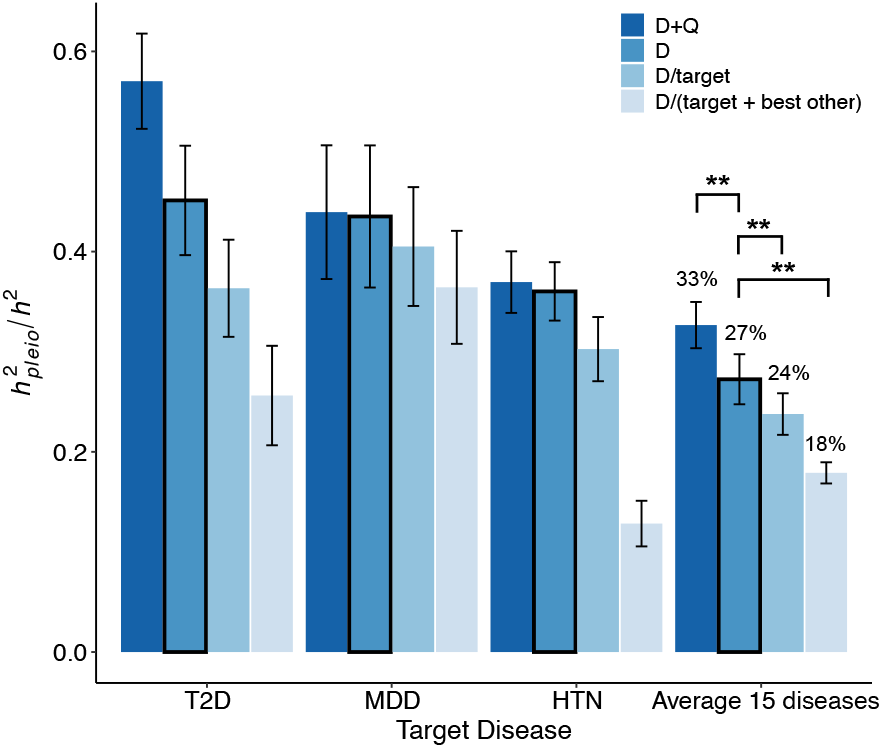
*h*^2^_*pleio*_/*h*^2^ estimates for three representative diseases and the average across 15 UK Biobank diseases. D+Q: *h*^2^_*pleio*_/*h*^2^ estimates with respect to 15 UK Biobank auxiliary diseases and 17 UK Biobank quantitative auxiliary traits. D: *h*^2^_*pleio*_/*h*^2^ estimates with respect to 15 UK Biobank auxiliary diseases. D\target: *h*^2^_*pleio*_/*h*^2^ estimates with respect to 15 UK Biobank auxiliary diseases excluding those from the same disease category as the target disease. D\(target+best other): *h*^2^_*pleio*_/*h*^2^ estimates with respect to 15 UK Biobank auxiliary diseases excluding those from the same disease category as the target disease and those from one other disease category whose removal had the greatest impact. Error bars denote jackknife standard errors. To assess statistical significance, we compare average estimates from D+Q, D\target and D\(target+best other) to estimates from D using inverse-variance weighting across diseases/traits. *: *P* < 0.05; **: *P* < 0.001. T2D: type 2 diabetes; MDD: depression; HTN: hypertension. Numerical results are reported in **Supplementary Tables 17 and 18**.

We performed three analyses in which we modified the set of auxiliary diseases (**Figure 4, Supplementary Figure 15** and **Supplementary Table 17**). First, we included 17 quantitative traits from the UK Biobank (**Supplementary Table 2**) as additional auxiliary traits, causing estimates of *h*^2^_*pleio*_/*h*^2^ to increase (avg = 33%, jackknife s.e.(avg) = 2%, s.d. = 21%). Second, we restricted to auxiliary diseases excluding the target disease category, causing estimates of *h*^2^_*pleio*_/*h*^2^ to decrease only slightly (avg = 24% (s.e. 2%); 41% (s.e. 6%) for MDD (removing mental) and 36% (s.e. 5%) for T2D (removing endocrine)). Third, we further removed one other disease category whose removal had the greatest impact, causing estimates of *h*^2^_*pleio*_/*h*^2^ to decrease only moderately (avg = 18% (s.e. 1%); 36% (s.e. 6%) for MDD (removing mental and digestive) and 26% (s.e.5%) for T2D (removing endocrine and circulatory system)). These results imply that pleiotropic heritability is broadly distributed across disease categories.

Estimates of *h*^2^_*pleio*_/*h*^2^ with respect to auxiliary diseases in each individual disease category are reported in **Figure 5, Supplementary Figure 17** and **Supplementary Table 19**. We determined that auxiliary diseases from a single disease category can often explain the majority of pleiotropic heritability. For example, MDD had 41% (s.e. 7%) and 31% (s.e. 2%) of SNP-heritability shared with digestive and respiratory disease categories, respectively (vs. 44% for all disease categories), consistent with the high pleiotropic overlap between brain-gut and brain-lung axes in previous studies^38,39^); T2D had 33% (s.e. 4%) and 25% (s.e. 4%) of SNP-heritability shared with circulatory and digestive disease categories, respectively (vs. 45% for all disease categories), consistent with the importance of T2D-related processes in the etiology of cardiometabolic diseases in previous studies^22^); and the 15 target diseases on average had 17% (s.e. 3%) and 16% (s.e. 4%) of SNP-heritability shared with endocrine and mental disorder categories, respectively (vs. 27% for all disease categories).

**Figure 5.**
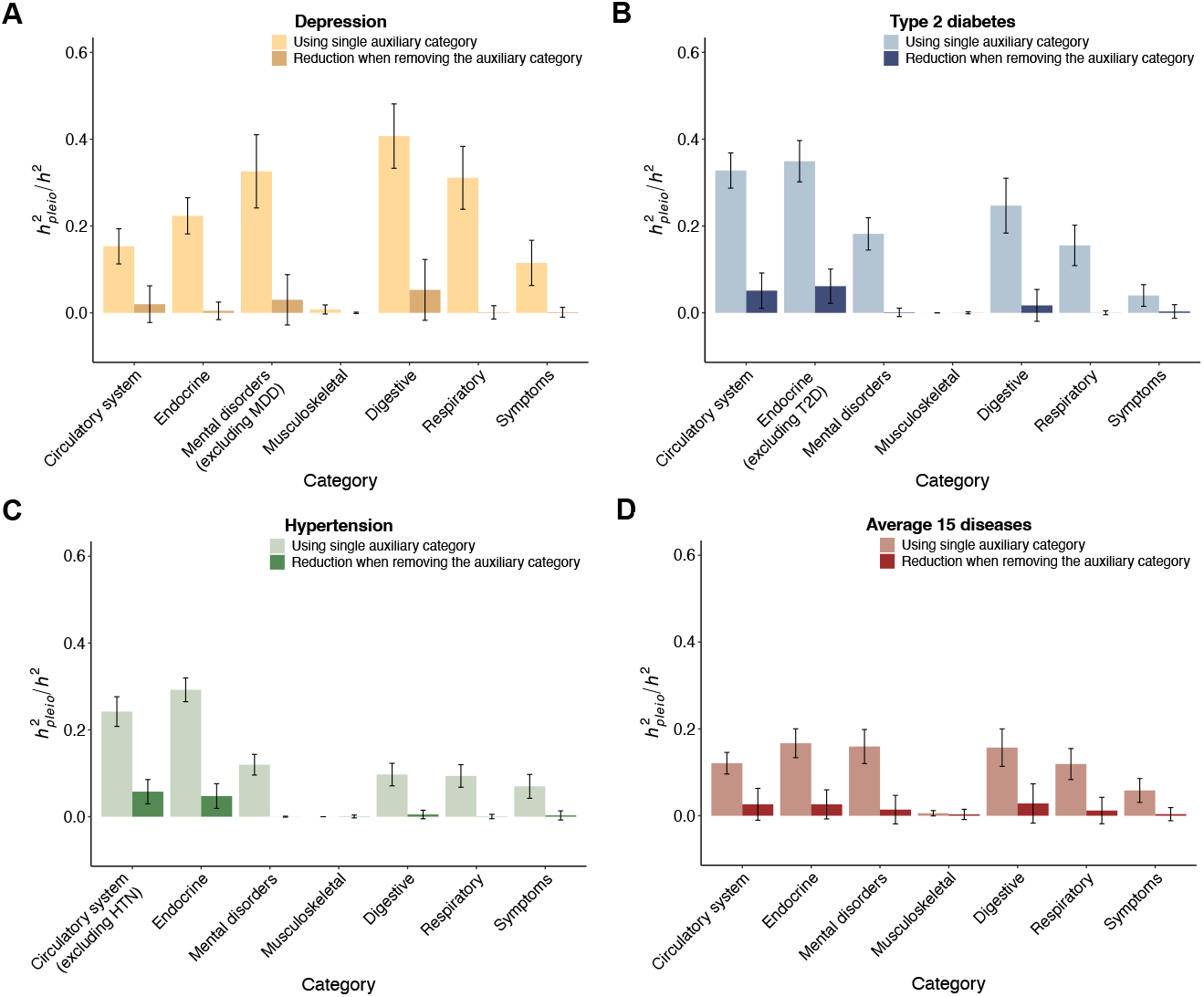
*h*^2^_*pleio*_/*h*^2^ is broadly shared across PheCode disease categories. (A-C) Comparison between *h*^2^_*pleio*_/*h*^2^ using a single auxiliary disease category (light bars) and the reduction in *h*^2^_*pleio*_/*h*^2^ when removing the auxiliary disease category (dark bars), for three representative UK Biobank diseases (depression, type 2 diabetes, hypertension) and the average across 15 UK Biobank diseases. Error bars denote jackknife standard errors. Numerical results are reported in **Supplementary Table 19**.

However, removing a single disease category from the set of auxiliary diseases generally reduced *h*^2^_*pleio*_/*h*^2^ much less than the *h*^2^_*pleio*_/*h*^2^ shared with that disease category (**Figure 5, Supplementary Figure 17** and **Supplementary Table 19**). For example, MDD *h*^2^_*pleio*_/*h*^2^ was reduced by only 5% (s.e. 7%) and 0.1% (s.e. 1.5%) when removing digestive and respiratory categories, respectively; T2D *h*^2^_*pleio*_/*h*^2^ was reduced by only 7% (s.e. 4%) and 2% (s.e. 3%) when removing circulatory and digestive disease categories, respectively; and average *h*^2^_*pleio*_/*h*^2^ across the 15 target diseases was reduced by only 3% (s.e. 3%) and 1% (s.e. 3%) when removing endocrine and mental disorder categories, respectively. These results imply that pleiotropic heritability is broadly shared across disease categories, such that removal of one disease category has a limited impact due to shared pleiotropic heritability with other disease categories.

To investigate the impact of educational attainment (EA) on pleiotropic heritability, we performed two analyses. First, we estimated *h*^2^_*pleio*_/*h*^2^ with respect to EA (years of education) as the only auxiliary trait. We determined that *h*^2^_*pleio*_/*h*^2^ with respect to EA is 9.9% (s.e. 0.8%) on average across 15 UK Biobank diseases (**Supplementary Figure 18** and **Supplementary Table 28**). Second, we assessed the impact of removing EA from the set of auxiliary traits by estimating the difference between (i) *h*^2^_*pleio*_/*h*^2^ with respect to 15 UK Biobank auxiliary diseases + 17 UK Biobank auxiliary quantitative traits vs. (ii) *h*^2^_*pleio*_/*h*^2^ with respect to 15 UK Biobank auxiliary diseases + 16 UK Biobank quantitative traits excluding EA. We determined that the differences were small, with an average reduction of 0.87% (s.e. 0.39%) (**Supplementary Figure 18** and **Supplementary Table 28**). This implies that the large contribution of EA to pleiotropic heritability is mostly captured by other auxiliary traits.

We performed five secondary analyses. First, we investigated whether there is a correlation between liability-scale heritability and *h*^2^_*pleio*_/*h*^2^ across the 15 UK Biobank diseases. We observed no significant correlation (correlation = −0.18 (*P* = 0.52); **Supplementary Figure 19**). Second, we estimated the average *h*^2^_*pleio*_/*h*^2^ across 15 UK Biobank diseases using different *r*_*g*_^2^ thresholds between target and auxiliary diseases (ranging from 0.1 to 0.8 in increments of 0.05) (**Supplementary Figure 20** and **Supplementary Table 29**). Average *h*^2^_*pleio*_/*h*^2^ increased at *r*_*g*_^2^ thresholds larger than 0.5, but we consider this increase to be not biologically meaningful because most instances of *r*_*g*_^2^ > 0.5 involved auxiliary traits that *are not biologically distinct* from the target trait, e.g. hypertension-diastolic blood pressure (*r*_*g*_^2^ = 0.72), hypertension-systolic blood pressure (*r*_*g*_^2^ = 0.72), type 2 diabetes-glucose (*r*_*g*_^2^ = 0.69), type 2 diabetes-HbA1C (*r*_*g*_^2^ = 0.79). In addition, average *h*^2^_*pleio*_/*h*^2^ decreased at *r*_*g*_^2^ thresholds smaller than 0.5, but all instances of 0.25 < *r*_*g*_^2^ < 0.5 involved auxiliary traits that *are biologically distinct* from the target trait: coronary atherosclerosis-hypertension (*r*_*g*_^2^ = 0.25), coronary atherosclerosis-type 2 diabetes (*r*_*g*_^2^ = 0.25), hypertension-type 2 diabetes (*r*_*g*_^2^ = 0.26), depression-tobacco use disorder (*r*_*g*_^2^ = 0.35), gastroesophageal reflux disease (GERD)-tobacco use disorder (*r*_*g*_^2^ = 0.27), obesity-type 2 diabetes (*r*_*g*_^2^ = 0.36), obesity-respiratory diseases (*r*_*g*_^2^ = 0.29), obesity-HbA1c (*r*_*g*_^2^ = 0.26), obesity-glucose (*r*_*g*_^2^ = 0.26), type 2 diabetes-body WHR (*r*_*g*_^2^ = 0.36), type 2 diabetes-BMI (*r*_*g*_^2^ = 0.34), type 2 diabetes-triglycerides (*r*_*g*_^2^ = 0.31), gastroesophageal reflux disease (GERD)-depression (*r*_*g*_^2^ = 0.40), gastroesophageal reflux disease (GERD)-respiratory diseases (*r*_*g*_^2^ = 0.39) and depression-respiratory diseases (*r*_*g*_^2^ = 0.32). These findings support the use of an estimand based on *r*_*g*_^2^ < 0.5 between target and auxiliary diseases/traits. Third, we compared *h*^2^_*pleio*_/*h*^2^ to the sum of (bias-corrected) *r*_*g*_^2^ across auxiliary diseases, across auxiliary disease sets for a given target disease. For each of 15 UK Biobank target diseases, we considered 14 auxiliary disease sets (containing 1, …, 14 auxiliary diseases), starting with the auxiliary disease with highest *r*_*g*_^2^ with the target disease and iteratively adding auxiliary diseases in the order of decreasing *r*_*g*_^2^ with the target disease; we then computed the correlation between *h*^2^_*pleio*_/*h*^2^ and the sum of (bias-corrected) *r*_*g*_^2^ across the 14 auxiliary disease sets. The average correlation (across 15 target diseases) was 0.28, implying that *h*^2^_*pleio*_/*h*^2^ captures different information than the sum of (bias-corrected) *r*_*g*_^2^ (**Supplementary Tables 30-31**). Fourth, we estimated genetic correlations by applying BOLT-REML^31^ to individual-data, instead of applying cross-trait LDSC^8^ to summary-level data. We determined that estimates of genetic correlation and *h*^2^_*pleio*_/*h*^2^ were broadly consistent (**Supplementary Figures 21-22, Supplementary Tables 20-21**). Fifth, we estimated genetic correlations using three modified versions of cross-trait LDSC: constraining heritability intercept, constraining genetic covariance intercept, and constraining both intercepts. We determined that all three modified versions deviated from BOLT-REML results in estimates of genetic correlation and *h*^2^_*pleio*_/*h*^2^ (**Supplementary Figures 21-22, Supplementary Tables 20-21**), supporting our primary use of the default version of cross-trait LDSC.

### Application to 30 diseases from publicly available GWAS meta-analyses

We expanded our analyses to 30 highly heritable diseases with publicly available GWAS summary statistics (heritability z-score > 6; observed-scale heritability under case-control ascertainment ranging from 0.007 to 0.821 (with unknown prevalence); *r*_*g*_^2^ < 0.5; average N=483K, spanning ten PheCode disease categories^33^) (**Supplementary Table 3**). These include many diseases (e.g. autoimmune diseases and cancers) that were not included in the 15 diseases in UK Biobank based on the criteria that we applied. For brevity, we subsequently refer to the above 15 as ‘UK Biobank’ diseases and the 30 as ‘non-UK Biobank’ diseases (while duly noting that a subset of the latter includes both non-UK Biobank and UK Biobank data). Genetic correlations across these 30 diseases are reported in **Supplementary Figure 23** and **Supplementary Table 16**. We again observed that most diseases have moderate genetic correlations within disease categories and between disease categories; we note that it is not feasible to estimate corresponding liability-scale phenotypic correlations in the absence of individual-level data.

We first estimated *h*^2^_*pleio*_/*h*^2^ for 15 UK Biobank target diseases with respect to different sets of auxiliary diseases. Results for three representative diseases (T2D, MDD, and HTN), and the average across 15 diseases, are reported in **Figure 6A, Supplementary Figure 24** and **Supplementary Table 22**. Average *h*^2^_*pleio*_/*h*^2^ increased to 42% with respect to 30 non-UK Biobank auxiliary diseases (jackknife s.e.(avg) = 2%, s.d. = 27% across diseases), 44% with respect to all 45 UK Biobank + non-UK Biobank auxiliary diseases (jackknife s.e.(avg) = 5%, s.d. = 31% across diseases), and 49% (jackknife s.e. (avg) = 4%, s.d. = 28% across diseases) with respect to all 45 UK Biobank + non-UK Biobank auxiliary diseases and 17 UK Biobank quantitative traits, as compared to 27% with respect to 15 UK Biobank auxiliary diseases. Several diseases were dominated by pleiotropic heritability, including depression (74%, s.e. 7% w.r.t. all 62 auxiliary traits) and type 2 diabetes (59%, s.e. 4% w.r.t. all 62 auxiliary traits).

**Figure 6.**
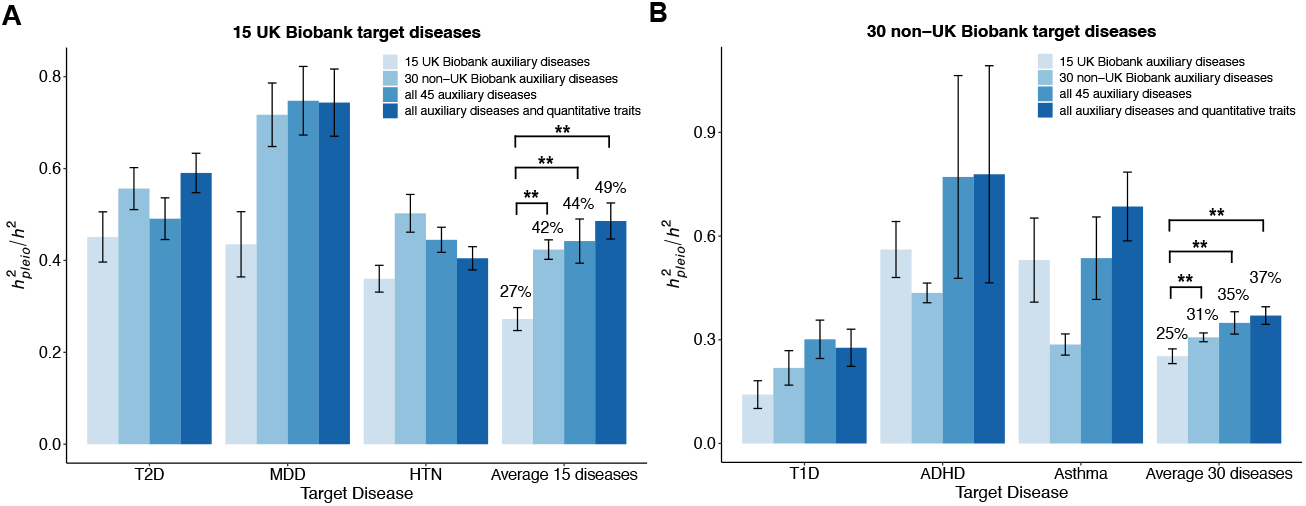
*h*^2^_*pleio*_/*h*^2^ estimates with respect to different sets of auxiliary diseases/traits. (A) *h*^2^_*pleio*_/*h*^2^ estimates for three representative UK Biobank diseases and the average across 15 UK Biobank diseases with respect to four sets of auxiliary diseases/traits. (B) *h*^2^_*pleio*_/*h*^2^ estimates for three representative non-UK Biobank diseases and the average across 30 non-UK Biobank diseases with respect to four sets of auxiliary diseases/traits. Error bars denote jackknife standard errors. To assess statistical significance, we compare average estimates from 30 non-UK Biobank auxiliary diseases, all 45 auxiliary diseases, and all 62 auxiliary diseases and quantitative traits to estimates from 15 UK Biobank auxiliary diseases using inverse-variance weighting across diseases/traits: * *P* < 0.05; \*\**P* < 0.001. Numerical results are reported in **Supplementary Tables 18, 22** and **23**.

We next estimated *h*^2^_*pleio*_/*h*^2^ for 30 non-UK Biobank target diseases with respect to different sets of auxiliary diseases. Results for three representative diseases (type 1 diabetes, attention-deficit/hyperactivity disorder, and asthma), and the average across 30 diseases, are reported in **Figure 6B, Supplementary Figure 25** and **Supplementary Table 23**. We observed a similar trend (increasing and then plateauing as the number of auxiliary traits increases) but with lower overall estimates: average *h*^2^_*pleio*_/*h*^2^ increased to 31% with respect to 30 non-UK Biobank auxiliary diseases (jackknife s.e.(avg) = 1%, s.d. = 21% across diseases), 35% with respect to all 45 UK Biobank + non-UK Biobank auxiliary diseases (jackknife s.e.(avg) = 3%, s.d. = 23% across diseases), and 37% with respect to all 45 UK Biobank + non-UK Biobank auxiliary diseases and 17 UK Biobank auxiliary quantitative traits (jackknife s.e.(avg) = 2%, s.d. = 25% across diseases), as compared to 25% with respect to 15 UK Biobank auxiliary diseases.

Finally, we estimated *h*^2^_*pleio*_/*h*^2^ of 15 UK Biobank target diseases or 30 non-UK Biobank target diseases with respect to auxiliary diseases in each of the 11 PheCode disease categories^33^ spanned by all 45 UK Biobank + non-UK Biobank auxiliary diseases (analogous to **Figure 5**). Results are reported in **Supplementary Figure 26** and **Supplementary Table 24**. We again determined that auxiliary diseases from a single disease category often explain the majority of pleiotropic heritability (e.g. MDD had 68% (s.e. 8%) of SNP-heritability shared with other mental disorders), and that removing a single disease category from the set of auxiliary diseases reduced *h*^2^_*pleio*_/*h*^2^ much less than the *h*^2^_*pleio*_/*h*^2^ shared with that disease category (e.g. MDD *h*^2^_*pleio*_/*h*^2^ was reduced by only 23% (s.e. 6%) when removing other mental disorders).

### Comparing shared genetic and non-genetic variance

We sought to compare *h*^2^_*pleio*_/*h*^2^ to the proportion of liability-scale total phenotypic variance of a target disease that is shared with a given set of auxiliary diseases (pleiotropic phenotypic variance, *V*^2^_*pleio*_; defined in **Methods**, analogous to Eq. 1, **Figure 1B**). We extended our method to estimate *V*^2^_*pleio*_ by replacing genetic correlations (*r*_*g*_) in Eq. 3 with liability-scale phenotypic correlations (*r*_*l*_), estimated from individual-level data as in ref. 31(**Methods**). We determined via simulations that *V*^2^_*pleio*_ estimates are unbiased without the need for bias correction, as *r*_*l*_ is accurately estimated at large sample sizes (**Supplementary Figure 27** and **Supplementary Table 25**). We used the ratio of pleiotropic phenotypic variance vs. total liability-scale phenotypic variance (*V*^2^_*pleio*_/*V*^2^) to quantify the proportion of total liability-scale phenotypic variance that is pleiotropic (noting that *V*^2^ = 1 in standard parametrizations). Because estimating *r*_*l*_ requires individual-level data, we analyzed 15 UK Biobank diseases only (**Table 1**).

A comparison of estimates of *h*^2^_*pleio*_/*h*^2^ vs. *V*^2^_*pleio*_/*V*^2^ is reported in **Figure 7** and **Supplementary Table 26**; to ensure a fair comparison, we pruned the same auxiliary diseases in estimates of *V*^2^_*pleio*_/*V*^2^ as in our prior estimates of *h*^2^_*pleio*_/*h*^2^. We determined that estimates of *h*^2^_*pleio*_/*h*^2^ were generally larger than estimates of *V*^2^_*pleio*_/*V*^2^ (ratio of averages = 1.51x (s.e. 0.14)). *h*^2^_*pleio*_/*h*^2^ was significantly larger than *V*^2^_*pleio*_/*V*^2^ (*P* < 0.05/15) for 5 of 15 diseases, including gastroesophageal reflux disease (GERD) (0.54 (s.e. 0.08) vs. 0.16 (s.e. 0.007)), respiratory diseases (0.53 (s.e. 0.07) vs. 0.16 (s.e. 0.007)), obesity (0.49 (s.e. 0.06) vs. 0.20 (s.e. 0.006)), MDD (0.44 (s.e. 0.07) vs. 0.16 (s.e. 0.007)), and tobacco use disorder (0.36 (s.e. 0.07) vs. 0.14 (s.e. 0.007)). Due to relatively small values of SNP-heritability, estimates of the proportion of non-genetic variance that is shared with auxiliary diseases (*E*^2^_*pleio*_/*E*^2^) were only slightly smaller than *V*^2^_*pleio*_/*V*^2^ estimates on average (**Supplementary Figure 29** and **Supplementary Table 26**). We note that the difference between *h*^2^_*pleio*_ and *V*^2^_*pleio*_ also includes heritability not captured by common SNPs, such as rare variant heritability, although we expect that rare variant heritability is generally relatively small^40–42^.

**Figure 7.**
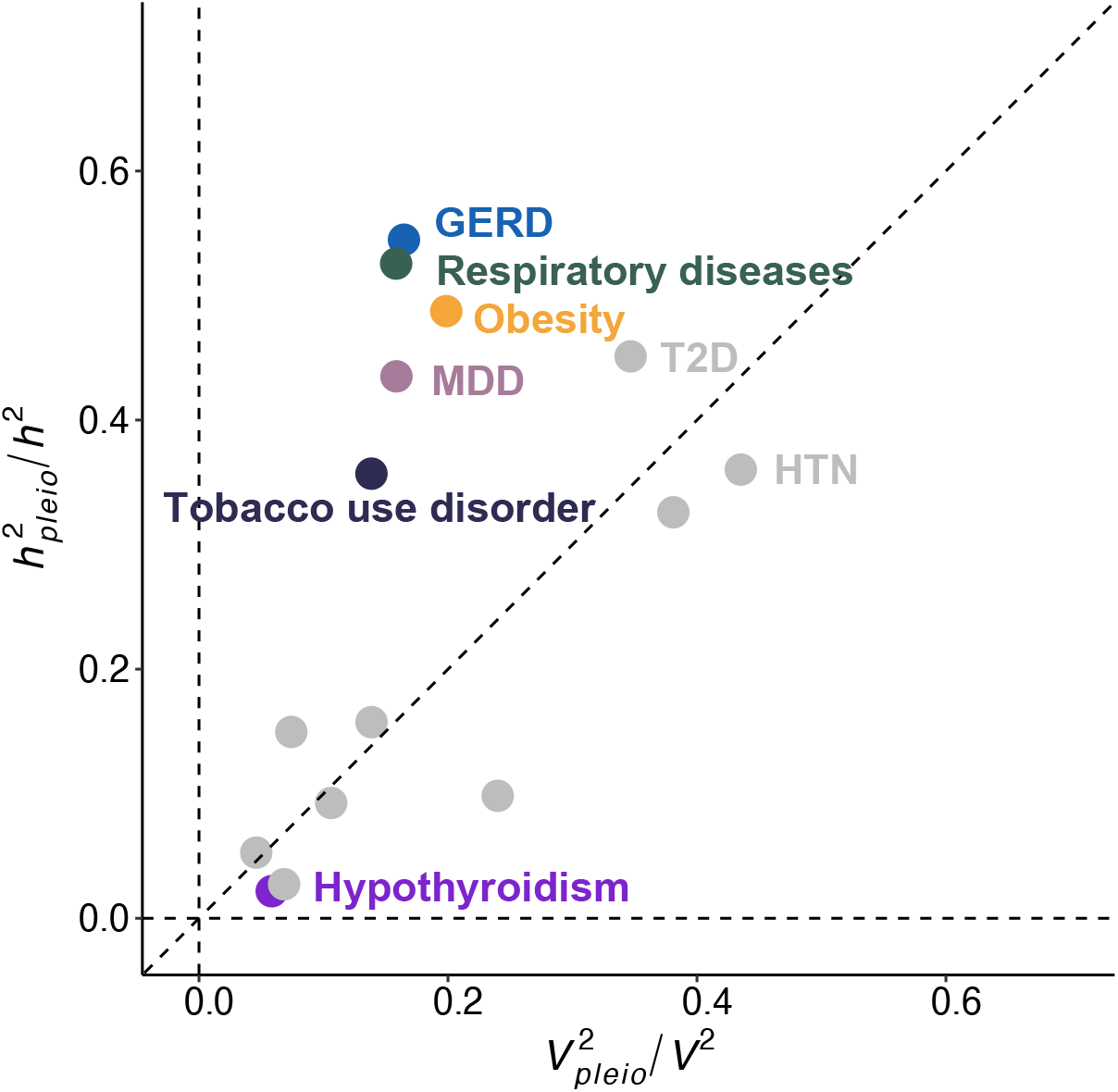
Comparison of *h*^2^_*pleio*_/*h*^2^ vs. *V*^2^_*pleio*_/*V*^2^ for 15 diseases in UK Biobank. Colored dots denote diseases for which *h*^2^_*pleio*_/*h*^2^ estimates are significantly different from *V*^2^_*pleio*_/*V*^2^ estimates (*P* < 0.05/15). Numerical results are reported in **Supplementary Table 26**. MDD: depression; T2D: type 2 diabetes; HTN: hypertension; GERD: gastroesophageal reflux disease.

We performed a secondary analysis to investigate whether there is a correlation between liability-scale heritability and the ratio of *h*^2^_*pleio*_/*h*^2^ to *V*^2^_*pleio*_/*V*^2^ across the 15 UK Biobank diseases. We observed a negative correlation (correlation = −0.65, *P* = 0.008; **Supplementary Figure 32**). The p-value of 0.008 is anti-conservative, as it treats the 15 diseases as independent when they are in fact correlated, and it is thus unclear whether the correlation is statistically significant.

## Discussion

We have defined and estimated *h*^2^_*pleio*_, the genetic variance of a target disease that is shared with a specific set of auxiliary diseases. We highlight four findings. First, bias correction had a large impact on estimates of *h*^2^_*pleio*_. Second, roughly half of the total SNP-heritability of common diseases is shared with other disease/traits. Third, the contribution of auxiliary diseases to *h*^2^_*pleio*_ is broadly distributed across disease categories, with *h*^2^_*pleio*_ decreasing only slightly when removing one auxiliary disease category. Finally, we compared *h*^2^_*pleio*_/*h*^2^ to *V*^2^_*pleio*_/*V*^2^ and determined that genetic variance is more strongly shared across diseases than environmental variance; this finding could be explained by pleiotropic stabilizing selection on complex traits, in which common variants with effects on multiple traits are preferentially retained^43^.

We emphasize four downstream implications. First, our results can inform the incorporation of genome-wide priors in polygenic risk scores that leverage pleiotropic diseases/traits^26,44–48^. Second, our work motivates the definition and estimation of *partitioned pleiotropic heritability*, in which genetic values of auxiliary diseases/traits are partitioned into tissue/cell-type-specific components^49–51^, increasing shared genetic variance with the target disease and elucidating shared biological processes^1–6,15,22,23^. Third, genetic values of auxiliary traits could also be partitioned into gene pathways, elucidating shared biological pathways^52–54^. As such, our method for pleiotropic heritability may be developed in further ways to inform risk prediction, drug repurposing, prioritization of drug targets for multimorbid diseases, or improve the detection of unintended drug side-effects. Finally, pleiotropic heritability could be applied to high-dimensional auxiliary traits, such as brain imaging traits in studies of psychiatric disease^55,56^.

We note several limitations of our work. First, jackknife standard errors on estimates of *h*^2^_*pleio*_ do not account for stochasticity that may in principle be introduced by pruning genetically correlated auxiliary diseases to ensure numerical stability; however, we anticipate that the set of pruned diseases would exhibit minimal variation across genomic jackknife blocks, due to limited variation in genetic correlation estimates across blocks. Second, the definition of *h*^2^_*pleio*_ is with respect to a specific set of auxiliary traits, and results depends on the set of auxiliary traits; however, the magnitude of the increase in *h*^2^_*pleio*_/*h*^2^ plateaus as the number of auxiliary traits increases (**Figure 6A** and **Figure 6B**), implying that expanding the set of auxiliary traits is unlikely to increase the *h*^2^_*pleio*_/*h*^2^ to the level of T2D and MDD for most other diseases; in addition, removing disease categories from the set of auxiliary diseases does not reduce *h*^2^_*pleio*_/*h*^2^ much, implying that individual auxiliary diseases do not have a large impact on *h*^2^_*pleio*_/*h*^2^ when a sufficiently large set of auxiliary diseases are included. Third, genetic correlation estimates include the effects of assortative mating, which can be substantial for some diseases/traits^57^ (e.g. educational attainment); our results do not distinguish between genetic correlation due to shared causal effects vs. genetic correlation due to assortative mating, which would require applying new methods to individual-level data^58^; however, we believe that cross-trait assortative mating effects involving disease risk (e.g. with educational attainment) are likely to be modest in most cases^57^. Fourth, definition and estimation of *h*^2^_*pleio*_ is with respect to a specific phenotype definition in a specific cohort. Ascertainment bias will carry through to our definition and estimation of *h*^2^_*pleio*_; in particular, UK Biobank is known to be subject to participation bias^59,60^. Phenotypic misclassification will also carry through to our definition and estimation of *h*^2^_*pleio*_; we have restricted our analysis to individuals in UK Biobank with both primary care data and hospital record data to limit the impact of phenotypic misclassification. However, shared controls do not impact definition or estimation of *h*^2^_*pleio*_, because definition is with respect to a population with infinite sample size and cross-trait LD score regression, which we use to estimate genetic correlations, is robust to shared controls in a finite sample. Finally, we restricted our analyses to individuals of European ancestry (represented in large sample size in UK Biobank), but it is important to analyze more diverse cohorts^61,62^, particularly for diseases whose prevalence varies across populations. Despite these limitations, our definition and estimation of *h*^2^_*pleio*_ provide a robust quantification of the genetic variance of a target disease that is shared with pleiotropic diseases/traits.

## Code Availability

We have released open-source software implementing PHBC at https://github.com/yjzhao1004/pleioh2g, and published on CRAN (https://cran.r-project.org/web/packages/pleioh2g). Other publicly available software packages used in this study are listed below: GWAS summary statistics were computed using PLINK2 (https://www.cog-genomics.org/plink/2.0/) and BOLT-LMM v2.4.1 (https://alkesgroup.broadinstitute.org/BOLT-LMM/downloads/), Heritability and genetic correlation were estimated using LDSC (https://github.com/bulik/ldsc), the ldscr R package (https://github.com/mglev1n/ldscr), and BOLT-REML as implemented in BOLT-LMM v 2.4.1.

## Data Availability

Summary association statistics for all diseases/traits analyzed in this study have been made publicly available at https://alkesgroup.broadinstitute.org/PHBC/.

## Acknowledgements

Use of the UK Biobank data was performed under application number 19542. This research was funded by Wellcome Trust early-career award 227566/Z/23/Z (X.J.) and NIH grants R01 HG006399, U01 HG012009, R01 MH101244, and R37 MH107649 (A.L.P.). M.I. is supported by the Munz Chair of Cardiovascular Prediction and Prevention and the NIHR Cambridge Biomedical Research Centre (NIHR203312) as well as by the UK Economic and Social Research 878 Council (ES/T013192/1). J.D. is supported by a British Heart Foundation Personal Chair. This work was supported by core funding from the British Heart Foundation (RG/18/13/33946: RG/F/23/110103), NIHR Cambridge Biomedical Research Centre (NIHR203312), BHF Chair Award (CH/12/2/29428), Cambridge BHF Centre of Research Excellence (RE/24/130011, RE/18/1/34212) and by Health Data Research UK, which is funded by the UK Medical Research Council, Engineering and Physical Sciences Research Council, Economic and Social Research Council, Department of Health and Social Care (England), Chief Scientist Office of the Scottish Government Health and Social Care Directorates, Health and Social Care Research and Development Division (Welsh Government), Public Health Agency (Northern Ireland), British Heart Foundation and the Wellcome Trust. The views expressed are those of the authors and not necessarily those of the NIHR or the Department of Health and Social Care. W.C. is supported by grants from the National Key R&D Program of China (No. 2023YFC3605400) and the National Natural Science Foundation of China (No. 82472055, No. 82071997).

## Methods

### Ethics statement

The UK Biobank has received ethical approval from the National Information Governance Board for Health and Social Care and the National Health Service North West Centre for Research Ethics Committee (ref. 11/NW/0382).

### Definition of *h*^2^_*pleio*_

We assume that the total liability of the target disease consists of genetic value (G) and environmental value (E). G of the target disease is divided into two components: private (disease-specific) genetic value (*G*_*private*_) and pleiotropic genetic value (*G*_*pleio*_), with the pleiotropic genetic value defined as a weighted linear combination of auxiliary disease genetic values (**Figure 1A**):

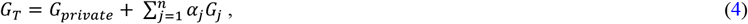

where *α*_*j*_ is the weight of the *j*^.*th*^ auxiliary disease (*D*_*j*_), *G*_*j*_ is the genetic value of *D*_*j*_, and *G*_*T*_is the genetic value of target disease.

We define pleiotropic heritability as the maximum proportion of the total target disease heritability explained by the auxiliary diseases in the entire population. We use the ratio of pleiotropic heritability vs. total heritability 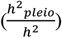 to quantify the proportion of genetic variance that is pleiotropic:

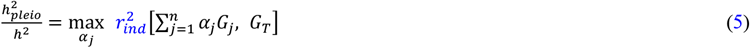

where *r*_*ind*_ denotes the correlation across individuals. We emphasize that *h*^2^_*pleio*_ is a function of both the target disease and the selected set of auxiliary diseases/traits.

Analogously, we assume that standardized causal SNP effect sizes (number of standard deviation increase in phenotype per 1 standard deviation increase in genotype) on the target disease consist of two components: disease-specific effect sizes (*β*_*disease-specific*_) and pleiotropic effect sizes (*β*_*pleio*_), with the pleiotropic effect sizes defined as a weighted linear combination of auxiliary disease effect sizes:

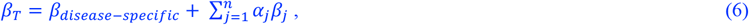

where *α*_*j*_ is the weight for the *j*^.*th*^ auxiliary disease (*D*_*j*_), *β* is the vector of additive causal SNP effect sizes (*β* = (*β*_1_, *β*_2_, …, *β*_*m*_), where *m* denotes the number of SNPs), *β*_*T*_is the vector of SNP effects on the target disease, and *β*_*j*_ is the vector of SNP effects on the *j*^.*th*^ auxiliary disease. In the general case that the genotype data does not include all causal SNPs, *β* are best-fit effect sizes obtained by projecting the phenotype onto the genotype space in the infinite population.

We restate the definition of equation (2):

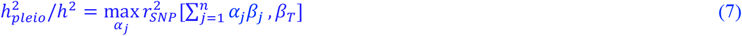

where *r*_*SNP*_ denotes the correlation across SNPs. We note that equation (7) applies equally to causal SNP effect sizes on either the liability or observed scale, as these effect sizes differ only by a constant scaling factor, and the squared correlation *r*^2^ is invariant under linear rescaling.

We note the definition in equation (1) and (2) are equivalent under the assumptions that

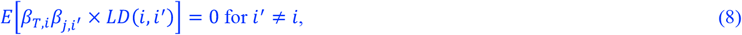

where *β* denotes the causal SNP effect sizes defined under a random effect model, *j* is the disease index (including the target disease), *i, i*′ are SNP indices, and *LD* is the correlation between genotypes. Intuitively, the assumption requires the causal effect sizes of SNPs that are in LD with each other to be uncorrelated. We note that causal effect sizes for different SNPs have been shown to have minimal correlations, particularly among common SNPs^63^.

In detail, we model genetic value using standardized genotypes *X* and causal SNP-effect vector *β*. The genetic value for the target disease is *G*_*T*_= *Xβ*_*T*_and the weighted linear combination of genetic value for *n* auxiliary disease is 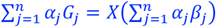. Using the property of the trace operator:

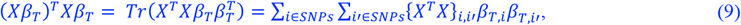

and

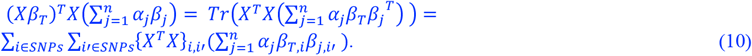

Using the assumption of equation (8), ∑_*i*≠*i*′_{*X*^*T*^*X*} _*i,i*′_*β*_*T,i*_*β*_*T,i*′_= 0 and 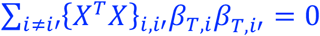 when the number of SNP goes to infinity, yielding:

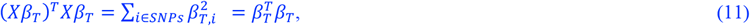

And

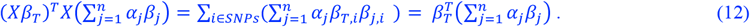

Therefore, the covariance between the target genetic value and weighted auxiliary disease genetic value is

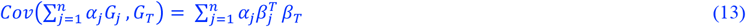

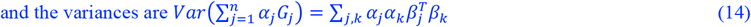

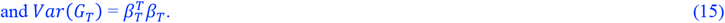

Therefore, the squared correlation of genetic value across individuals 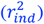 in a population of infinite sample size equals the squared correlation across SNPs 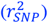:

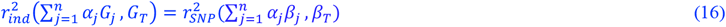

### Initial estimate of *h*^2^_*pleio*_

We employ pleiotropic heritability with bias correction (PHBC) to obtain an unbiased estimate of 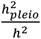 with finite sample sizes.

According to the definition in equation (7), pleiotropic heritability is not changed when multiplying ***α*** by a non-zero scaling factor. We note that the quantity in equation (7) is equivalent to regression ***R***^**2**^, therefore we change the optimization to:

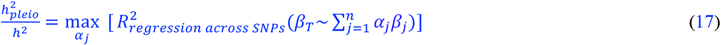

so that the *α*_*j*_ are identifiable. We derive the expression of 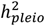 as follows:

Let ***α***^∗^ be the estimated ***α*** that maximizes the right-hand side of equation (17).

Let ***C***_*DD*_ be the *n* × *n* covariance matrix of 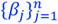

Let ***c***_*TD*_ be the *n* × 1 covariance vector between *β* and 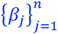

Under the assumption of equation (8), the variance of pleiotropic genetic value 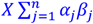 is:

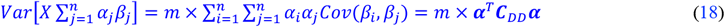

To estimate *h*^2^_*pleio*_, we first estimate the ***α***^∗^ by minimizing the squared error between *β*_*T*_and 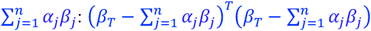 (which is equivalent to maximizing regression ***R***^**2**^). The parameters are estimated across all SNPs which is sufficiently large that prevents overfitting. By taking the partial derivative of 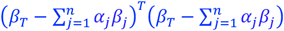 with respect to ***α***:

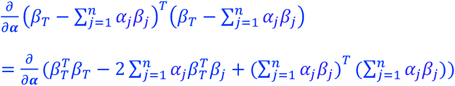

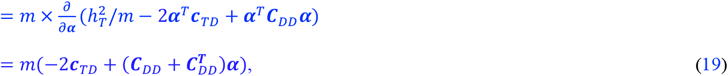

where *m* is the number of SNPs, the expectation of each SNP effect is 0. By setting the partial derivative to zero, we can obtain the optimal weight vector for each SNP:

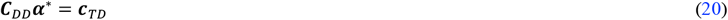

If ***C***_*DD*_ is invertible, the optimal ***α***^∗^ is:

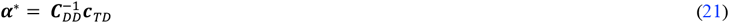

After substituting the ***α***^∗^ into equation (18):

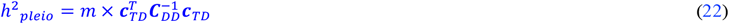

Elements in ***c***_*TD*_ and ***C***_*DD*_ denote per-SNP heritability and genetic covariance; in this study we focus on SNP-heritability and genetic correlation between causal SNP effect sizes. Let 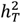 be the heritability of the target disease. 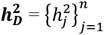 is the vector consisting of the heritability of all auxiliary diseases. *r*_*g*_[***D***, *T*] is the genetic correlation vector between the target disease and all auxiliary diseases, *r*_*g*_[***D, D***]^−1^ is the inverse of genetic correlation matrix of all auxiliary diseases. diag(.) is diagonal matrix with all non-diagonal elements equal to zero.

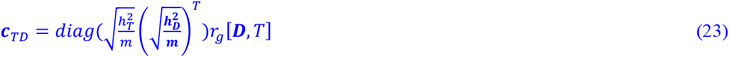

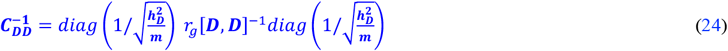

After substituting equation (23) and (24) into equation (22), *h*^2^_*pleio*_ can be expressed as:

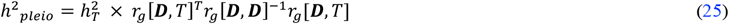

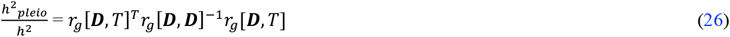

Equation (26) provides the initial pleiotropic heritability estimates, prior to bias correction described in the next section. In this study, we used GWAS summary statistics as input and reimplemented cross-trait LDSC^8^ so that the standard errors of all quantities on the right-hand side of equation (26) are estimated through the same genomic jackknife blocks. We used 1000 Genomes Project^64^ Europeans as a reference LD panel to estimate genetic correlation. Standard error estimates were obtained via genomic block-jackknife with 200 blocks applied across all traits, which were created by partitioning 1,217,311 HapMap 3 SNPs^36^. We primarily focus on *h*^2^_*pleio*_/*h*^2^ estimates (instead of *h*^2^_*pleio*_ estimates) throughout the manuscript.

### Monte Carlo bias correction to initial estimate of *h*^2^_*pleio*_

The initial estimates 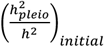 are based on estimates of genetic correlation in finite GWAS sample size, which introduces an upward bias in estimating 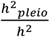 due to inverting the estimated genetic correlation matrix and sampling correlation between *r*_*g*_[***D***, *T*]^*T*^, *r*_*g*_[***D, D***] ^− 1^, and *r*_*g*_[***D***, *T*] in equation (26):

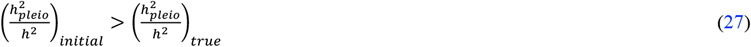

Specifically, if only one auxiliary disease is included, *h*^2^_*pleio*_/*h*^2^ will be equivalent to 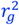; the expectation of the squared genetic correlation estimates 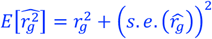 will be larger than the square of true genetic correlation 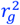, indicating an upward bias in *h*^2^_*pleio*_/*h*^2^. If multiple auxiliary diseases are included, the expectation of Eq. 3 is upward biased for noisy 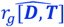 when *r*_*g*_[***D, D***] ^− 1^ is positive definite: 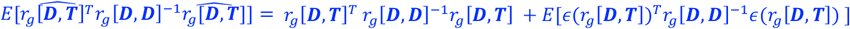, where *ε*(*r*_*g*_[***D, T***]) is the estimation error of 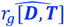; we used 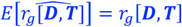. Since *r*_*g*_[***D, D***]^− 1,^ is the estimate of the inverse of a genetic correlation matrix which is positive semi-definite, it is likely positive semi-definite. Therefore, the second term in this equation is non-negative, likely causing an upward bias in our initial estimate of *h*^2^_*pleio*_/*h*^2^ in Eq. 3.

To obtain an unbiased estimate, we employ a Monte-Carlo bias correction procedure to estimate the proportion of bias in the point estimate of 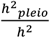 and then correct the bias. First, we generate Monte-Carlo samples of the estimation error *ε*_*i*_ in the genetic correlation matrix from the sampling covariance matrix^17^ generated from genomic block-jackknife; if the dimension of the genetic correlation matrix is *n* × *n*, the sampling covariance matrix is a symmetric matrix with dimension *n*(*n*-1)/2 × *n*(*n*-1)/2, which captures the joint distribution across estimation errors of different elements in *r*_*g*_[***D***, *T*] and *r*_*g*_[***D, D***]. Second, we add the estimation noise to the point estimate of genetic correlation matrix to create Monte-Carlo samples of estimated *r*_*g*_[***D***, *T*] and *r*_*g*_[***D, D***]^− 1^. We filter these Monte-Carlo samples to only keep those whose *r*_*g*_[***D, D***] are non-singular (i.e. the smallest eigenvalue of *r*_*g*_[***D, D***] is larger than 0). We repeat this Monte-Carlo procedure until we have generated 1,000 samples of genetic correlation estimates.

Using these Monte-Carlo samples, we next estimate a scaling coefficient *ξ*_*c*_ so that:

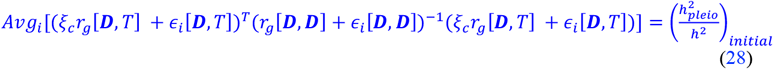

The left-hand side is the average across Monte-Carlo samples, *ε*_*i*_ are the respective Monte-Carlo samples of estimation errors for *r*_*g*_[***D***, *T*] and *r*_*g*_[***D, D***], where *i* indexes Monte-Carlo samples. The above equation is based on the heuristic that the magnitude of pleiotropic heritability is proportional to the average genetic correlation between target disease and auxiliary diseases *r*_*g*_ [***D***, *T*]. Downward scaling *r*_*g*_ [***D***, *T*] will downward scale 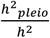, which controls the upward bias in equation (27). On the other hand, *r*_*g*_[***D, D***] are between pairs of auxiliary diseases which is not directly connected to the scale of 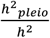. The error term *ε*_*i*_ is not multiplied by *ξ*_*c*_ as estimation error for genetic correlation is a function of sample size and the heritability, which is not proportional to the scale of the point estimate of genetic correlation.

We estimate scaling coefficient *ξ*_*c*_ using binary search on the interval [0,1], in order to satisfy equation (28). We multiply the initial estimate 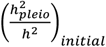 and its standard error by 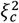 to obtain the bias-corrected estimate of *h*^2^_*pleio*_/*h*^2^ and its standard error.

### Pruning auxiliary diseases with high genetic correlation

In the presence of collinearity among auxiliary diseases and large uncertainty in genetic correlation estimates, the estimator in equation (26) can be numerically unstable, which we define as (1) The pre-correction jackknife standard error of 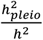 with respect to the pre-pruning auxiliary disease sets is greater than 0.5, and/or (2) the scaling coefficient *ξ*_*c*_ is less than 0.5 (implying that >75% of the initial 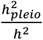 estimate is due to uncertainty in genetic correlation matrix estimates). We also apply the pruning procedure if (3) it takes more than 50 iterations to sample a non-singular *r*_*g*_[***D, D***] in the Monte-Carlo sampling of bias correction, which indicates that the collinearity of auxiliary diseases causes singularity in samples of *r*_*g*_[***D, D***].

To apply the pruning procedure, we prune the auxiliary diseases by progressively applying stricter 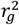 thresholds between the auxiliary diseases: 0.5, 0.4, 0.3, 0.2, and 0.1. For each 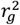 threshold, when a pair of auxiliary diseases has 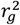 above the threshold, we keep the disease with larger *r*_*g*_ z-score with the target disease. We repeat this pruning step until all three criteria no longer hold, and we report the 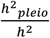 estimate from the pruned set of auxiliary diseases.

### Pleiotropic phenotypic variance

To compare *h*^2^_*pleio*_/*h*^2^ with the non-genetic variance that can be explained by the same set of auxiliary diseases, we extend our method to estimate pleiotropic phenotypic variance (*V*^2^_*pleio*_), defined as the proportion of phenotypic variance of target disease liability (*σ*_*T*_) that can be explained by any linear combination of liabilities of auxiliary disease (*σ*_*D*_) (**Figure 1B**).

We convert the observed-scale phenotypic correlation to liability-scale phenotypic correlation *r*_*l*_ using Monte-Carlo simulations^31^ under the liability threshold model^65^. To model the relationship between liability-scale correlation *r*_*l*_ and observed-scale correlations *r*_*o*_ for two binary traits with different prevalences, we simulate liabilities using *r*_*l*_ on a grid between −1 to 1. For each *r*_*l*_, we simulate pairs of disease liabilities from a multivariate normal distribution with covariance equal to specified *r*_*l*_, and then use liability threshold model to create the binary disease status by thresholding the liabilities based on their respective prevalences. We then calculate the *r*_*o*_ between the simulated binary traits. This process generates a map between *r*_*l*_ and *r*_*o*_ for two disease traits of given prevalences. Using this mapping, we can map the observed *r*_*o*_ for the 15 UK Biobank diseases to obtain the corresponding *r*_*l*_. We use the *r*_*l*_ matrix to estimate *V*^2^_*pleio*_ in the same way as pleiotropic heritability:

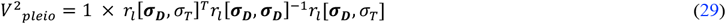

where 1 is the total liability variance for the target disease, *r*_*l*_[***σ***_***D***_, *σ*_*T*_] is the liability-scale phenotypic correlation matrix between target disease and auxiliary diseases, *r*_*l*_[***σ***_***D***_, ***σ***_***D***_] ^−1^ is the inverse of liability-scale phenotypic correlation among auxiliary diseases. We estimate the standard error of *r*_*l*_ and *V*^2^_*pleio*_ by jackknifing over 1000 blocks of individuals across all diseases. We also compare the difference between *r*_*g*_ and *r*_*l*_ with Bonferroni-corrected significant threshold set at *P* < 4.76 × 10^−4^ (0.05/105, where 105 is the number of pairs of the 15 diseases) (**Figure 3** and **Supplementary Table 15**).

We compare three quantities: 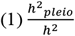, the proportion of genetic variance that is pleiotropic; 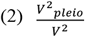, the proportion of liability variance that is shared with the set of auxiliary diseases, and 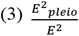, the proportion of non-genetic variance (including rare variant heritability and environmental variance) that is shared with the set of auxiliary diseases, calculated as:

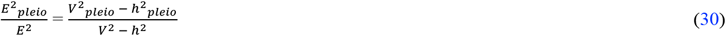

We computed the ratios between 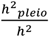 vs. 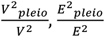 vs. 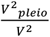 vs. *V*^2^_*pleio*_, and 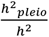 vs. 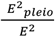. Full results are listed in **Supplementary Figures 28-31** and **Supplementary Table 26**.

### Simulations

We performed simulation using real UK Biobank genotypes from 1,122,329 HapMap3^36^ SNPs across 157,206 unrelated individuals (see below). We simulated 16 diseases with liability-scale heritabilities equal to 0.13 (median of 15 UK Biobank diseases) and prevalences equal to 0.1 (median of 15 UK Biobank diseases); we have also included simulations with heritability z-scores matching the median value of 15 UK Biobank diseases (**Supplementary Figure 8** and **Supplementary Figure 10**). True genetic correlations were set to 0.5 within diseases categories and 0.1 between diseases categories for the first 15 diseases (based on the 7 PheCode disease categories from **Table 1**) and 0.0 for the 16th disease (with all other 15 diseases), which results in 5 different values of true 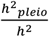. The proportion of causal SNPs was set to 5% in the primary simulations (and 1% in a secondary simulation). For each simulation, we first simulated the causal effect sizes using infinitesimal model based on the specified heritability and genetic correlation. Then, we used PLINK2^37^ to compute genetic values (which sum over causal_effect_size × causal_allele_dosage) for 157,206 unrelated individuals and added environmental noise to generate the individual liabilities for the 16 diseases. We used the liability threshold model to generate binary phenotype based on the specified prevalence and computed GWAS summary statistics using PLINK2^37^. We applied cross-trait LDSC^8^ to the GWAS summary statistics for the 1,121,509 SNPs (MAF > 0.01) and used reference LD from 1000 Genomes Europeans individuals (9,254,535 SNPs) to estimate genetic correlations. We applied PHBC to estimate 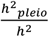 and compared it with the true value. We also compared the squared genomic-jackknife standard error of 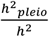 with the squared deviation (defined as the squared difference between the estimated post-corrected 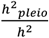 and true value) to investigate whether the reported standard errors were well-calibrated.

We also performed a simulation to validate *V*^2^_*pleio*_ estimation. True *r*_*o*_ was set to 0.5 within diseases categories and 0.1 between diseases categories for the 15 diseases (based on the 7 PheCode disease categories). We simulated liabilities for 228,258 individuals (see below). We computed true *r*_*l*_ and true *V*^2^_*pleio*_ based on the simulated liabilities. Then, we used the liability threshold model to generate binary phenotypes based on the empirical prevalences of the 15 UK Biobank diseases. We applied our method to the simulated binary phenotypes to estimate *V*^2^_*pleio*_ and compared it to the true value.

### UK Biobank data

For disease phenotype pre-processing, we collected diagnoses from both inpatient data and primary care data. An individual is defined as a case if there is a diagnosis from either inpatient data or primary care data. We selected 228,258 samples with both primary care and hospital inpatient records. Diagnoses from hospital inpatient records were obtained from Hospital Episode Statistics (HES) for England, which are recorded as the International Classification of Diseases (ICD-10) system codes. Diagnoses from primary care data were retrieved from a subset of UK Biobank samples (see below), which are recorded in Read Codes v.2 (Read v.2) and Read Codes Clinical Terms v.3 (Read CTv.3). We mapped primary care data from Read v.2 to ICD-10 codes, and combine with the ICD-10codes from HES data. We kept ICD-10 codes starting with the letters A to N which are disease codes. Then, we mapped the ICD-10 records to PheCode system to obtain the phenotype definition^33,66^ and selected the PheCode phenotypes with > 1% prevalence in the 228,258 samples.

We selected 15 relatively independent heritable diseases with heritability *z-*scores larger than 6 (heritabilities were computed using cross-trait LDSC^8^), which are distributed across 7 PheCode disease categories. We note that we did not include any UK Biobank autoimmune diseases or cancers, because they did not meet our criteria of prevalence > 1% and heritability z-score > 6. We used 228,258 samples to compute pleiotropic phenotypic variance across these 15 UK Biobank disease phenotypes. Detailed information on phenotype codes, prevalences, and categories are listed in **Table 1**.

We selected 157,206 unrelated (defined as having less than 11 putative third-degree relatives in the kinship table) British ancestry individuals from the 228,258 samples to compute GWAS summary statistics for 15 UK Biobank diseases, where we used 1,141,346 HapMap 3 SNPs (see below; 1,117,934 of them overlap with the SNPs used in simulations)^36^.We used BOLT-LMM^67,68^ to compute summary statistics. Since BOLT-LMM test statistics are well-calibrated for traits with a case fraction of at least 10%^68^, for diseases with prevalence lower than 10% we matched each case with a subset of nine randomly sampled controls, to avoid miscalibration in unbalanced case-control samples.

Detailed genotyping and quality control procedures in the UK Biobank are described previously^32^. We excluded SNPs with call rates of < 95%, minor allele frequency of < 0.1%, deviation from the Hardy–Weinberg equilibrium with P < 1 × 10 ^− 10^. We then generated the PLINK-format data for BOLT-LMM using two steps: 1) subset to the 157,206 unrelated British individuals with genotype data; 2) subset the SNPs to HapMap3 SNPs and using PLINK2 to LD prune at *r*^2^<0.8 to obtain the set of relatively independent SNPs (PLINK2 command “-indep-pairwise 50 5 0.8”). 470,851 SNPs from 157,206 samples remained after the pruning step, and were carried forward to BOLT-REML for variance components analysis; we reused the full 1,141,346 HapMap 3 SNPs to compute the BOLT-LMM association test statistics.

We selected 17 quantitative traits for relevance to disease from ref. 34,35 for the same set of individuals (Average *N* = 230K) and corrected for medication use, as follows. We corrected blood biochemistry measurements for cholesterol, hypertensive, and diabetic medications: total cholesterol, LDL, and triglyceride levels were corrected for lipid lowering medications^69^; systolic blood pressure and diastolic blood pressure were corrected for hypertension medications^70^; and HbA1c level was corrected for non-insulin diabetic medications and insulin drugs. We verified that the correlation between the corrected quantitative traits and corresponding binary traits was higher compared to uncorrected quantitative traits. We also included educational attainment (years of education) to evaluate the impact of socio-economic status. Detailed information on trait names, categories, heritability estimates and genetic correlation estimates for these 17 quantitative traits is provided in **Supplementary Tables 2** and **16**.

### GWAS summary statistics for 30 diseases from publicly available GWAS meta-analyses

We collected publicly available GWAS summary statistics for 30 relatively independent heritable diseases under the same criteria (heritability z-score > 6 and *r*_*g*_^2^ < 0.5) that are primarily from European ancestry. We note that some of these 30 publicly available disease summary statistics are meta-analyses that include UK Biobank samples. The 30 diseases are assigned to 10 PheCode categories^33^. Details of phenotype information, heritability, and genetic correlation estimates from cross-trait LDSC for these 30 diseases are reported in **Supplementary Tables 3** and **16**.

### Analyses with one or more disease categories excluded from the set of auxiliary diseases

We performed leave-category-out analyses to investigate the change in 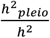 when removing PheCode Categories. First, we estimated the 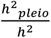 with respect to three auxiliary disease sets: 1) all auxiliary diseases; 2) all diseases excluding the auxiliary diseases in the target disease category; 3) all diseases excluding the auxiliary diseases in target disease category and one other category. We implemented leave-category-out analyses in the PHBC software.

To evaluate the contribution of a single PheCode category to 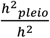, we computed the reduction of 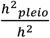 when removing the auxiliary PheCode category. To remove the impact of randomness in Monte-Carlo bias correction when computing the reduction, we first computed the difference of 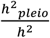 with respect to all auxiliary diseases vs. 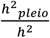 with respect to auxiliary diseases excluding a PheCode category before bias correction. We multiplied the difference by the scaling coefficient 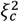 of 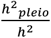 with respect to all auxiliary diseases, which represented the contribution of an auxiliary PheCode category to the 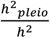 with respect to all auxiliary diseases. We reported the jackknife standard error of the before-bias-correction difference of 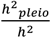 as the standard error of the reduction, which is a conservative standard error shown by simulations (**Supplementary Figure 14** and **Supplementary Table 13**).

### Alternative approaches for estimating *r*_*g*_

To validate the *r*_*g*_ estimated from cross-trait LDSC without constrained intercept, we used BOLT-REML and cross-trait LDSC using constrained intercept to estimate *r*_*g*_. We applied BOLT-REML to individual-data using 470,851 SNPs across 157,206 unrelated individuals of British ancestry to estimate *r*_*g*_ among these 15 UK Biobank diseases. We constrained heritability intercept in cross-trait LDSC to be 1. We also computed the analytical intercept for genetic covariance considering sample overlap for each disease pair using equation 16 in the Supplementary Note of ref. 8. The constrained intercepts for genetic covariance for each disease pair are provided in **Supplementary Table 27**.

