## Supplementary Materials (Supplementary Table captions and Supplementary Figures) for "Pleiotropic heritability quantifies the shared genetic variance of common diseases"

**Supplementary Table 1. List of 15 UK Biobank diseases in primary analyses**. We select 15 relatively independent heritable UK Biobank diseases with > 1% prevalence in 228,258 samples (see **Methods**) and heritability *z* scores larger than 6. We restrict their squared genetic correlations to be smaller than 0.5 among 15 diseases (**Supplementary Table 14**) (both heritability and genetic correlation are computed using cross-trait LDSC). We obtain their PheCode definitions by mapping their ICD-10 records to PheCode system (See **Methods**). These 15 diseases are distributed across 7 PheCode disease categories. Detailed sample size using in computing GWAS are described in **Methods**. We report the disease name, PheCode, observed-scale and liability-scale SNP-heritability, standard error, z-score, category, prevalence and ICD-10 code.

**Supplementary Table 2. List of 17 UK Biobank quantitative traits**. We select 17 relatively independent heritable UK Biobank quantitative traits with heritability *z* scores larger than 6 and squared genetic correlation smaller than 0.5 (**Supplementary Table 16**). The preprocess of these 17 traits is described in **Methods**. We report the trait name, observed-scale SNP-heritability, standard error, z-score, and category.

**Supplementary Table 3. List of 30 diseases from publicly available GWAS meta-analyses**. We selected publicly available GWAS summary statistics from 30 relatively independent heritable diseases with heritability *z* scores larger than 6 and squared genetic correlation smaller than 0.5 (**Supplementary Table 16**). These 30 GWAS summary statistics are all from European ancestry. For brevity, we subsequently refer to the 30 as ‘non-UK Biobank’ diseases (while duly noting that a subset of the latter includes both non-UK Biobank and UK Biobank data). These 30 non-UKB diseases are assigned to 10 PheCode categories. We reported the trait name, trait identifier used to index traits in plots and tables, source reference, sample size, observed-scale SNP-heritability, standard error, z-score and category.

**Supplementary Table 4. Simulation results when $r_g$ within-disease categories equals to 0.5 and $r_g$ between-disease categories equals to 0.1.** The first column for each sub-table lists the disease PheCodes that have the same true ${h^2}_{pleio}/h^2$ based on the specified genetic architecture. In the top two sub-tables, we show the estimated ${h^2}_{pleio}/h^2$, true ${h^2}_{pleio}/h^2$ and their absolute bias for 5 different values both before bias correction and after bias correction. Estimated ${h^2}_{pleio}/h^2$ before bias correction shows an upward bias, but it is approximately unbiased after bias correction. These results correspond to **Figure 2**. In the bottom two sub-tables, we show the sum of squared deviation vs. the sum of squared jackknife standard errors of $h^2{}_{pleio}/h^2$. Deviation is the difference between true $h^2{}_{pleio}/h^2$ and the average estimated $h^2{}_{pleio}/h^2$ across diseases that have the same true $h^2{}_{pleio}/h^2$. The estimated standard errors of $h^2{}_{pleio}/h^2$ before bias correction is anti-conservative as the sum of squared jackknife standard errors of $h^2{}_{pleio}/h^2$ is smaller than the sum of squared deviation across simulations. The estimated standard errors of $h^2{}_{pleio}/h^2$ after bias correction were conservative as the ratio of estimated standard errors to empirical standard deviations was equal to 1.63 (jackknife s.e. 0.14). These results correspond to **Supplementary Figure 1**.

**Supplementary Table 5. Simulation results when $r_g$ within-disease categories equals to 0.4 and $r_g$ between-disease categories equals to 0.1.** The first column for each sub-table lists the disease PheCodes that have the same true $h^2{}_{pleio}/h^2$ based on the specified genetic architecture. In the top two sub-tables, we show the estimated $h^2{}_{pleio}/h^2$, true $h^2{}_{pleio}/h^2$ and their absolute bias for 5 different values both before bias correction and after bias correction. Estimated $h^2{}_{pleio}/h^2$ before bias correction shows an upward bias, but it is approximately unbiased after bias correction. These results correspond to **Supplementary Figure 4A** and **4B**. In the bottom two sub-tables, we show the sum of squared deviation vs. the sum of squared jackknife standard errors of $h^2{}_{pleio}/h^2$. Deviation is the difference between true $h^2{}_{pleio}/h^2$ and the average estimated $h^2{}_{pleio}/h^2$ across diseases that have the same true $h^2{}_{pleio}/h^2$. The estimated standard errors of $h^2{}_{pleio}/h^2$ before bias correction is anti-conservative as the sum of squared jackknife standard errors of $h^2{}_{pleio}/h^2$ is smaller than the sum of squared deviation across simulations. The estimated standard errors of $h^2{}_{pleio}/h^2$ after bias correction is conservative as the ratio of estimated standard errors to empirical standard deviations was equal to 1.42 (jackknife s.e. 0.10). These results correspond to **Supplementary Figure 5A** and **5B**.

**Supplementary Table 6. Simulation results when $r_g$ within-disease categories equals to 0.3 and $r_g$ between-disease categories equals to 0.1.** The first column for each sub-table lists the disease PheCodes that have the same true $h^2{}_{pleio}/h^2$ based on the specified genetic architecture. In the top two sub-tables, we show the estimated $h^2{}_{pleio}/h^2$, true $h^2{}_{pleio}/h^2$ and their absolute bias for 5 different values both before bias correction and after bias correction. Estimated $h^2{}_{pleio}/h^2$ before bias correction shows an upward bias, but it is approximately unbiased after bias correction. These results correspond to **Supplementary Figure 4C** and **4D**. In the bottom two sub-tables, we show the sum of squared deviation vs. the sum of squared jackknife standard errors of $h^2{}_{pleio}/h^2$. Deviation is the difference between true $h^2{}_{pleio}/h^2$ and the average estimated $h^2{}_{pleio}/h^2$ across diseases that have the same true $h^2{}_{pleio}/h^2$. The estimated standard errors of $h^2{}_{pleio}/h^2$ before bias correction is anti-conservative as the sum of squared jackknife standard errors of $h^2{}_{pleio}/h^2$ is smaller than the sum of squared deviation across simulations. The estimated standard errors of $h^2{}_{pleio}/h^2$ after bias correction is conservative as the ratio of estimated standard errors to empirical standard deviations was equal to 1.54 (jackknife s.e. 0.11). These results correspond to **Supplementary Figure 5C** and **5D**.

**Supplementary Table 7. Simulation results when $r_g$ within-disease categories equals to 0.2 and $r_g$ between-disease categories equals to 0.1.** The first column for each sub-table lists the disease PheCodes that have the same true $h^2{}_{pleio}/h^2$ based on the specified genetic architecture. In the top two sub-tables, we show the estimated $h^2{}_{pleio}/h^2$, true $h^2{}_{pleio}/h^2$ and their absolute bias for 5 different values both before bias correction and after bias correction. Estimated $h^2{}_{pleio}/h^2$ before bias correction shows an upward bias, but it is approximately unbiased after bias correction. These results correspond to **Supplementary Figure 4E** and **4F**. In the bottom two sub-tables, we show the sum of squared deviation vs. the sum of squared jackknife standard errors of $h^2{}_{pleio}/h^2$. Deviation is the difference between true $h^2{}_{pleio}/h^2$ and the average estimated $h^2{}_{pleio}/h^2$ across diseases that have the same true $h^2{}_{pleio}/h^2$. The estimated standard errors of $h^2{}_{pleio}/h^2$ before bias correction is anti-conservative as the sum of squared jackknife standard errors of $h^2{}_{pleio}/h^2$ is smaller than the sum of squared deviation across simulations. The estimated standard errors of $h^2{}_{pleio}/h^2$ after bias correction is conservative as as the ratio of estimated standard errors to empirical standard deviations was equal to 1.51 (jackknife s.e. 0.09). These results correspond to **Supplementary Figure 5E** and **5F**.

**Supplementary Table 8. Simulation results with the true liability-scale heritability set to 0.25 (instead of 0.13).** The first column for each sub-table lists the disease PheCodes that have the same true $h^2{}_{pleio}/h^2$ based on the specified genetic architecture. In the top two sub-tables, we show the estimated $h^2{}_{pleio}/h^2$, true $h^2{}_{pleio}/h^2$ and their absolute bias for 5 different values both before bias correction and after bias correction. Estimated $h^2{}_{pleio}/h^2$ before bias correction shows an upward bias, but it is approximately unbiased after bias correction. These results correspond to **Supplementary Figure 6**. In the bottom two sub-tables, we show the sum of squared deviation vs. the sum of squared jackknife standard errors of $h^2{}_{pleio}/h^2$. Deviation is the difference between true $h^2{}_{pleio}/h^2$ and the average estimated $h^2{}_{pleio}/h^2$ across diseases that have the same true $h^2{}_{pleio}/h^2$. The estimated standard errors of $h^2{}_{pleio}/h^2$ before bias correction is anti-conservative as the sum of squared jackknife standard errors of $h^2{}_{pleio}/h^2$ is smaller than the sum of squared deviation across simulations. The estimated standard errors of $h^2{}_{pleio}/h^2$ after bias correction were conservative as the ratio of estimated standard errors to empirical standard deviations was equal to 1.29 (jackknife s.e. 0.10). These results correspond to **Supplementary Figure 7**.

**Supplementary Table 9. Simulation results with the true liability-scale heritability set to 0.06 (instead of 0.13).** The first column for each sub-table lists the disease PheCodes that have the same true $h^2{}_{pleio}/h^2$ based on the specified genetic architecture. In the top two sub-tables, we show the estimated $h^2{}_{pleio}/h^2$, true $h^2{}_{pleio}/h^2$ and their absolute bias for 5 different values both before bias correction and after bias correction. Estimated $h^2{}_{pleio}/h^2$ before bias correction shows an upward bias, but modest downward bias for values of $h^2{}_{pleio}/h^2$ above 25% and modest upward bias for values below 5% after bias correction. These results correspond to **Supplementary Figure 8**. In the bottom two sub-tables, we show the sum of squared deviation vs. the sum of squared jackknife standard errors of $h^2{}_{pleio}/h^2$. Deviation is the difference between true $h^2{}_{pleio}/h^2$ and the average estimated $h^2{}_{pleio}/h^2$ across diseases that have the same true $h^2{}_{pleio}/h^2$. The estimated standard errors of $h^2{}_{pleio}/h^2$ before bias correction is anti-conservative as the sum of squared jackknife standard errors of $h^2{}_{pleio}/h^2$ is smaller than the sum of squared deviation across simulations. The estimated standard errors of $h^2{}_{pleio}/h^2$ after bias correction were approximately well-calibrated as the ratio of estimated standard errors to empirical standard deviations was equal to 1.02 (jackknife s.e. 0.08). These results correspond to **Supplementary Figure 9**.

**Supplementary Table 10. Simulation results with the prevalence set to 0.05 (instead of 0.1).** The first column for each sub-table lists the disease PheCodes that have the same true $h^2{}_{pleio}/h^2$ based on the specified genetic architecture. In the top two sub-tables, we show the estimated $h^2{}_{pleio}/h^2$, true $h^2{}_{pleio}/h^2$ and their absolute bias for 5 different values both before bias correction and after bias correction. Estimated $h^2{}_{pleio}/h^2$ before bias correction shows an upward bias, but it is approximately unbiased after bias correction. These results correspond to **Supplementary Figure 10**. In the bottom two sub-tables, we show the sum of squared deviation vs. the sum of squared jackknife standard errors of $h^2{}_{pleio}/h^2$. Deviation is the difference between true $h^2{}_{pleio}/h^2$ and the average estimated $h^2{}_{pleio}/h^2$ across diseases that have the same true $h^2{}_{pleio}/h^2$. The estimated standard errors of $h^2{}_{pleio}/h^2$ before bias correction is anti-conservative as the sum of squared jackknife standard errors of $h^2{}_{pleio}/h^2$ is smaller than the sum of squared deviation across simulations. The estimated standard errors of $h^2{}_{pleio}/h^2$ after bias correction were conservative as the ratio of estimated standard errors to empirical standard deviations was equal to 1.24 (jackknife s.e. 0.09). These results correspond to **Supplementary Figure 11**.

**Supplementary Table 11. Simulation results with the proportion of causal SNPs set to 1% (instead of 5%).** The first column for each sub-table lists the disease PheCodes that have the same true $h^2{}_{pleio}/h^2$ based on the specified genetic architecture. In the top two sub-tables, we show the estimated $h^2{}_{pleio}/h^2$, true $h^2{}_{pleio}/h^2$ and their absolute bias for 5 different values both before bias correction and after bias correction. Estimated $h^2{}_{pleio}/h^2$ before bias correction shows an upward bias, but it is approximately unbiased after bias correction. These results correspond to **Supplementary Figure 2**. In the bottom two sub-tables, we show the sum of squared deviation vs. the sum of squared jackknife standard errors of $h^2{}_{pleio}/h^2$. Deviation is the difference between true $h^2{}_{pleio}/h^2$ and the average estimated $h^2{}_{pleio}/h^2$ across diseases that have the same true $h^2{}_{pleio}/h^2$. The estimated standard errors of $h^2{}_{pleio}/h^2$ before bias correction is anti-conservative as the sum of squared jackknife standard errors of $h^2{}_{pleio}/h^2$ is smaller than the sum of squared deviation across simulations. The estimated standard errors of $h^2{}_{pleio}/h^2$ after bias correction were conservative as the ratio of estimated standard errors to empirical standard deviations was equal to 1.31 (jackknife s.e. 0.11). These results correspond to **Supplementary Figure 3**.

**Supplementary Table 12. Simulation results when $r_g$ within-disease categories equals to 0.5 and $r_g$ between-disease categories equals to 0.1, without pruning procedure.** The first column for each sub-table lists the disease PheCodes that have the same true $h^2{}_{pleio}/h^2$ based on the specified genetic architecture. In the top two sub-tables, we show the estimated $h^2{}_{pleio}/h^2$, true $h^2{}_{pleio}/h^2$ and their absolute bias for 5 different values without pruning procedure in both before bias correction and after bias correction. In the bottom two sub-tables, we show the sum of squared deviation vs. the sum of squared jackknife standard errors of $h^2{}_{pleio}/h^2$. Estimated $h^2{}_{pleio}/h^2$ without pruning both before and after bias correction are similar to results in **Supplementary Table 4** (results after pruning). These results correspond to **Supplementary Figure 12** and **13**.

**Supplementary Table 13. Simulations results of the reduction in $h^2{}_{pleio}/h^2$ and its standard error in analyses with one auxiliary disease category removed.** In each sub-table, we report the reduction of estimated $h^2{}_{pleio}/h^2$ , standard error, true reduction of $h^2{}_{pleio}/h^2$, the sum of squared jackknife standard error and the sum of deviation (the difference between true reduction of $h^2{}_{pleio}/h^2$ and the average estimated reduction of $h^2{}_{pleio}/h^2$ across diseases that have the same true reduction of $h^2{}_{pleio}/h^2$) when removing the 7 auxiliary categories and zero-trait-category for 15 diseases, respectively. These results correspond to **Supplementary Figure 14**.

**Supplementary Table 14. Genetic correlations between 15 UK Biobank diseases.** We report the genetic correlations and their corresponding categories. These results correspond to **Figure 3A**.

**Supplementary Table 15. Genetic correlations and liability correlations between 15 UK Biobank diseases.** We reported the genetic correlations, liability correlations, and their corresponding standard errors. We computed the z value using $\frac{r_g - r_l}{\sqrt{r_{g_{se}}^2 + r_{l_{se}}^2}}$, and then compute the right tailed p value. A Bonferroni-corrected significance threshold of $P < 4.76 \times$

$10^{-4}$(0.05/105) is applied. We also tested the $\frac{r_l - r_g}{\sqrt{{r_g}_{se}^2 + {r_l}_{se}^2}}$, and found that there is no pairs with liability correlation significantly larger than genetic correlation. These results correspond to **Figure 3**.

**Supplementary Table 16. Genetic correlations across all 15 UK Biobank diseases, 30 non-UK Biobank diseases, 17 UK Biobank quantitative traits.** These results correspond to **Figure 3A** and **Supplementary Figure 23**.

**Supplementary Table 17. $h^2{}_{pleio}/h^2$ before and after bias correction for all 15 UK Biobank diseases and the average with respect to 4 types of auxiliary diseases/traits.** D+Q: $h^2{}_{pleio}/h^2$ estimates with respect to 14 UK Biobank auxiliary diseases and 17 UK Biobank quantitative auxiliary traits. D: $h^2{}_{pleio}/h^2$ estimates with respect to 14 UK Biobank auxiliary diseases. D\target: $h^2{}_{pleio}/h^2$ estimates with respect to 14 UK Biobank auxiliary diseases excluding those from the same disease category as the target disease. D\(target+best other): $h^2{}_{pleio}/h^2$ estimates with respect to 14 UK Biobank auxiliary diseases excluding those from the same disease category as the target disease and from the next disease category whose removal had the greatest impact. We reported disease PheCode, pre-correction and post-correction $h^2{}_{pleio}/h^2$ and its standard error, scaling coefficient $\xi_c$ in bias correction, selected target and auxiliary diseases/traits. These results correspond to **Figure 4**, **Supplementary Figure 15** and **16**.

**Supplementary Table 18. Differences between auxiliary diseases choices for average estimates in Figure 4 and Figure 6.** In **Figure 4**, we compared average estimates from D+Q, D\target and D\(target+best other) to estimates from D. We reported the p value of these comparisons 1) using flat average; 2) using inverse-variance weighting average by weighting each disease (weightings were computed using the D group and applied to all four groups to facilitate comparison). In **Figure 6A** and **6B**, we compared average estimates from 30 non-UK Biobank auxiliary diseases, all 45 auxiliary diseases, and all 62 auxiliary diseases and quantitative traits to estimates from 15 UK Biobank auxiliary diseases. We also reported the p value of these comparisons 1) using flat average; 2) using inverse-variance weighting average by weighting each disease based on its $\frac{1}{s.e.^2}$ from the 15 UK Biobank auxiliary diseases group and applied them to all auxiliary trait choices.

**Supplementary Table 19. $\boldsymbol{h^2{}_{pleio}/h^2}$ estimates using the single auxiliary category and the reduction in $\boldsymbol{h^2{}_{pleio}/h^2}$ when one auxiliary disease category removed for all 15 UK Biobank target diseases.** In each sub-table, we report the $h^2{}_{pleio}/h^2$ estimates using the single auxiliary category, reduction of $h^2{}_{pleio}/h^2$ when one auxiliary disease category removed and their standard errors for 15 UK Biobank diseases with respect to all 7 auxiliary categories, respectively. $h^2{}_{pleio}/h^2$ of single-auxiliary-category estimate are zero when this auxiliary category only has the target disease. The reduction of $h^2{}_{pleio}/h^2$ when removing the auxiliary category are zero when pruning procedure has removed all diseases from that category. Therefore, further removal of this auxiliary category makes no difference. These results correspond to **Figure 5** and **Supplementary Figure 17**.

**Supplementary Table 20. Comparison of $r_g$ from BOLT-REML and $r_g$ from cross-trait LDSC with 4 options of constraining intercept.** We reported five estimates of $\mathbf{r_g}$ in following order: $r_g$ from BOLT-REML, $r_g$ from cross-trait LDSC without constraint on intercept, $r_g$ from cross-trait LDSC constraining heritability intercept, $r_g$ from cross-trait LDSC constraining genetic covariance intercept, $r_g$ from cross-trait LDSC constraining both intercepts. These results correspond to **Supplementary Figure 21**.

**Supplementary Table 21. Comparison of $\boldsymbol{h^2{}_{pleio}/h^2}$ estimated from BOLT-REML $r_g$ and $\boldsymbol{h^2{}_{pleio}/h^2}$ estimated from cross-trait LDSC $r_g$ with 4 options of constraining intercept.** We reported five estimates in following order: $h^2{}_{pleio}/h^2$ from BOLT-REML $r_g$, $h^2{}_{pleio}/h^2$ estimated from cross-trait LDSC $r_g$ without constraint on intercept, $h^2{}_{pleio}/h^2$ estimated from cross-trait LDSC $r_g$ constraining heritability intercept, $h^2{}_{pleio}/h^2$ estimated from cross-trait LDSC $r_g$ constraining genetic covariance intercept, $h^2{}_{pleio}/h^2$ estimated from cross-trait LDSC $r_g$ constraining both intercepts. These results correspond to **Supplementary Figure 22**.

**Supplementary Table 22. $h^2{}_{pleio}/h^2$ before and after bias correction for all 15 UK Biobank diseases and the average with respect to four types of auxiliary disease sets: 15 UK Biobank auxiliary diseases, 30 non-UK Biobank auxiliary diseases, all 45 auxiliary diseases, and all 62 auxiliary diseases and quantitative traits.** We reported disease PheCode, pre-correction and post-correction $h^2{}_{pleio}/h^2$ and its standard error, scaling coefficient $\xi_c$ in bias correction, selected target and auxiliary diseases/traits. These results correspond to **Supplementary Figure 24**.

**Supplementary Table 23. $h^2{}_{pleio}/h^2$ before and after bias correction for all 30 non-UK Biobank diseases and the average with respect to four types of auxiliary disease sets: 15 UK Biobank auxiliary diseases, 30 non-UK Biobank auxiliary diseases, all 45 auxiliary diseases and all 62 auxiliary diseases and quantitative traits.** We reported disease PheCode, pre-correction and post-correction $h^2{}_{pleio}/h^2$ and its standard error, scaling coefficient $\xi_c$ in bias correction, selected target and auxiliary diseases/traits. These results correspond to **Supplementary Figure 25**.

**Supplementary Table 24. $h^2{}_{pleio}/h^2$ estimates using the single auxiliary category and the reduction in $h^2{}_{pleio}/h^2$ when one auxiliary disease category removed for all 45 UK Biobank target diseases + 30 non-UK Biobank target diseases.** In each sub-table, we reported the $h^2{}_{pleio}/h^2$ estimates using the single auxiliary category, reduction of $h^2{}_{pleio}/h^2$ when one auxiliary disease category removed and their standard errors for 45 UK Biobank diseases with respect to all 11 auxiliary categories, respectively. $h^2{}_{pleio}/h^2$ of single-auxiliary-category estimate are zero when this auxiliary category only has the target disease. Reduction of $h^2{}_{pleio}/h^2$ when removing the auxiliary category are zero when pruning procedure has removed all diseases from that category. Therefore, further removal of this auxiliary category makes no difference. These results correspond to **Supplementary Figure 26**.

**Supplementary Table 25. Simulations results of $V^2{}_{pleio}/V^2$ and its standard error.** True $r_o$ is set to 0.5 within diseases categories and 0.1 between diseases categories for the 15 diseases (based on the 7 Phecode disease categories), which implies 4 different values of true $V^2{}_{pleio}/V^2$ for each target disease (ranging from 0.05 to 0.38). We simulated liabilities for 228,258 individuals. We computed true $r_l$ and true $V^2{}_{pleio}$ based on the simulated liabilities. Then, we used liability threshold model to generate binary phenotypes based on the empirical prevalence for these 15 UKB diseases. We applied our method on the simulated binary phenotypes to estimate $V^2{}_{pleio}$ and compared it to the true value. The first column for each sub-table lists the disease PheCodes that have the same true $V^2{}_{pleio}/V^2$ based on the specified correlation architecture. In the top sub-table, we show the estimated $V^2{}_{pleio}/V^2$, true $V^2{}_{pleio}/V^2$ and their absolute bias for 5 different values before bias correction. Estimated $V^2{}_{pleio}/V^2$ is unbiased without the need for bias correction. In the bottom sub-table, we show the sum of squared deviation vs. the sum of squared jackknife standard errors of $V^2{}_{pleio}/V^2$. Deviation is the difference between true $V^2{}_{pleio}/V^2$ and the average estimated $V^2{}_{pleio}/V^2$ across diseases that have the same true $V^2{}_{pleio}/V^2$. We estimate the standard error by jackknifing over blocks of individuals across all diseases. The estimated standard errors of $V^2{}_{pleio}/V^2$ are anti-conservative, as the ratio of the average estimated squared jackknife standard error for $V^2{}_{pleio}/V^2$ to the average squared deviation across simulations was equal to 0.65 (jackknife s.e. 0.04); we determined that this does not impact our results, as the standard errors of $V^2{}_{pleio}/V^2$ estimates are small given the large sample size of UK Biobank data. These results correspond to **Supplementary Figure 27**.

**Supplementary Table 26. Comparing $\boldsymbol{h^2_{pleio}/h^2}$ $\boldsymbol{vs.V^2_{pleio}/V^2}$ $\boldsymbol{vs.E^2_{pleio}/E^2}$.** In the top sub-table, we report the post-correction $h^2_{pleio}/h^2$, pre-correction $V^2_{pleio}/V^2$, standard errors, z score for comparing their difference and two-tailed p value. A Bonferroni-corrected significance threshold of $P < 3.33 \times 10^{-3}$(0.05/15) is applied. In the bottom sub-table, we report the post-correction $h^2_{pleio}$, pre-correction $V^2_{pleio}/V^2$, $E^2_{pleio}/E^2$, and their standard errors. These results correspond to **Figure 7** and **Supplementary Figures 28-31**.

**Supplementary Table 27. The constrained intercepts for genetic covariance for each disease pair**. Computation of constrained intercepts for genetic covariance are described in **Methods** section.

**Supplementary Table 28. Comparison of $\boldsymbol{h^2_{pleio}/h^2}$ between estimate using only EA and reduction when removing the EA.** To investigate the impact of educational attainment (EA) on pleiotropy, we performed two analyses. First, we estimated $h^2_{pleio}/h^2$ with respect to EA as the only auxiliary trait for each of the 15 UK Biobank diseases and determined that EA had a substantial contribution, with an average of 9.9% (s.e. 0.8%). Second, we assessed the impact of removing EA from the set of auxiliary traits by estimating the difference between (i) $h^2_{pleio}/h^2$ with respect to 15 UK Biobank auxiliary diseases + 17 UK Biobank auxiliary quantitative traits vs. (ii) $h^2_{pleio}/h^2$ with respect to 15 UK Biobank auxiliary diseases + 16 UK Biobank quantitative traits excluding EA. We determined that differences were small, with an average reduction of 0.87% (s.e. 0.39%). These results correspond to **Supplementary Figures 18**.

**Supplementary Table 29. Changes of $\boldsymbol{h^2_{pleio}/h^2}$ across 15 UK Biobank diseases for various $\boldsymbol{r_g}^2$ thresholds between target and auxiliary diseases.** We performed analyses to test the $h^2_{pleio}/h^2$ for 15 UK Biobank diseases and the average for all $r_g{}^2$ thresholds between target and auxiliary diseases ranging from 0.1 to 0.8 in increments of 0.05. These results correspond to **Supplementary Figures 20**.

**Supplementary Table 30. Comparison between $\boldsymbol{h^2_{pleio}/h^2}$ with the sum of (bias-corrected) $\boldsymbol{r_g}^2$ across auxiliary diseases, for different target diseases and auxiliary disease sets.** For each of 15 UK Biobank target diseases, we considered 14 auxiliary disease sets (containing 1, …, 14 auxiliary diseases), defined by starting with the auxiliary disease with highest $r_g{}^2$ with the target disease and iteratively adding auxiliary diseases in order of decreasing $r_g{}^2$ with the target disease. We note that for some target disease, pruning will happen when adding auxiliary diseases one by one, so that the number of the final auxiliary disease set will decrease.

**Supplementary Table 31. Correlation between $\boldsymbol{h^2_{pleio}/h^2}$ and the sum of (bias-corrected and without-bias-corrected) $\boldsymbol{r_g}^2$ across auxiliary diseases, across the 14 auxiliary disease sets.** The average correlation (across 15 target diseases) was 0.28 (0.30 when not applying bias correction to ${r_g}^2$), implying that ${h^2}_{pleio}/h^2$ captures different information than the sum of (bias-corrected) ${r_g}^2$. These results correspond to **Supplementary Tables 30**.

### Supplementary Figures

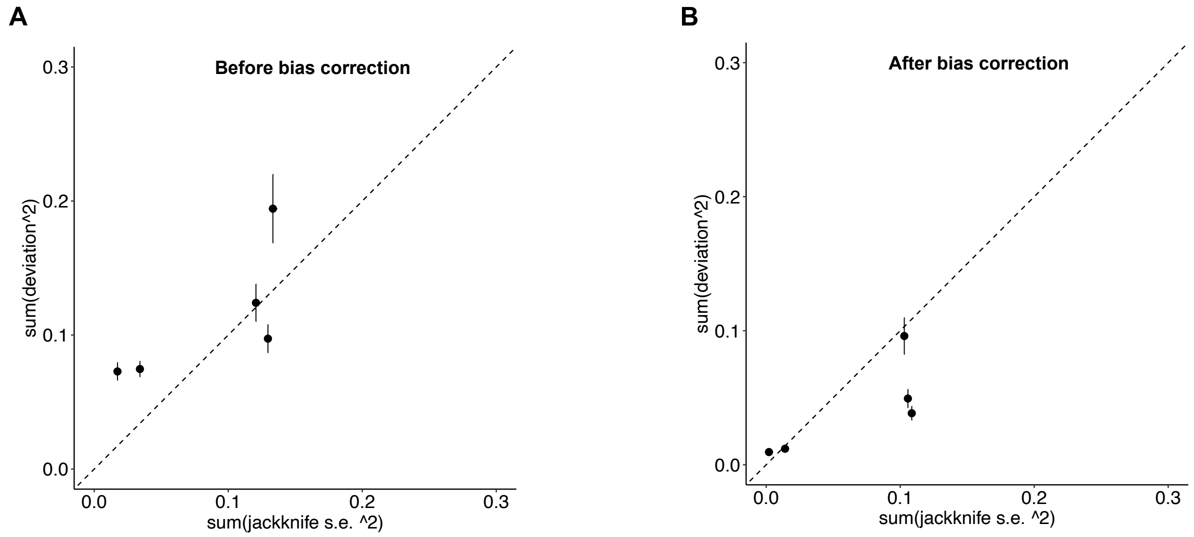


**Supplementary Figure 1. Estimated standard errors of $h^2_{pleio}/h^2$ from PHBC were conservative**. Deviation is the difference between true $h^2_{pleio}/h^2$ and the average estimated $h^2_{pleio}/h^2$ . We average across simulated diseases that have the same true $h^2_{pleio}/h^2$ as in **Figure 2**. Five dots correspond to the 5 different values of true $h^2_{pleio}/h^2$. (A) The estimated standard errors of $h^2_{pleio}/h^2$ before bias correction is anti-conservative as the sum of squared jackknife standard errors of $h^2_{pleio}/h^2$ is smaller than the sum of squared deviation across simulations. (B) The estimated standard errors of $h^2_{pleio}/h^2$ after bias correction were **conservative** as the ratio of estimated standard errors to empirical standard deviations was equal to 1.63 (jackknife s.e. 0.14). Detailed results are provided in **Supplementary Table 4**.

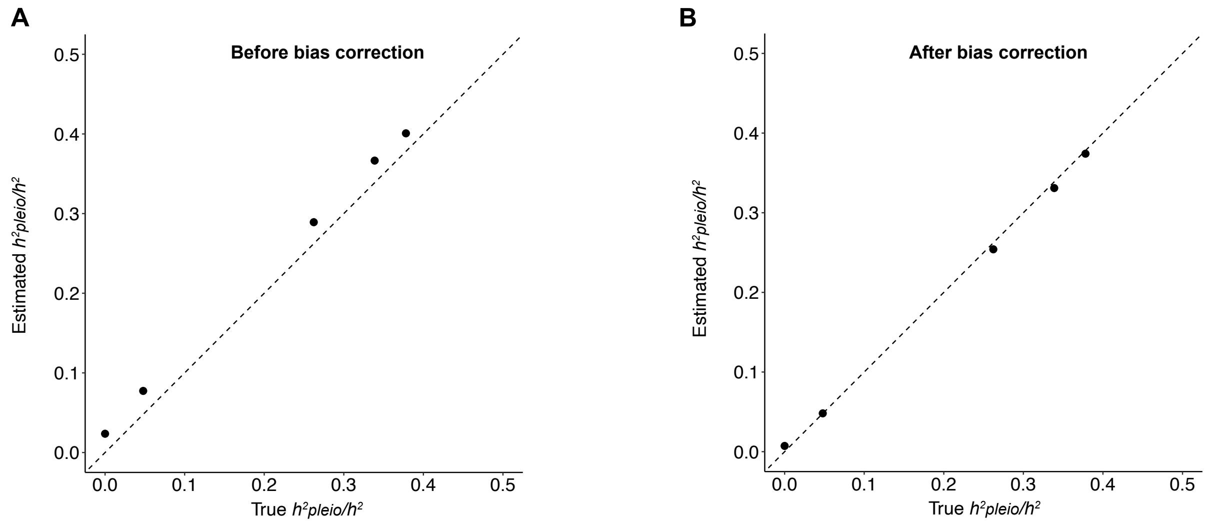


**Supplementary Figure 2. PHBC corrects the upwards bias in simulations with the proportion of causal SNPs set to 1% (instead of 5%).** Estimated $h^2{}_{pleio}/h^2$ is approximately unbiased compared to the simulated truth, which is similar to **Figure 2**. (A) Estimated $h^2{}_{pleio}/h^2$ without the bias correction step shows an upward bias. (B) Estimated $h^2{}_{pleio}/h^2$ is approximated unbiased after the Monte-Carlo bias correction. Each dot and error bar represents the mean and standard error of diseases that has the same true $h^2{}_{pleio}/h^2$ across simulations, in which error bars are smaller than dot size in some cases. Detailed results are reported in **Supplementary Table 11**.

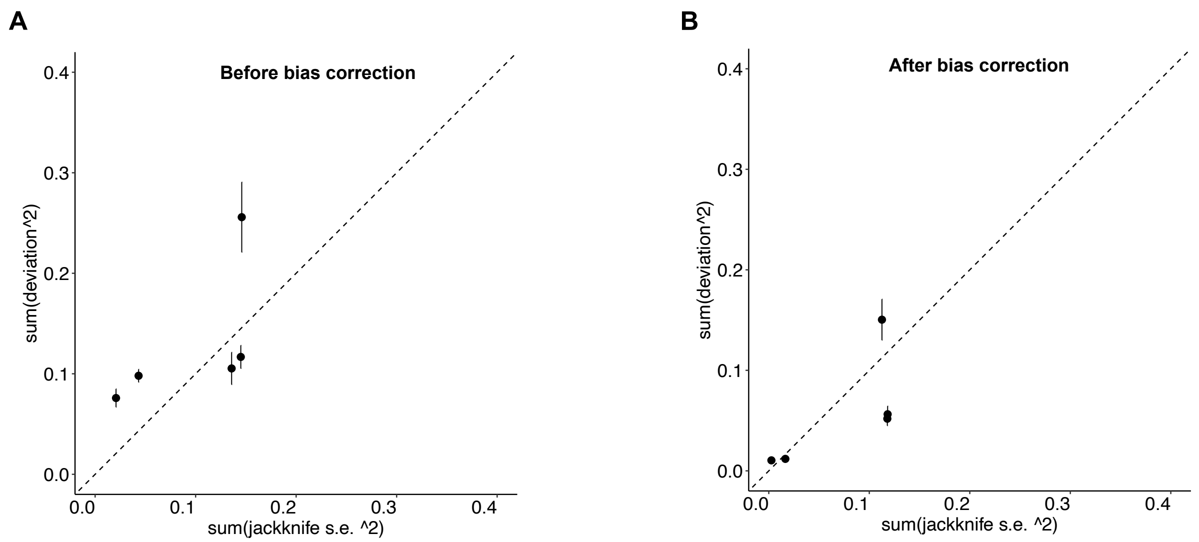


**Supplementary Figure 3. Estimated standard errors of $h^2{}_{pleio}/h^2$ from PHBC were conservative in simulations with the proportion of causal SNPs set to 1% (instead of 5%).** Deviation is the difference between true $h^2{}_{pleio}/h^2$ and the average estimated $h^2{}_{pleio}/h^2$ across diseases that have the same true $h^2{}_{pleio}/h^2$ in **Figure 2**. Five dots correspond to the 5 different values of true $h^2{}_{pleio}/h^2$. (A) The estimated standard errors of $h^2{}_{pleio}/h^2$ before bias correction is anti-conservative showing that the sum of squared jackknife standard errors of $h^2{}_{pleio}/h^2$ is smaller than the sum of squared deviation across simulations. (B) The estimated standard errors of $h^2{}_{pleio}/h^2$ after bias correction were conservative as the ratio of the average estimated squared jackknife standard error to the average empirical squared deviation equal to 1.31 (jackknife s.e. 0.11). Detailed results are provided in **Supplementary Table 11**.

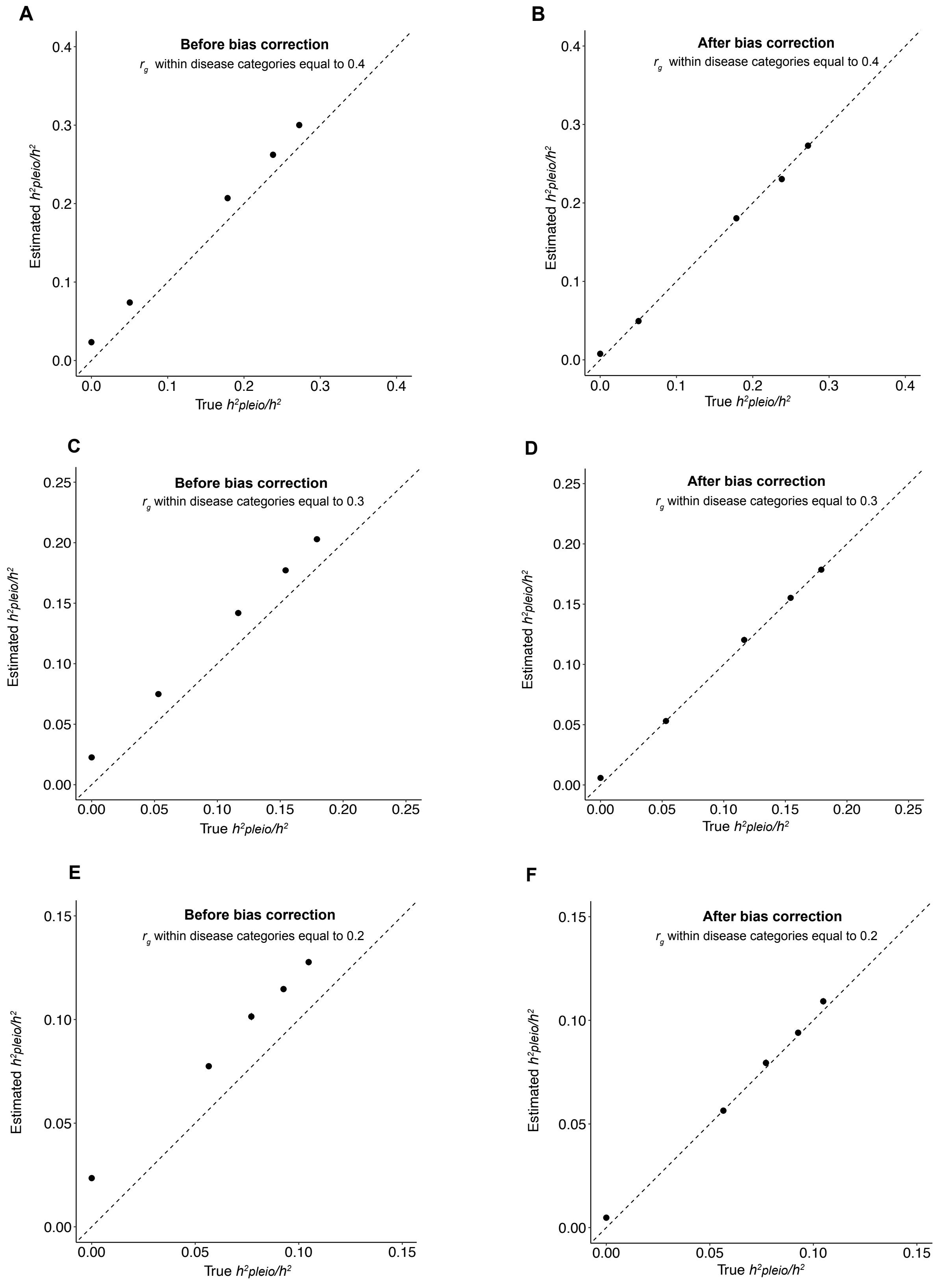

A
Before bias correction
$r_g$ within disease categories equal to 0.4
B
After bias correction
$r_g$ within disease categories equal to 0.4
C
Before bias correction
$r_g$ within disease categories equal to 0.3
D
After bias correction
$r_g$ within disease categories equal to 0.3
E
Before bias correction
$r_g$ within disease categories equal to 0.2
F
After bias correction
$r_g$ within disease categories equal to 0.2
Estimated $h^2pleio/h^2$
True $h^2pleio/h^2$
0.0
0.1
0.2
0.3
0.4
0.00
0.05
0.10
0.15
0.20
0.25

**Supplementary Figure 4. Simulations with $r_g$ within-disease categories equal to 0.4, 0.3, 0.2 (and $r_g$ between-disease categories still equal to 0.1).** (A, C, E) In simulation with $r_g$ within-disease categories equal to 0.4, 0.3, 0.2, estimated ${h^2}_{pleio}/h^2$ without the bias correction step shows an upward bias. (B, D, F) In simulation with $r_g$ within-disease categories equal to 0.4, 0.3, 0.2, we observed that approximately unbiased results after Monte-Carlo bias correction. Each dot and error bar in each panel represents the mean and standard error of diseases that has the same true ${h^2}_{pleio}/h^2$ across simulations, in which error bars are smaller than dot size in some cases. Detailed results are reported in **Supplementary Tables 5-7**.

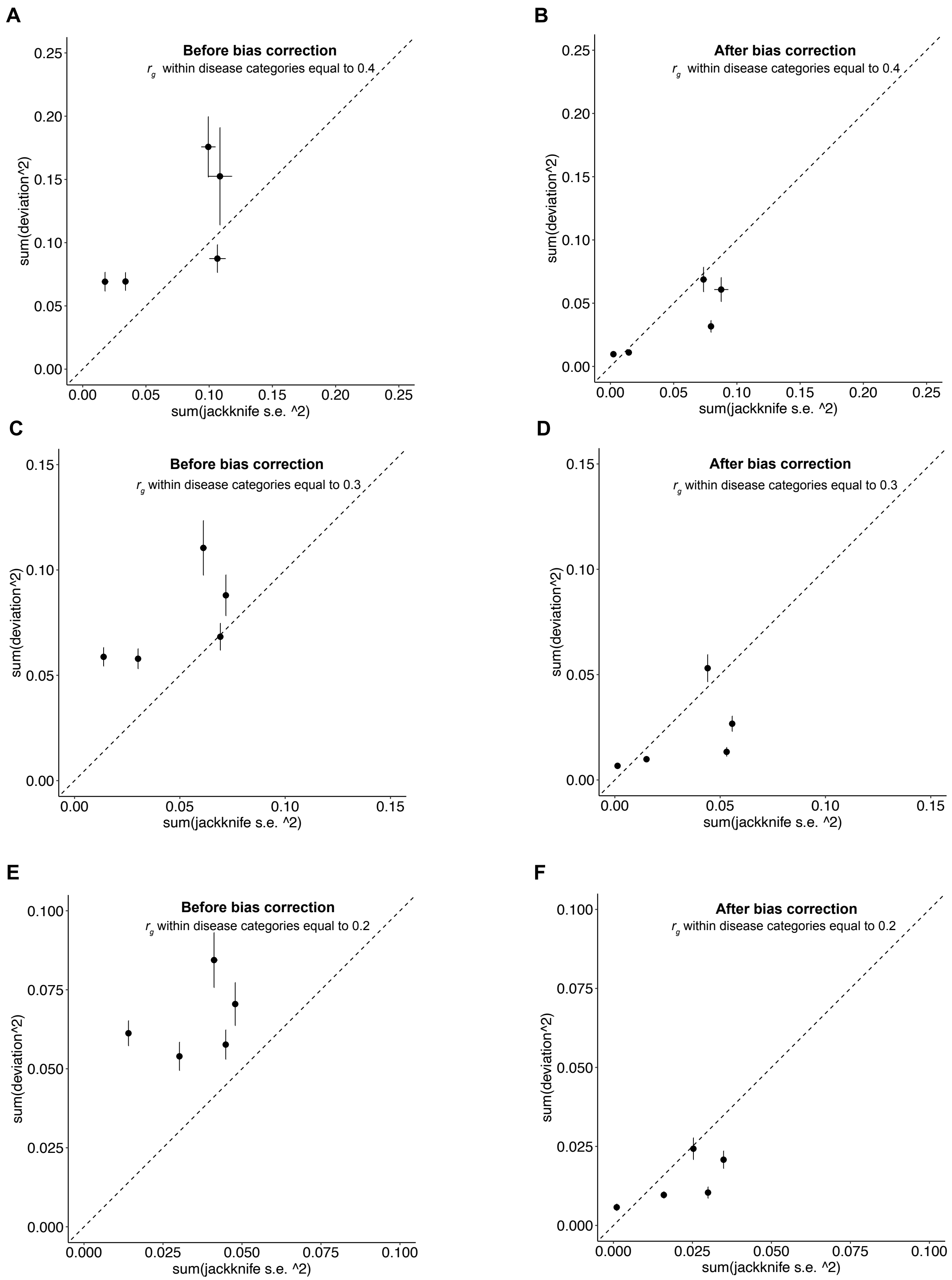


**Supplementary Figure 5. Estimated standard errors of $h^2{}_{pleio}/h^2$ from PHBC were conservative in simulations with $r_g$ within-disease categories equal to 0.4, 0.3, 0.2 (and $r_g$**

**between-disease categories still equal to 0.1).** Deviation is the difference between true $h^2{}_{pleio}/h^2$ and the average estimated $h^2{}_{pleio}/h^2$ across diseases that have the same true $h^2{}_{pleio}/h^2$. Five dots correspond to the 5 different values of true $h^2{}_{pleio}/h^2$. (A, C, E) In simulation with $r_g$ within-disease categories equal to 0.4, 0.3, 0.2, the estimated standard errors of $h^2{}_{pleio}/h^2$ before bias correction is anti-conservative as the sum of squared jackknife standard errors of $h^2{}_{pleio}/h^2$ is smaller than the sum of squared deviation across simulations. (B, D, F) In simulation with $r_g$ within-disease categories equal to 0.4, 0.3, 0.2, the estimated standard errors of $h^2{}_{pleio}/h^2$ after bias correction were conservative the ratio of the average estimated squared jackknife standard error to the average empirical squared deviation equal to 1.42 (jackknife s.e. 0.10), 1.54 (jackknife s.e. 0.11) and 1.51 (jackknife s.e. 0.09). Detailed results are provided in **Supplementary Tables 5-7**.

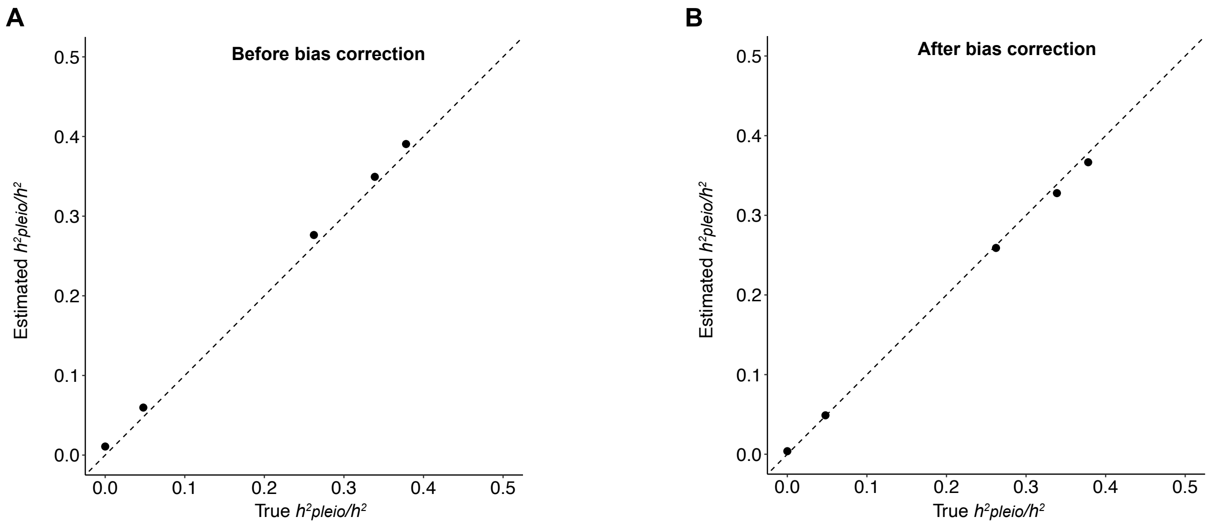


**Supplementary Figure 6. PHBC corrects the upwards bias in simulations with the true liability-scale heritability set to 0.25 (instead of 0.13).** Estimated $h^2{}_{pleio}/h^2$ is approximately unbiased compared to the simulated truth, which is similar to **Figure 2**. (A) Estimated $h^2{}_{pleio}/h^2$ without the bias correction step shows an upward bias. (B) Estimated $h^2{}_{pleio}/h^2$ is approximated unbiased after the Monte-Carlo bias correction. Each dot and error bar represents the mean and standard error of diseases that has the same true $h^2{}_{pleio}/h^2$ across simulations, in which error bars are smaller than dot size in some cases. Detailed results are reported in **Supplementary Table 8**.

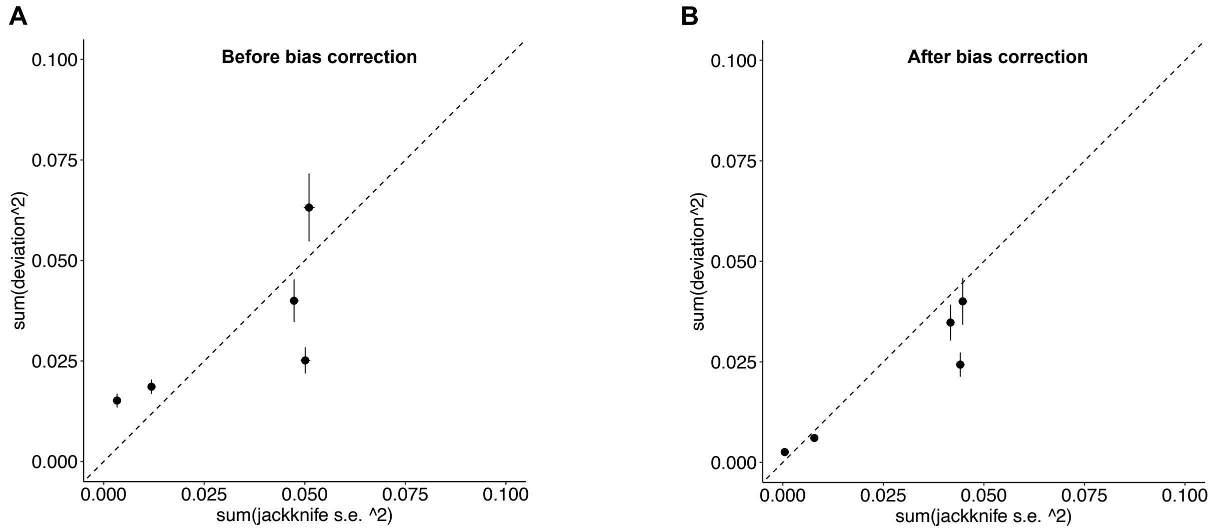


**Supplementary Figure 7. Estimated standard errors of $\boldsymbol{h^2}_{\boldsymbol{pleio}}/\boldsymbol{h^2}$ from PHBC were conservative in simulations with the true liability-scale heritability set to 0.25 (instead of 0.13).** Deviation is the difference between true $h^2{}_{pleio}/h^2$ and the average estimated $h^2{}_{pleio}/h^2$ across diseases that have the same true $h^2{}_{pleio}/h^2$ in **Figure 2**. Five dots correspond to the 5 different values of true $h^2{}_{pleio}/h^2$. (A) The estimated standard errors of $h^2{}_{pleio}/h^2$ before bias correction is anti-conservative showing that the sum of squared jackknife standard errors of $h^2{}_{pleio}/h^2$ is smaller than the sum of squared deviation across simulations. (B) The estimated standard errors of $h^2{}_{pleio}/h^2$ after bias correction were conservative as the ratio of the average estimated squared jackknife standard error to the average empirical squared deviation equal to 1.29 (jackknife s.e. 0.10). Detailed results are provided in **Supplementary Table 8**.

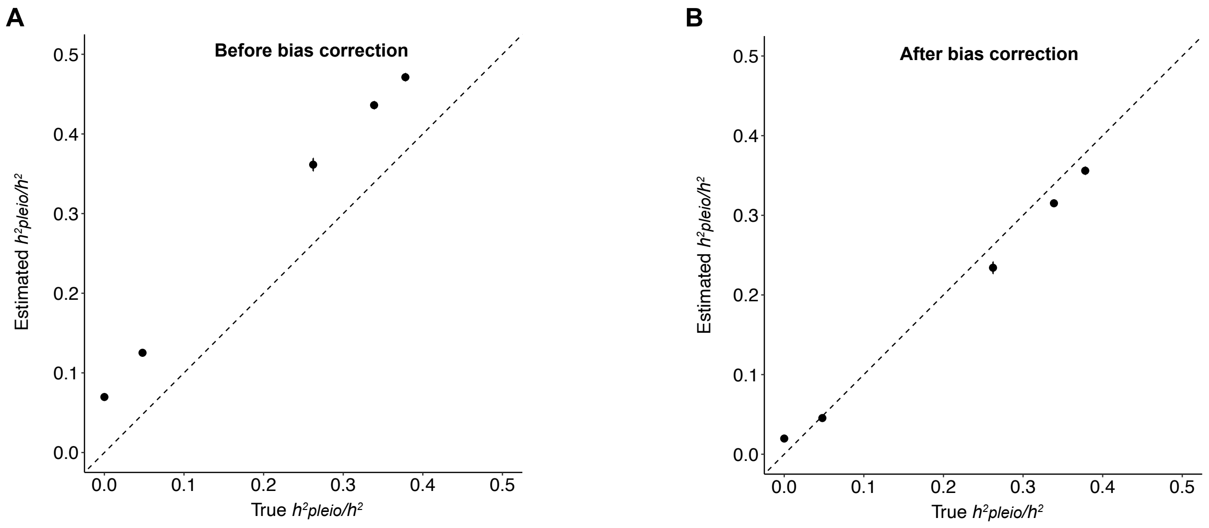


**Supplementary Figure 8. PHBC corrects the upwards bias in simulations with the true liability-scale heritability set to 0.06 (instead of 0.13).** Estimated $h^2{}_{pleio}/h^2$ is approximately unbiased compared to the simulated truth, which is similar to **Figure 2**. (A) Estimated $h^2{}_{pleio}/h^2$ without the bias correction step shows an upward bias. (B) Estimated $h^2{}_{pleio}/h^2$ show modest downward bias for values above 25% and modest upward bias for values below 5% after the Monte-Carlo bias correction. Each dot and error bar represents the mean and standard error of diseases that has the same true $h^2{}_{pleio}/h^2$ across simulations, in which error bars are smaller than dot size in some cases. The average simulated heritability z-score (average of 11.3 across simulated diseases) is similar to the average of 10.2 across 15 UK Biobank diseases. Detailed results are reported in **Supplementary Table 9**.

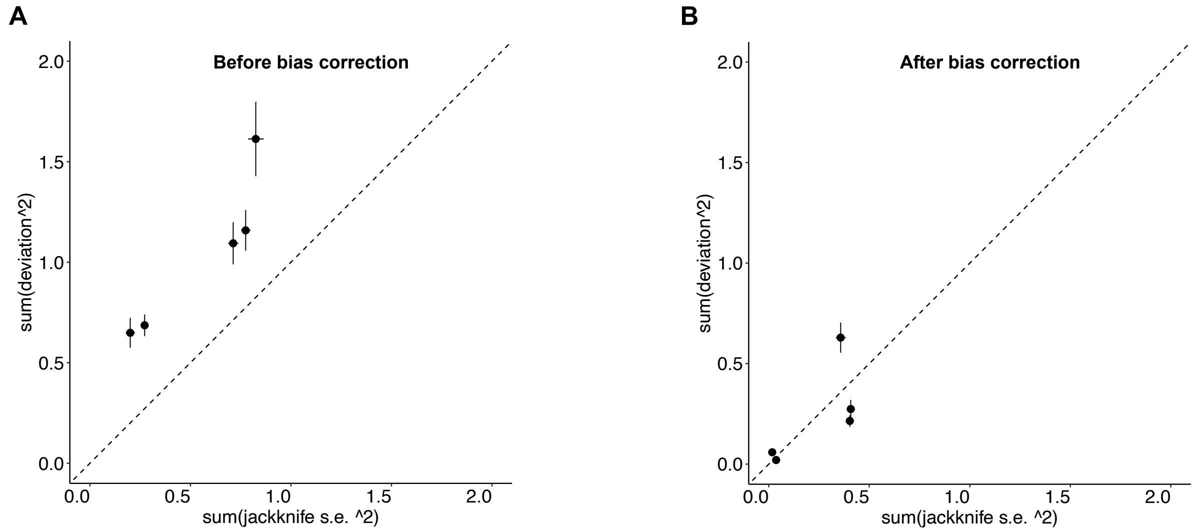


**Supplementary Figure 9. Estimated standard errors of $h^2{}_{pleio}/h^2$ from PHBC were approximately well-calibrated in simulations with the true liability-scale heritability set to 0.06 (instead of 0.13).** Deviation is the difference between true $h^2{}_{pleio}/h^2$ and the average estimated $h^2{}_{pleio}/h^2$ across diseases that have the same true $h^2{}_{pleio}/h^2$ in **Figure 2**. Five dots correspond to the 5 different values of true $h^2{}_{pleio}/h^2$. (A) The estimated standard errors of $h^2{}_{pleio}/h^2$ before bias correction is anti-conservative showing that the sum of squared jackknife standard errors of $h^2{}_{pleio}/h^2$ is smaller than the sum of squared deviation across simulations. (B) The estimated standard errors of $h^2{}_{pleio}/h^2$ after bias correction were approximately well-calibrated as the ratio of the average estimated squared jackknife standard error to the average empirical squared deviation equal to 1.02 (jackknife s.e. 0.08). Detailed results are provided in **Supplementary Table 9**.

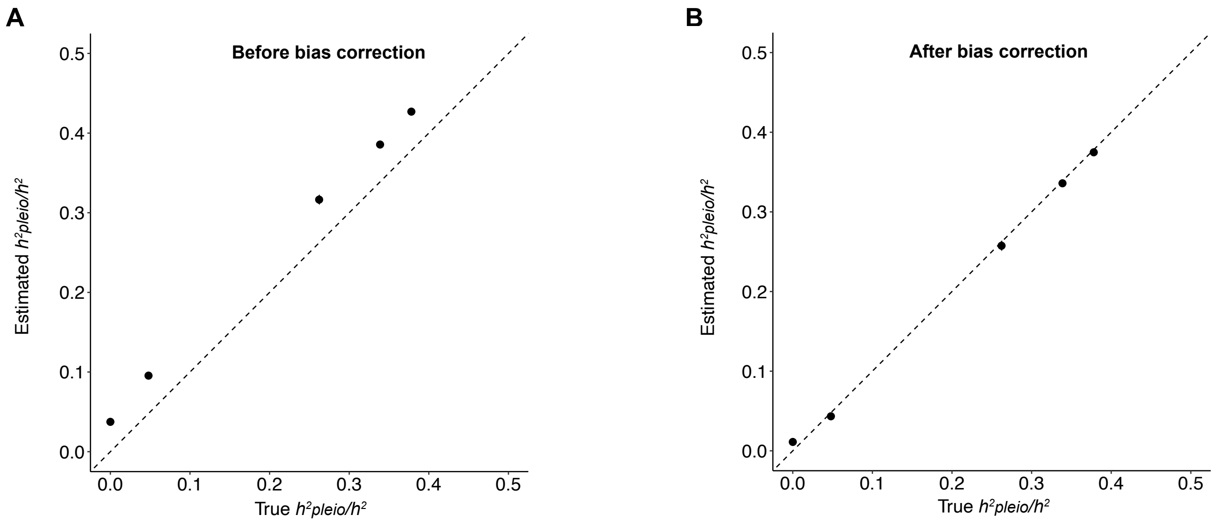


**Supplementary Figure 10. PHBC corrects the upwards bias in simulations with the prevalence set to 0.05 (instead of 0.1).** Estimated $h^2{}_{pleio}/h^2$ is approximately unbiased compared to the simulated truth, which is similar to **Figure 2**. (A) Estimated $h^2{}_{pleio}/h^2$ without the bias correction step shows an upward bias. (B) Estimated $h^2{}_{pleio}/h^2$ is approximated unbiased after the Monte-Carlo bias correction. The average simulated heritability z-score (average of 9.6 across simulated diseases) is similar to the average of 10.2 across 15 UK Biobank diseases. Each dot and error bar represents the mean and standard error of diseases that has the same true $h^2{}_{pleio}/h^2$ across simulations, in which error bars are smaller than dot size in some cases. Detailed results are reported in **Supplementary Table 10**.

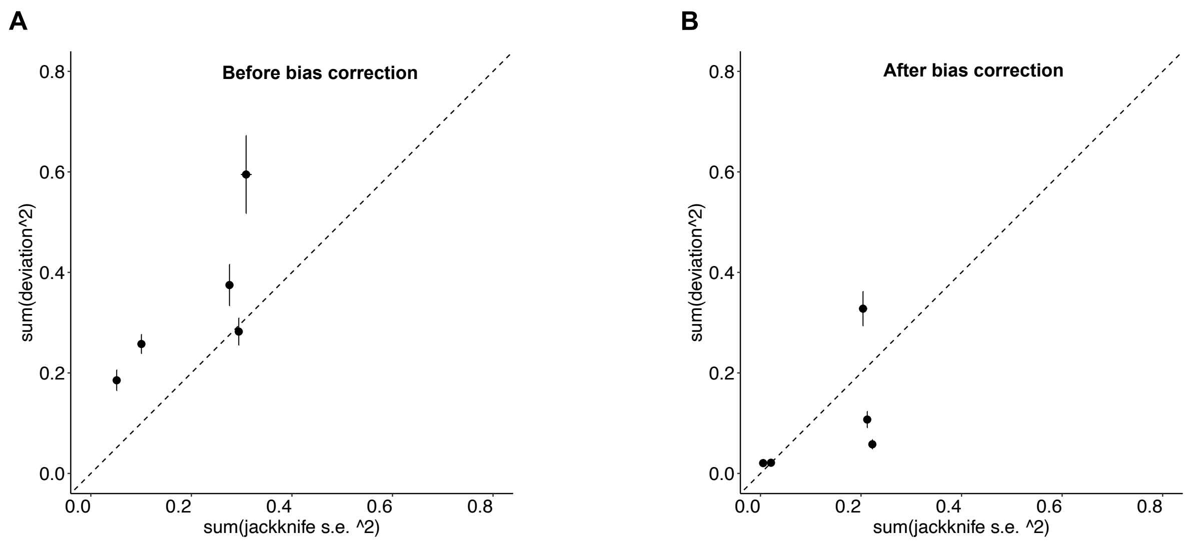


**Supplementary Figure 11. Estimated standard errors of $h^2{}_{pleio}/h^2$ from PHBC were conservative in simulations with the prevalence set to 0.05 (instead of 0.1).** Deviation is the difference between true $h^2{}_{pleio}/h^2$ and the average estimated $h^2{}_{pleio}/h^2$ across diseases that have the same true $h^2{}_{pleio}/h^2$ in **Figure 2**. Five dots correspond to the 5 different values of true $h^2{}_{pleio}/h^2$. (A) The estimated standard errors of $h^2{}_{pleio}/h^2$ before bias correction is anti-conservative showing that the sum of squared jackknife standard errors of $h^2{}_{pleio}/h^2$ is smaller than the sum of squared deviation across simulations. (B) The estimated standard errors of $h^2{}_{pleio}/h^2$ after bias correction were conservative as the ratio of the average estimated squared jackknife standard error to the average empirical squared deviation equal to 1.24 (jackknife s.e. 0.09). Detailed results are provided in **Supplementary Table 10**.

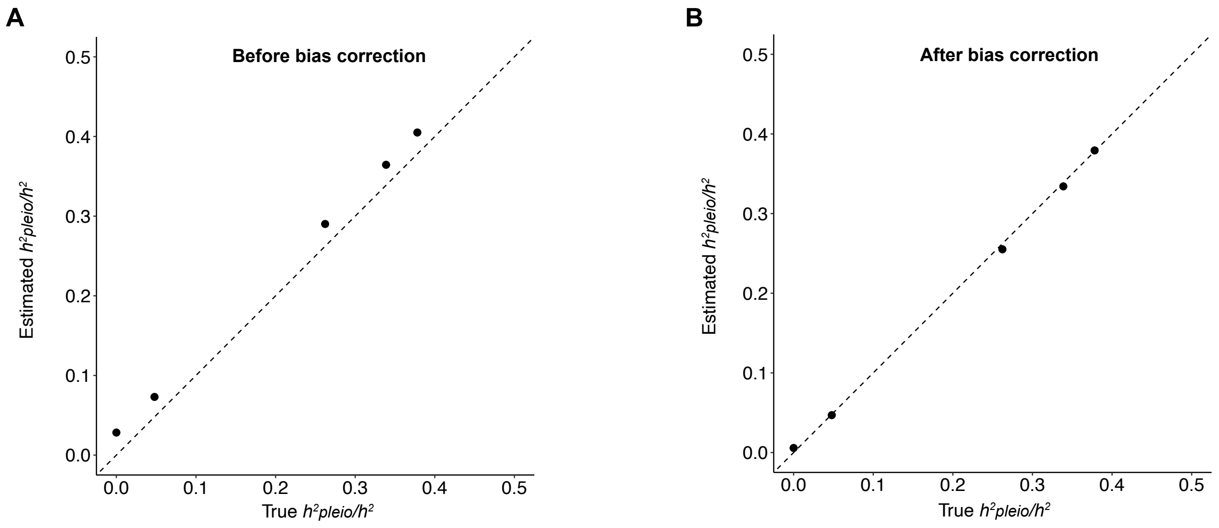


**Supplementary Figure 12. PHBC corrects the upwards bias in simulations without pruning procedure.** Estimated ${h^2}_{pleio}/h^2$ is approximately unbiased compared to the simulated truth, which is similar to **Figure 2**. (A) Estimated ${h^2}_{pleio}/h^2$ without the bias correction step shows an upward bias. (B) Estimated ${h^2}_{pleio}/h^2$ is approximated unbiased after the Monte-Carlo bias correction. Each dot and error bar represents the mean and standard error of diseases that has the same true ${h^2}_{pleio}/h^2$ across simulations, in which error bars are smaller than dot size in some cases. Detailed results are reported in **Supplementary Table 12**.

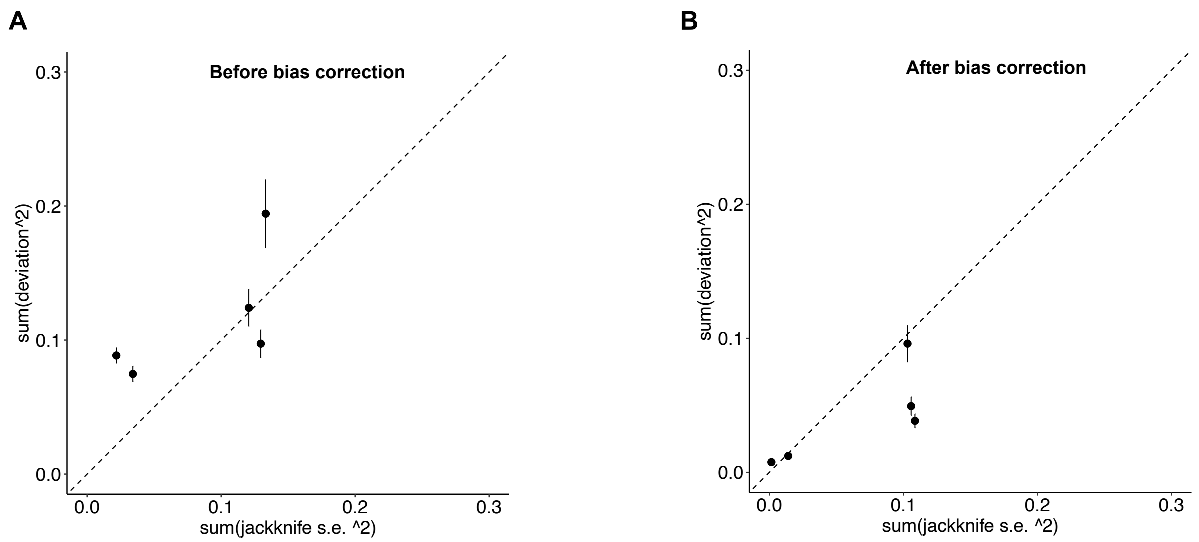


**Supplementary Figure 13. Estimated standard errors of $h^2{}_{pleio}/h^2$ from PHBC were conservative in simulations without pruning procedure**. Deviation is the difference between true $h^2{}_{pleio}/h^2$ and the average estimated $h^2{}_{pleio}/h^2$ across diseases that have the same true $h^2{}_{pleio}/h^2$ in **Figure 2**. Five dots correspond to the 5 different values of true $h^2{}_{pleio}/h^2$. (A) The estimated standard errors of $h^2{}_{pleio}/h^2$ before bias correction is anti-conservative showing that the sum of squared jackknife standard errors of $h^2{}_{pleio}/h^2$ is smaller than the sum of squared deviation across simulations. (B) The estimated standard errors of $h^2{}_{pleio}/h^2$ after bias correction were conservative as the ratio of the average estimated squared jackknife standard error to the average empirical squared deviation equal to 1.63 (jackknife s.e. 0.14). Detailed results are provided in **Supplementary Table 12**.

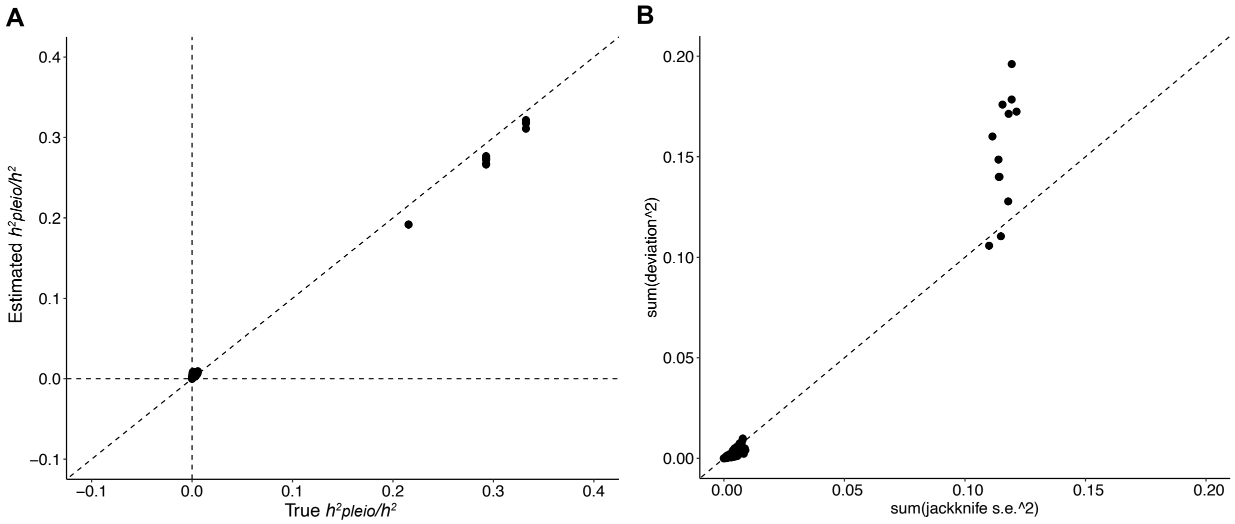


**Supplementary Figure 14. Simulations of the reduction in $h^2{}_{pleio}/h^2$ and its standard error in analyses with one auxiliary disease category removed.** (A) To remove the impact of randomness in Monte-Carlo bias correction when computing the reduction, we first computed the difference of $h^2{}_{pleio}/h^2$ w.r.t. all auxiliary diseases vs. $h^2{}_{pleio}/h^2$ w.r.t. auxiliary diseases excluding a PheCode category before bias correction. We multiplied the difference by the scaling coefficient $\xi_c^2$ of $h^2{}_{pleio}/h^2$ w.r.t. all auxiliary diseases, which represented the contribution of an auxiliary PheCode category to the $h^2{}_{pleio}/h^2$ w.r.t. all auxiliary diseases. The estimated reduction of $h^2{}_{pleio}/h^2$ will have modest downward bias when true reduction is above 20% but noted that estimates in empirical data are usually very small (less than 10% in **Supplementary Tables 19** and **24**). (B) Deviation is the difference between true reduction of $h^2{}_{pleio}/h^2$ when removing auxiliary category and the average estimated reduction of $h^2{}_{pleio}/h^2$ across diseases that have the same true reduction of $h^2{}_{pleio}/h^2$. We reported the jackknife standard error of the before-bias-correction difference of $h^2{}_{pleio}/h^2$ as the standard error of the reduction, which is a conservative standard error shown that most dots have the sum of squared standard errors matching the sum of squared deviation across simulations. Detailed results are reported in **Supplementary Table 13**.

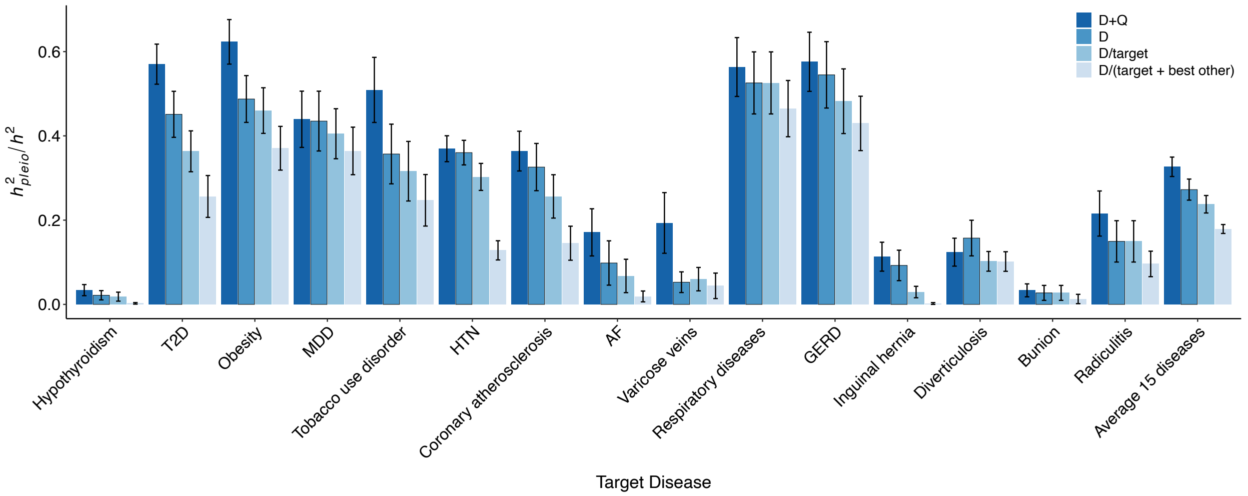


**Supplementary Figure 15.** $h^2{}_{pleio}/h^2$ **after bias correction for all 15 UK Biobank diseases and the average.** D+Q: $h^2{}_{pleio}/h^2$ estimates with respect to 14 UK Biobank auxiliary diseases and 17 UK Biobank quantitative auxiliary traits. D: $h^2{}_{pleio}/h^2$ estimates with respect to 14 UK Biobank auxiliary diseases. D\target: $h^2{}_{pleio}/h^2$ estimates with respect to 14 UK Biobank auxiliary diseases excluding those from the same disease category as the target disease. D\(target+best other): $h^2{}_{pleio}/h^2$ estimates with respect to 14 UK Biobank auxiliary diseases excluding those from the same disease category as the target disease and from the next disease category whose removal had the greatest impact. Error bars represent jackknife standard errors. Detailed results are provided in **Supplementary Table 17**. Abbreviation: T2D: type 2 diabetes; MDD: depression; HTN: hypertension.

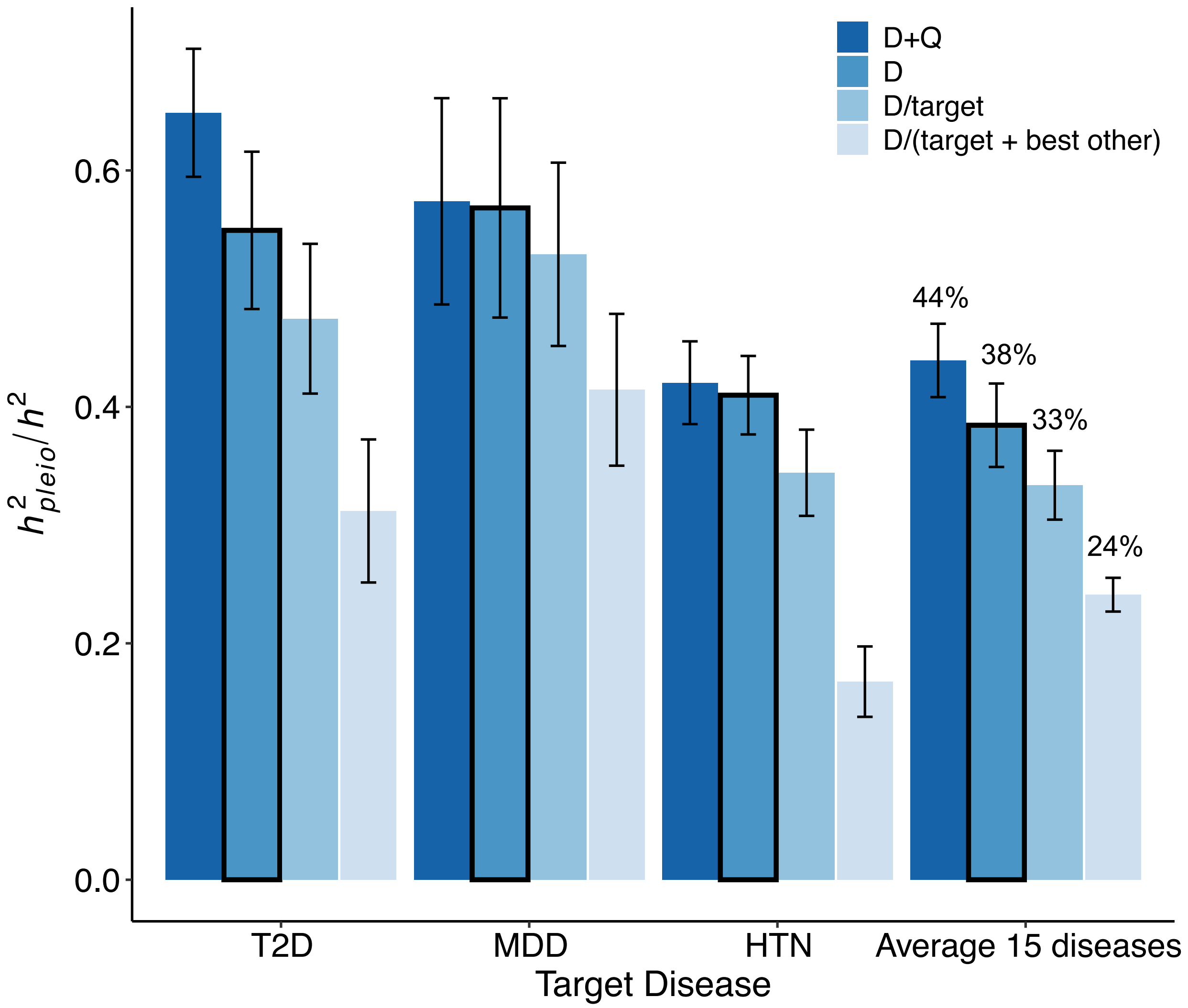


**Supplementary Figure 16.** $h^2{}_{pleio}/h^2$ **before bias correction for three representative diseases and the average across 15 UK Biobank diseases.** Comparing to **Figure 4**, we observed an upwards bias for $h^2{}_{pleio}/h^2$ before bias correction. D+Q: $h^2{}_{pleio}/h^2$ estimates with respect to 14 UK Biobank auxiliary diseases and 17 UK Biobank quantitative auxiliary traits. D: $h^2{}_{pleio}/h^2$ estimates with respect to 14 UK Biobank auxiliary diseases. D\target: $h^2{}_{pleio}/h^2$ estimates with respect to 14 UK Biobank auxiliary diseases excluding those from the same disease category as the target disease. D\(target+best other): $h^2{}_{pleio}/h^2$ estimates with respect to 14 UK Biobank auxiliary diseases excluding those from the same disease category as the target disease and from the next disease category whose removal had the greatest impact. Error bars represent jackknife standard errors. Detailed results are provided in **Supplementary Table 17**. Abbreviation: T2D: type 2 diabetes; MDD: depression; HTN: hypertension.

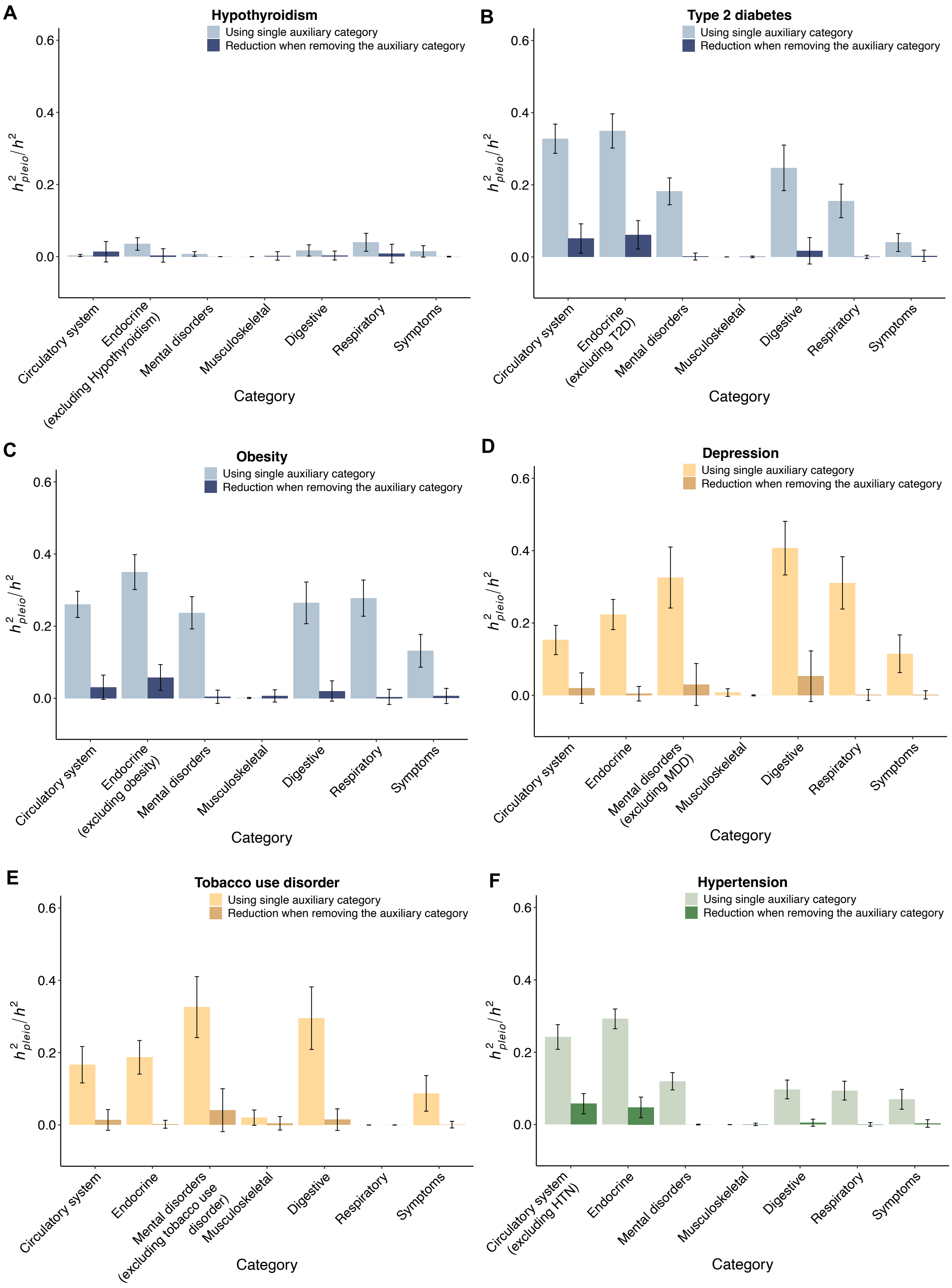
A
Hypothyroidism
Using single auxiliary category
Reduction when removing the auxiliary category
$h^2_{pleio}/h^2$
Circulatory system
Endocrine (excluding Hypothyroidism)
Mental disorders
Musculoskeletal
Digestive
Respiratory
Symptoms
Category
B
Type 2 diabetes
Using single auxiliary category
Reduction when removing the auxiliary category
Endocrine (excluding T2D)
C
Obesity
Using single auxiliary category
Reduction when removing the auxiliary category
Endocrine (excluding obesity)
D
Depression
Using single auxiliary category
Reduction when removing the auxiliary category
Mental disorders (excluding MDD)
E
Tobacco use disorder
Using single auxiliary category
Reduction when removing the auxiliary category
Mental disorders (excluding tobacco use disorder)
F
Hypertension
Using single auxiliary category
Reduction when removing the auxiliary category
Circulatory system (excluding HTN)

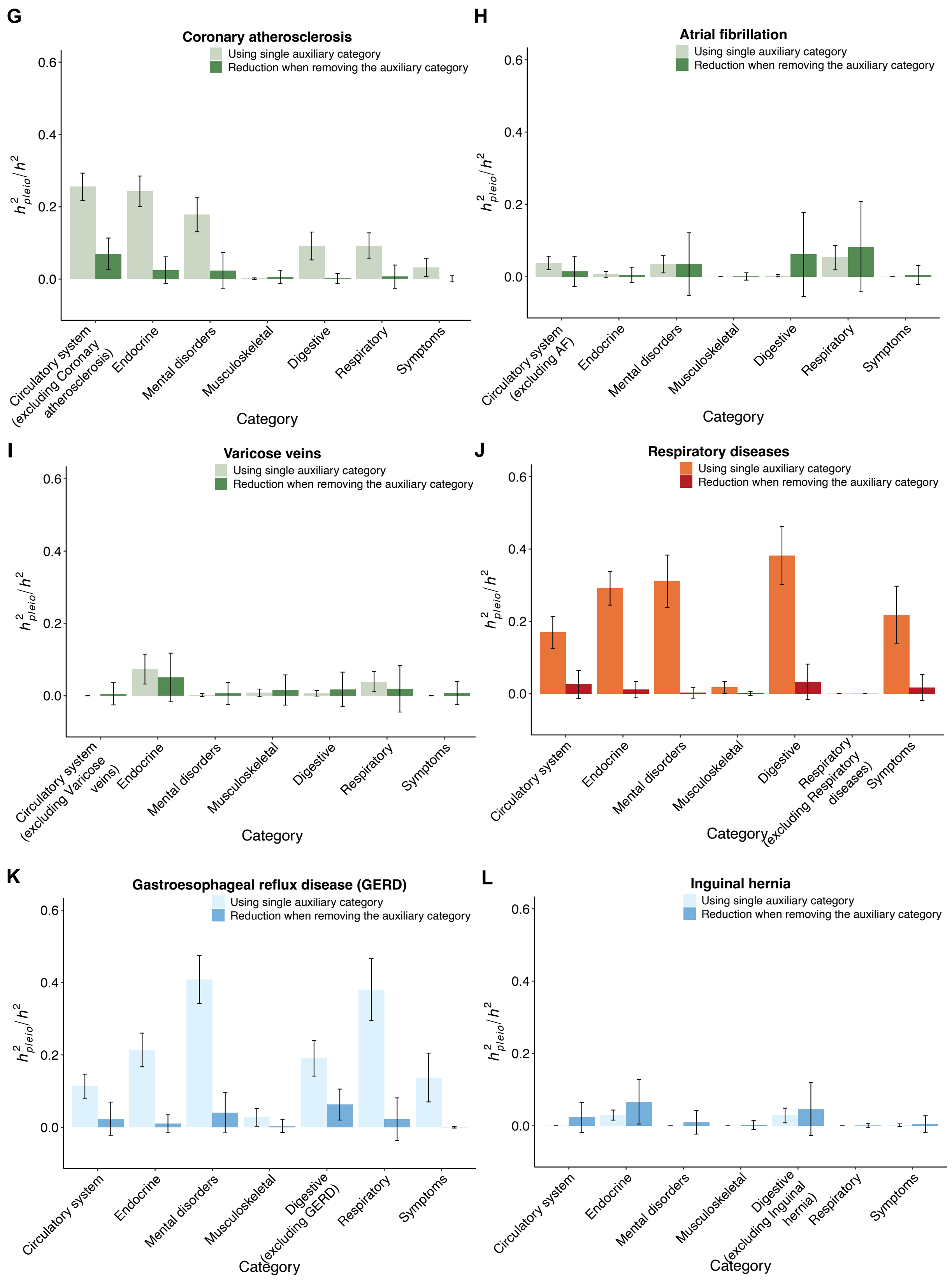
G
Coronary atherosclerosis
Using single auxiliary category
Reduction when removing the auxiliary category
Circulatory system (excluding Coronary atherosclerosis)
Endocrine
Mental disorders
Musculoskeletal
Digestive
Respiratory
Symptoms
Category
H
Atrial fibrillation
Circulatory system (excluding AF)
I
Varicose veins
Circulatory system (excluding Varicose veins)
J
Respiratory diseases
Respiratory (excluding Respiratory diseases)
K
Gastroesophageal reflux disease (GERD)
Digestive (excluding GERD)
L
Inguinal hernia
Digestive (excluding Inguinal hernia)
$h^2_{pleio}/h^2$
0.0
0.2
0.4
0.6

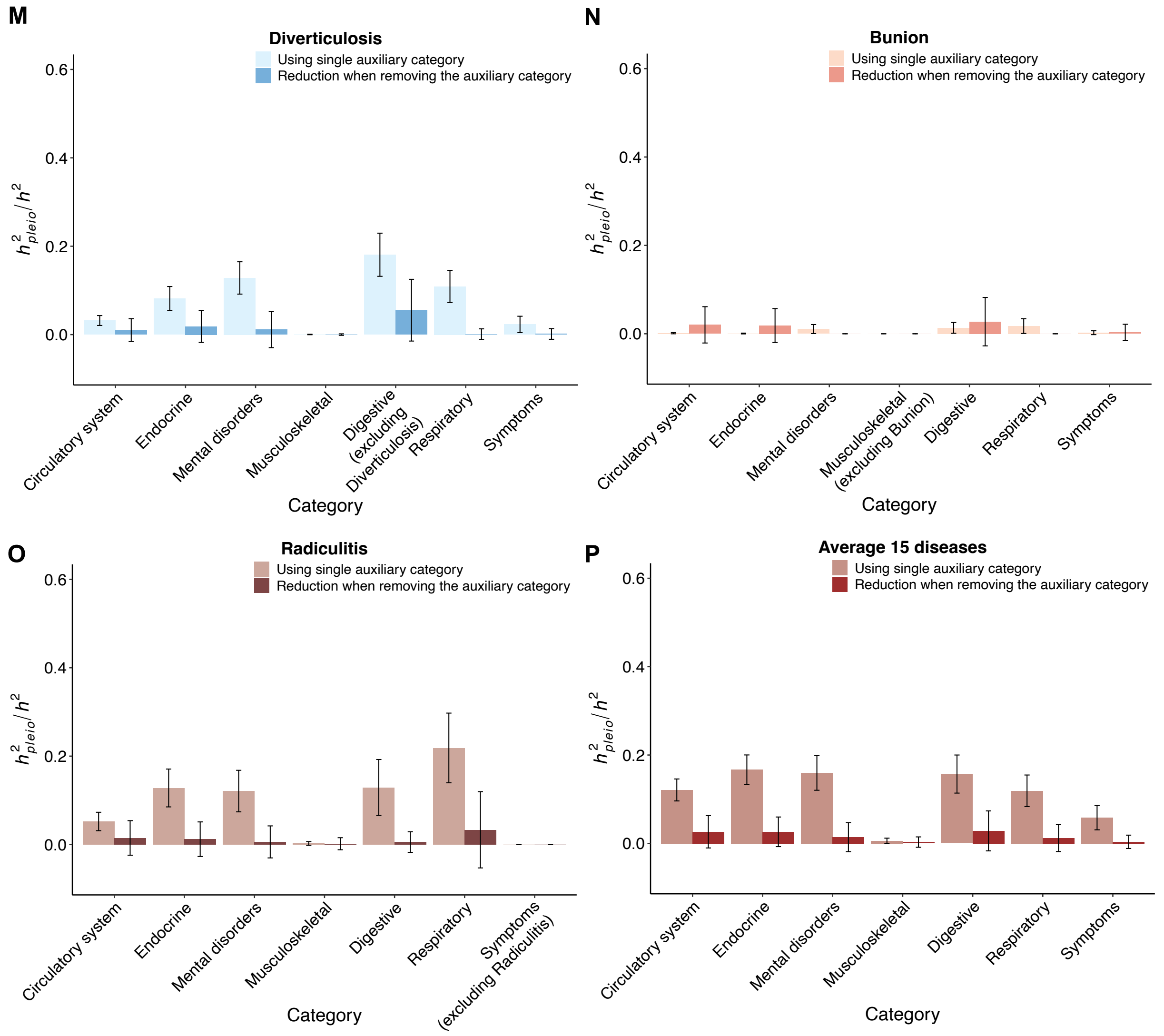


**Supplementary Figure 17. Distribution of $\boldsymbol{h^2}_{\boldsymbol{pleio}}/\boldsymbol{h^2}$ across Phecode disease categories.** (A-O) Comparison of $h^2_{pleio}/h^2$ between single-auxiliary-category estimate and reduction when removing the auxiliary category for all 15 UK Biobank target diseases and (P) the average across 15 UK Biobank diseases. Light color bar is the single-auxiliary-category estimate of $h^2_{pleio}/h^2$, dark color bar is the reduction of $h^2_{pleio}/h^2$ when removing the auxiliary category. The zero $h^2_{pleio}/h^2$ of single-auxiliary-category estimate indicates that this auxiliary category only has the target disease. The zero reduction of $h^2_{pleio}/h^2$ when removing the auxiliary category indicates that the estimation of $h^2_{pleio}/h^2$ for all auxiliary diseases has already excluded those auxiliary diseases in the removed auxiliary category after the pruning procedure. Therefore, further removal of this auxiliary category makes no difference. Different colors for different disease panels represent their target disease category. Error bar shows the jackknife standard error. Detailed results are provided in **Supplementary Table 19**. Abbreviation: T2D: type 2 diabetes; MDD: depression; HTN: hypertension; GERD: gastroesophageal reflux disease; AF: Atrial Fibrillation.

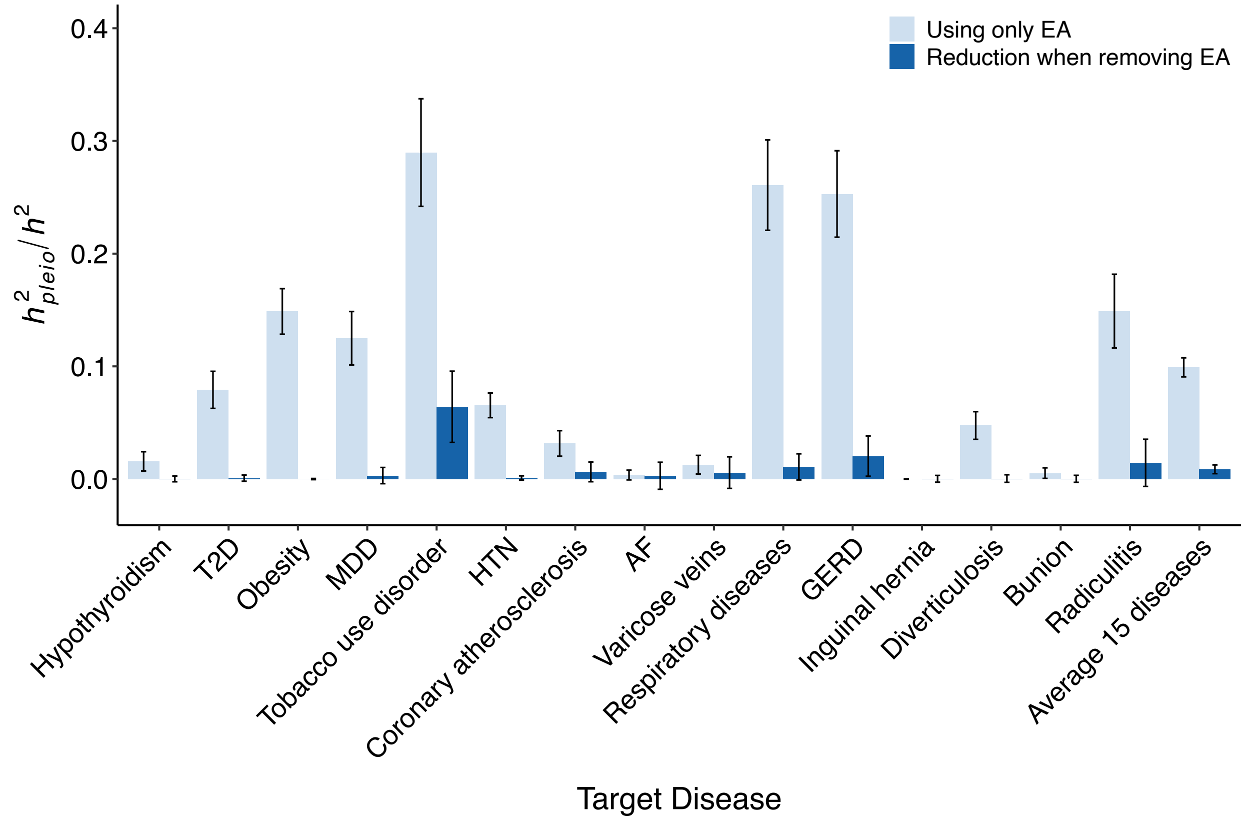


**Supplementary Figure 18. The impact of educational attainment (EA) on pleiotropy.** We performed two analyses. First, we estimated ${h^2}_{pleio}/h^2$ with respect to years of education as the only auxiliary trait for each of the 15 UK Biobank diseases (Light color bar is the ${h^2}_{pleio}/h^2$ using only EA as auxiliary trait with average of 9.9% (s.e. 0.8%)). Second, we assessed the impact of removing EA from the set of auxiliary traits by estimating the difference between (i) ${h^2}_{pleio}/h^2$ with respect to 15 UK Biobank auxiliary diseases + 17 UK Biobank auxiliary quantitative traits vs. (ii) ${h^2}_{pleio}/h^2$ with respect to 15 UK Biobank auxiliary diseases + 16 UK Biobank quantitative traits excluding EA (Dark color bar is the reduction of ${h^2}_{pleio}/h^2$ when removing EA from the auxiliary trait set with average of 0.87% (s.e. 0.39%). Error bar shows the jackknife standard error. Detailed results are provided in **Supplementary Table 28**. Abbreviation: T2D: type 2 diabetes; MDD: depression; HTN: hypertension; GERD: gastroesophageal reflux disease; AF: Atrial Fibrillation.

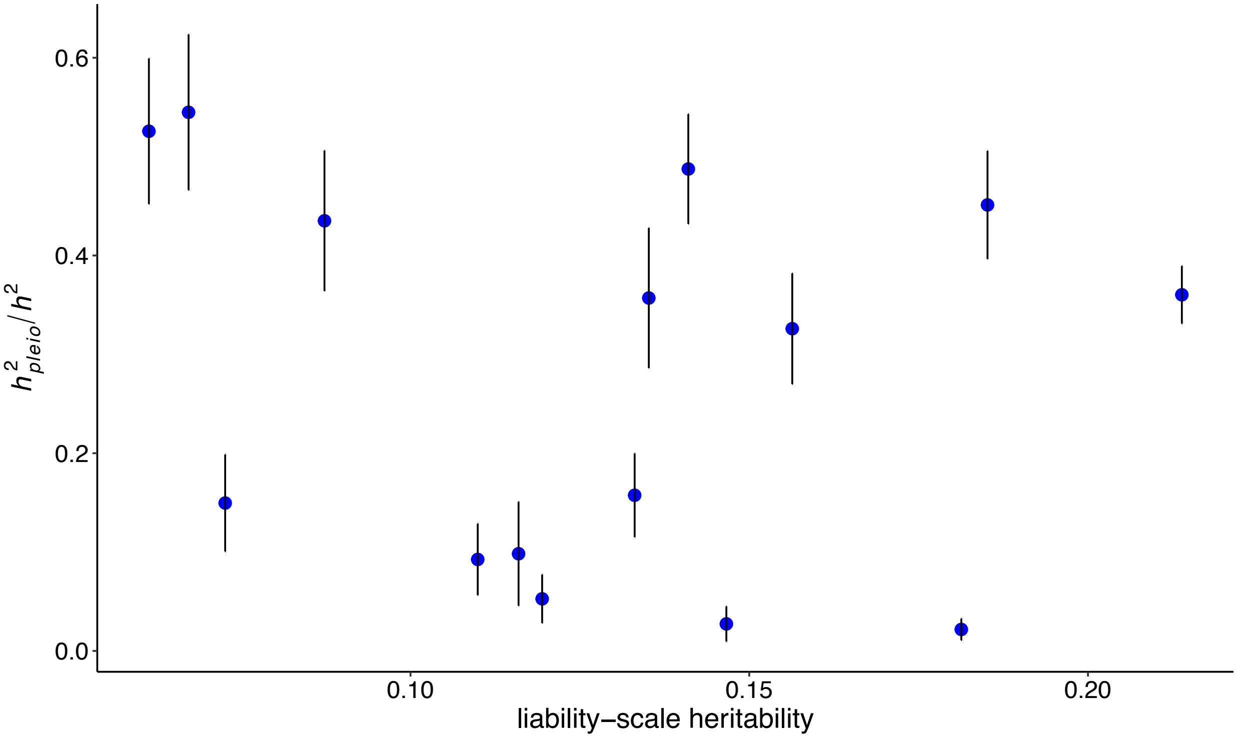


**Supplementary Figure 19. Comparison on liability-scale heritability and $h^2{}_{pleio}/h^2$ across the 15 UK Biobank diseases.** We observed no significant correlation (correlation = -0.18 (*P* = 0.52)).

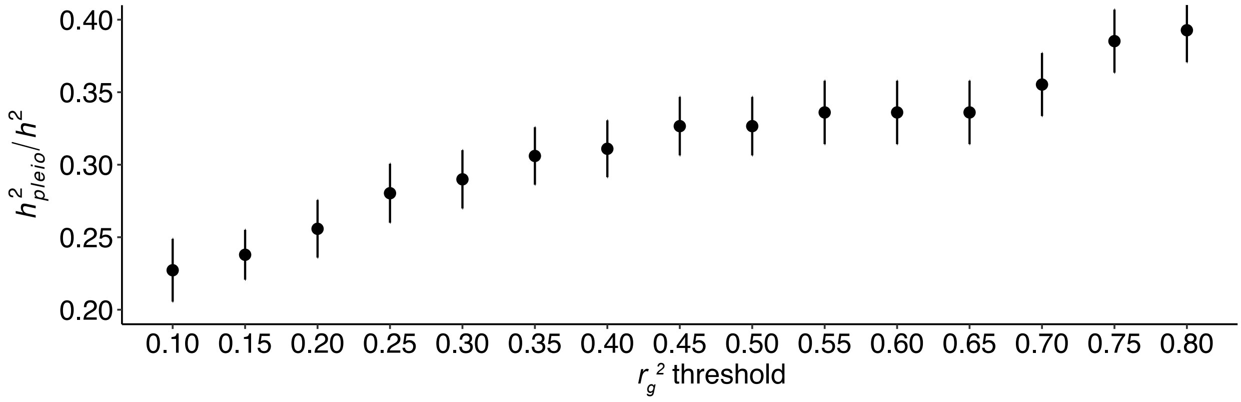


**Supplementary Figure 20. Changes in average $h^2{}_{pleio}/h^2$ across 15 UK Biobank diseases for $r_g{}^2$ thresholds between target and auxiliary diseases ranging from 0.1 to 0.8 (step = 0.05).** Average $h^2{}_{pleio}/h^2$ increased at $r_g{}^2$ thresholds larger than 0.5, but we consider this increase to be not biologically meaningful because most instances of $r_g{}^2$ > 0.5 involved auxiliary traits that *are not biologically distinct* from the target trait, e.g. hypertension-diastolic blood pressure ($r_g{}^2$ = 0.72), hypertension-systolic blood pressure ($r_g{}^2$ = 0.72), type 2 diabetes-glucose ($r_g{}^2$ = 0.69), type 2 diabetes-HbA1C ($r_g{}^2$ = 0.79). In addition, average $h^2{}_{pleio}/h^2$ decreased at $r_g{}^2$ thresholds smaller than 0.5, but all instances of 0.25 < $r_g{}^2$ < 0.5 involved auxiliary traits that *are biologically distinct* from the target trait: coronary atherosclerosis-hypertension ($r_g{}^2$ = 0.25), coronary atherosclerosis-type 2 diabetes ($r_g{}^2$ = 0.25), hypertension-type 2 diabetes ($r_g{}^2$ = 0.26), depression-tobacco use disorder ($r_g{}^2$ = 0.35), gastroesophageal reflux disease (GERD)-tobacco use disorder ($r_g{}^2$ = 0.27), obesity-type 2 diabetes ($r_g{}^2$ = 0.36), obesity-respiratory diseases ($r_g{}^2$ = 0.29), obesity- HbA1c ($r_g{}^2$ = 0.26), obesity-glucose ($r_g{}^2$ = 0.26), type 2 diabetes-body WHR ($r_g{}^2$ = 0.36), type 2 diabetes-BMI ($r_g{}^2$ = 0.34), type 2 diabetes-triglycerides ($r_g{}^2$ = 0.31), gastroesophageal reflux disease (GERD)-depression ($r_g{}^2$ = 0.40), gastroesophageal reflux disease (GERD)-respiratory diseases ($r_g{}^2$ = 0.39) and depression-respiratory diseases ($r_g{}^2$ = 0.32)—such that we believe that an estimand based on an $r_g{}^2$ threshold of 0.5 is preferred. Detailed genetic correlation results refer to **Supplementary Table 29**.

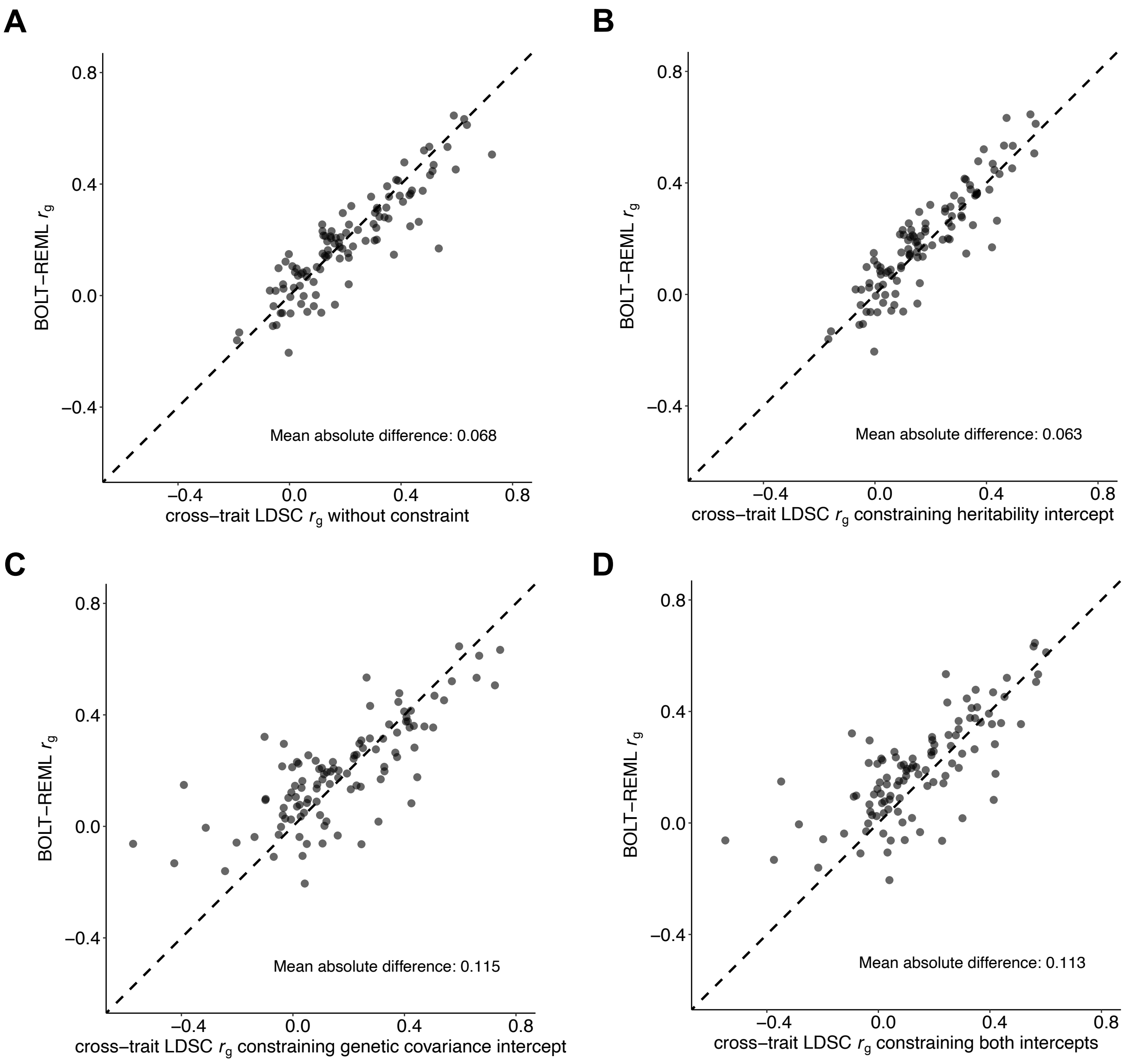


**Supplementary Figure 21. Comparison on $r_g$ from BOLT-REML and $r_g$ from cross-trait LDSC with 4 options of constraining intercept.** (A) Compare $r_g$ from BOLT-REML and $r_g$ from cross-trait LDSC without constraint on intercept, which is the default version we used in main analyses. (B-D) Compare $r_g$ from BOLT-REML and $r_g$ from three modified version of cross-trait LDSC: constraining heritability intercept, constraining genetic covariance intercept, and constraining both intercepts. The computation of constrained intercept is provided in **Methods** section and **Supplementary Table 27**. Detailed results are provided in **Supplementary Table 20**.

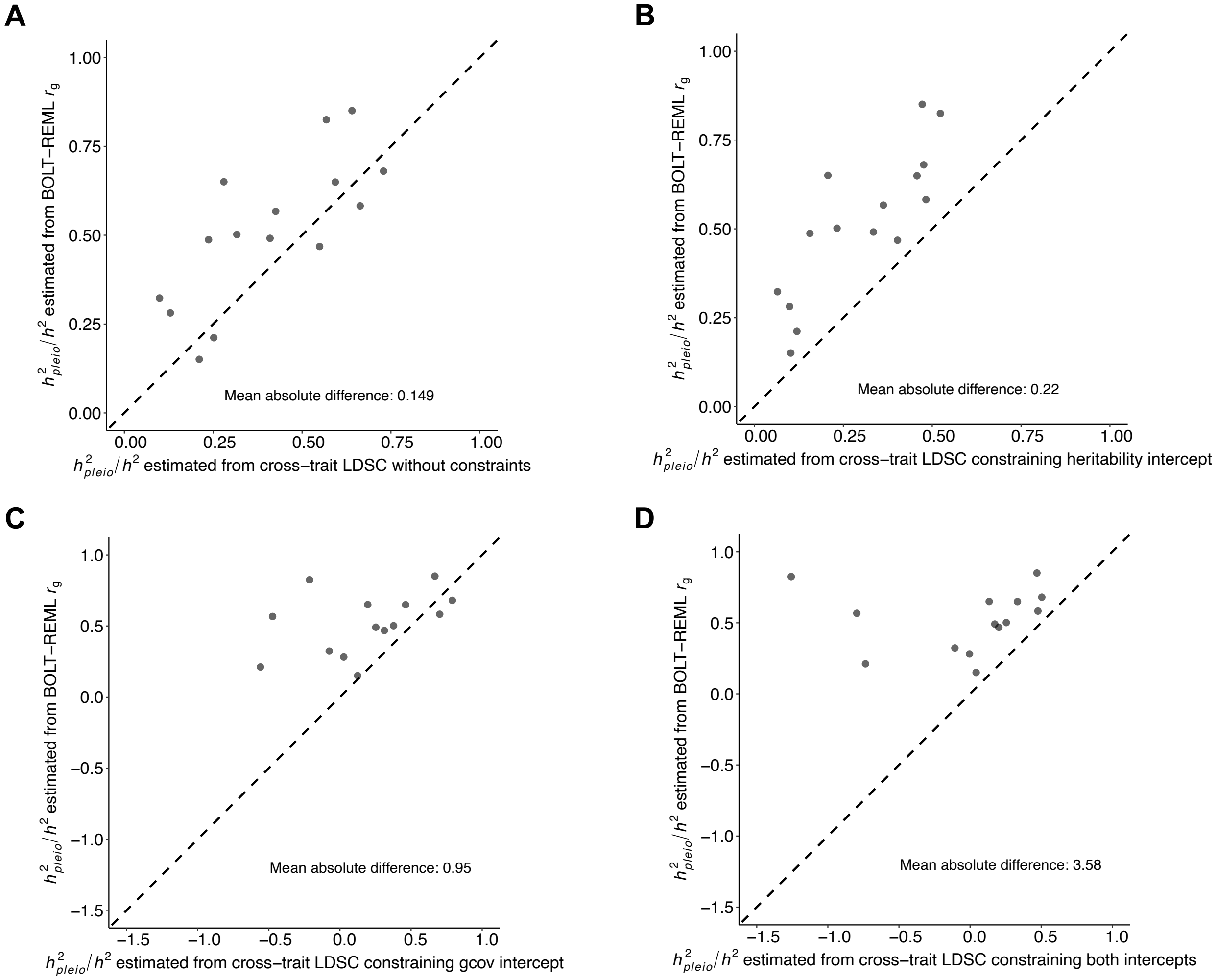


**Supplementary Figure 22. Comparison on $h^2{}_{pleio}/h^2$ estimated from BOLT-REML $r_g$ and $h^2{}_{pleio}/h^2$ estimated from cross-trait LDSC $r_g$ with 4 options of constraining intercept.** (A) Compare $h^2{}_{pleio}/h^2$ estimated from BOLT-REML $r_g$ and $h^2{}_{pleio}/h^2$ estimated from cross-trait LDSC $r_g$ without constraint on intercept, which is the default version we estimated in main analyses. (B-D) Compare $h^2{}_{pleio}/h^2$ estimated from BOLT-REML $r_g$ and $h^2{}_{pleio}/h^2$ estimated from $r_g$ from three modified version of cross-trait LDSC: constraining heritability intercept, constraining genetic covariance intercept, and constraining both intercepts. We found that our results on $h^2{}_{pleio}/h^2$ using default version of cross-trait LDSC were broadly consistent with results from BOLT-REML, and all three modified versions deviated from BOLT-REML results in estimates of $h^2{}_{pleio}/h^2$. One outlier dot was removed from both panels C and D for clarity in the figure visualization, while the mean absolute difference was computed based on the full 15 diseases. Detailed results are provided in **Supplementary Table 21**.

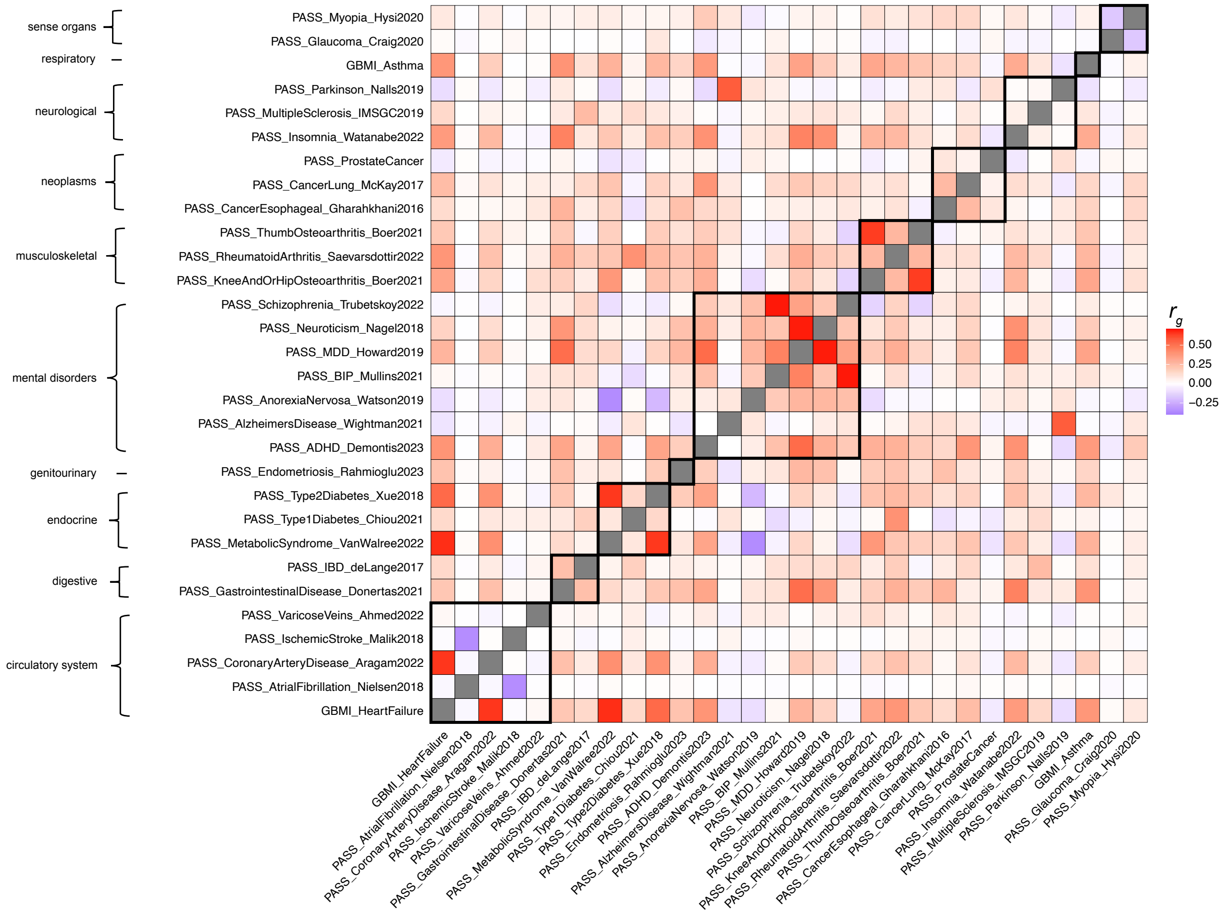


**Supplementary Figure 23. Estimates of genetic correlation of 30 non-UK Biobank diseases.** Most diseases have moderate genetic correlations within disease categories and between disease categories. Black boxes demarcate correlations between diseases within the same disease category. Detailed results are provided in **Supplementary Table 16**.

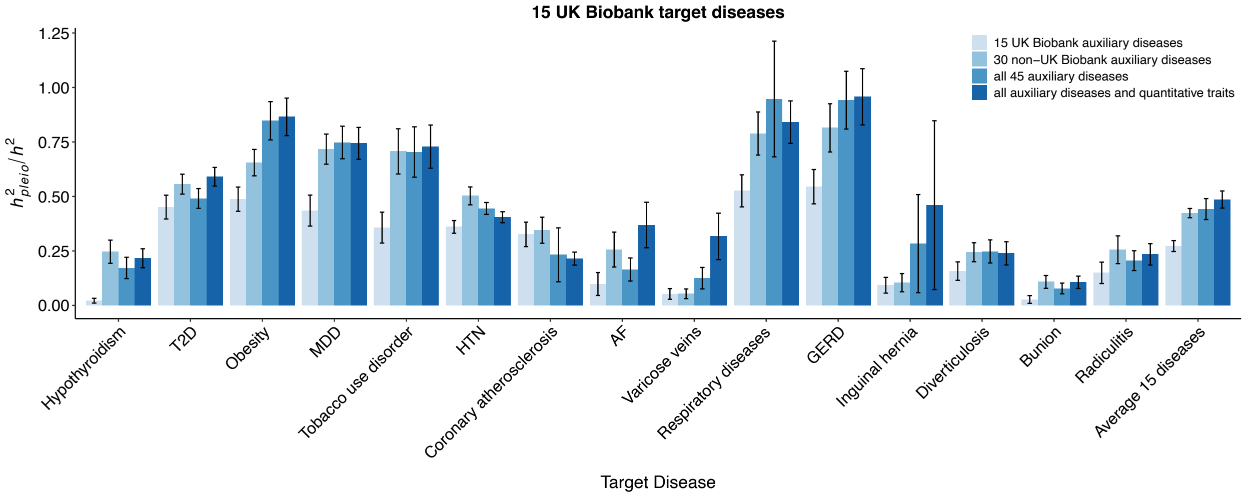


**Supplementary Figure 24.** $h^2{}_{pleio}/h^2$ **of all 15 UK Biobank target diseases with respect to four types of auxiliary disease sets: 15 UK Biobank auxiliary diseases, 30 non-UK Biobank auxiliary diseases, all 45 auxiliary diseases and all 62 auxiliary diseases and quantitative traits.** Error bar shows the jackknife standard error. Detailed results are provided in **Supplementary Table 22**. Abbreviation: T2D: type 2 diabetes; MDD: depression; HTN: hypertension; GERD: gastroesophageal reflux disease; AF: Atrial Fibrillation.

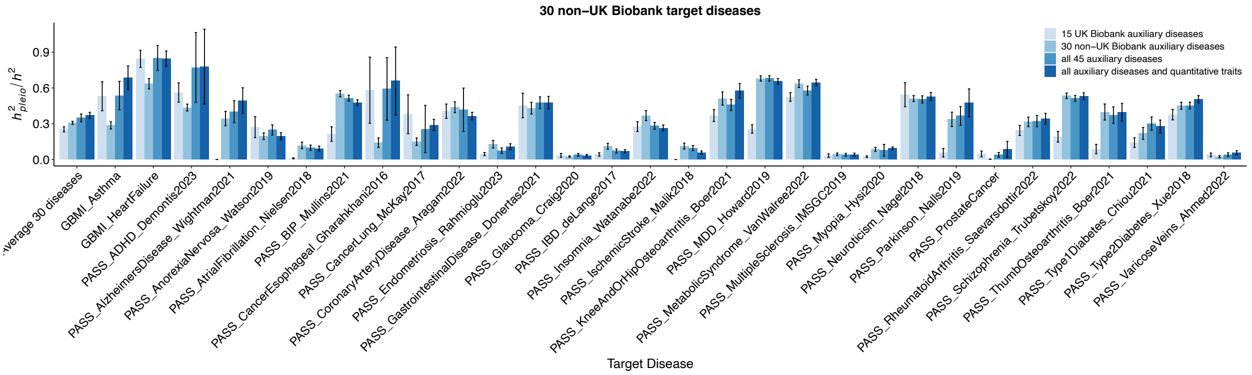


**Supplementary Figure 25**. $h^2{}_{pleio}/h^2$ **of all 30 non-UK Biobank target diseases with respect to four types of auxiliary disease sets: 15 UK Biobank auxiliary diseases, 30 non-UK Biobank auxiliary diseases, all 45 auxiliary diseases and all 62 auxiliary diseases and quantitative traits.** Error bar shows the jackknife standard error. Detailed results are provided in **Supplementary Table 23**.

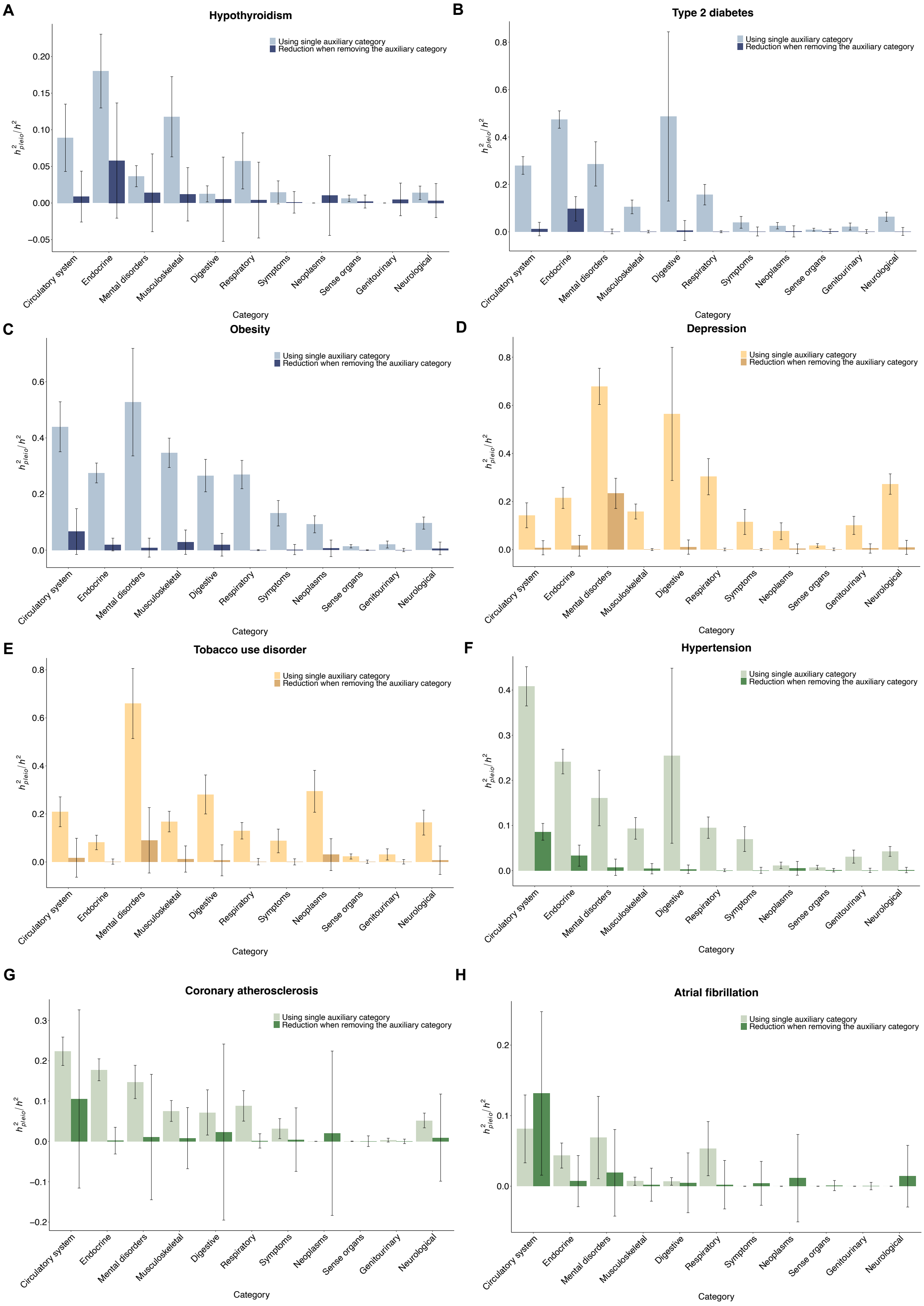
A
Hypothyroidism
B
Type 2 diabetes
C
Obesity
D
Depression
E
Tobacco use disorder
F
Hypertension
G
Coronary atherosclerosis
H
Atrial fibrillation
Using single auxiliary category
Reduction when removing the auxiliary category
$h^2_{pleio}/h^2$
Category
Circulatory system
Endocrine
Mental disorders
Musculoskeletal
Digestive
Respiratory
Symptoms
Neoplasms
Sense organs
Genitourinary
Neurological

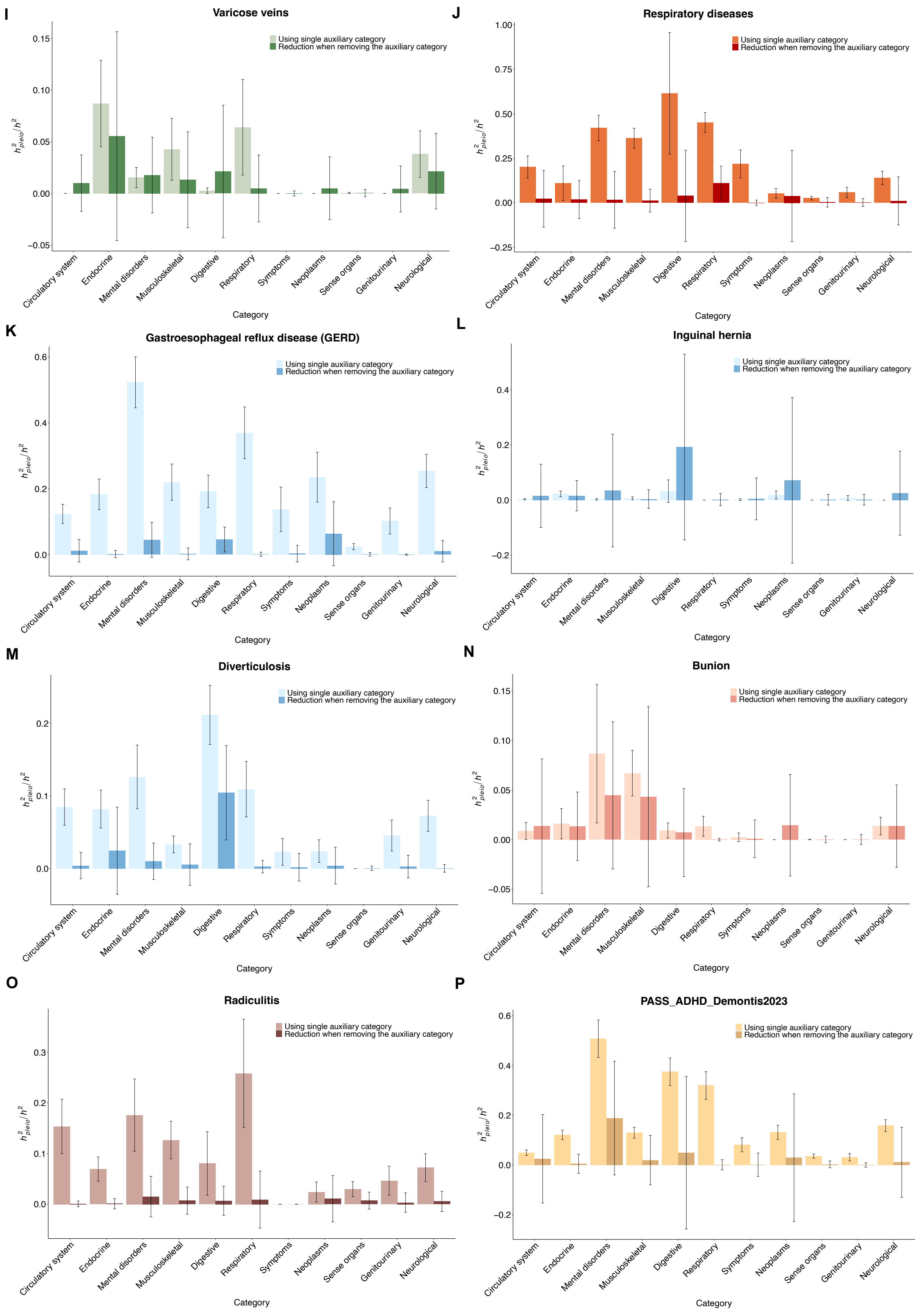

I
Varicose veins
Using single auxiliary category
Reduction when removing the auxiliary category
$h^2_{pleio}/h^2$
0.15
0.10
0.05
0.00
−0.05
Circulatory system
Endocrine
Mental disorders
Musculoskeletal
Digestive
Respiratory
Symptoms
Neoplasms
Sense organs
Genitourinary
Neurological
Category
J
Respiratory diseases
Using single auxiliary category
Reduction when removing the auxiliary category
$h^2_{pleio}/h^2$
1.00
0.75
0.50
0.25
0.00
−0.25
Circulatory system
Endocrine
Mental disorders
Musculoskeletal
Digestive
Respiratory
Symptoms
Neoplasms
Sense organs
Genitourinary
Neurological
Category
K
Gastroesophageal reflux disease (GERD)
Using single auxiliary category
Reduction when removing the auxiliary category
$h^2_{pleio}/h^2$
0.6
0.4
0.2
0.0
Circulatory system
Endocrine
Mental disorders
Musculoskeletal
Digestive
Respiratory
Symptoms
Neoplasms
Sense organs
Genitourinary
Neurological
Category
L
Inguinal hernia
Using single auxiliary category
Reduction when removing the auxiliary category
$h^2_{pleio}/h^2$
0.4
0.2
0.0
−0.2
Circulatory system
Endocrine
Mental disorders
Musculoskeletal
Digestive
Respiratory
Symptoms
Neoplasms
Sense organs
Genitourinary
Neurological
Category
M
Diverticulosis
Using single auxiliary category
Reduction when removing the auxiliary category
$h^2_{pleio}/h^2$
0.2
0.1
0.0
Circulatory system
Endocrine
Mental disorders
Musculoskeletal
Digestive
Respiratory
Symptoms
Neoplasms
Sense organs
Genitourinary
Neurological
Category
N
Bunion
Using single auxiliary category
Reduction when removing the auxiliary category
$h^2_{pleio}/h^2$
0.15
0.10
0.05
0.00
−0.05
Circulatory system
Endocrine
Mental disorders
Musculoskeletal
Digestive
Respiratory
Symptoms
Neoplasms
Sense organs
Genitourinary
Neurological
Category
O
Radiculitis
Using single auxiliary category
Reduction when removing the auxiliary category
$h^2_{pleio}/h^2$
0.3
0.2
0.1
0.0
Circulatory system
Endocrine
Mental disorders
Musculoskeletal
Digestive
Respiratory
Symptoms
Neoplasms
Sense organs
Genitourinary
Neurological
Category
P
PASS_ADHD_Demontis2023
Using single auxiliary category
Reduction when removing the auxiliary category
$h^2_{pleio}/h^2$
0.6
0.4
0.2
0.0
−0.2
Circulatory system
Endocrine
Mental disorders
Musculoskeletal
Digestive
Respiratory
Symptoms
Neoplasms
Sense organs
Genitourinary
Neurological
Category

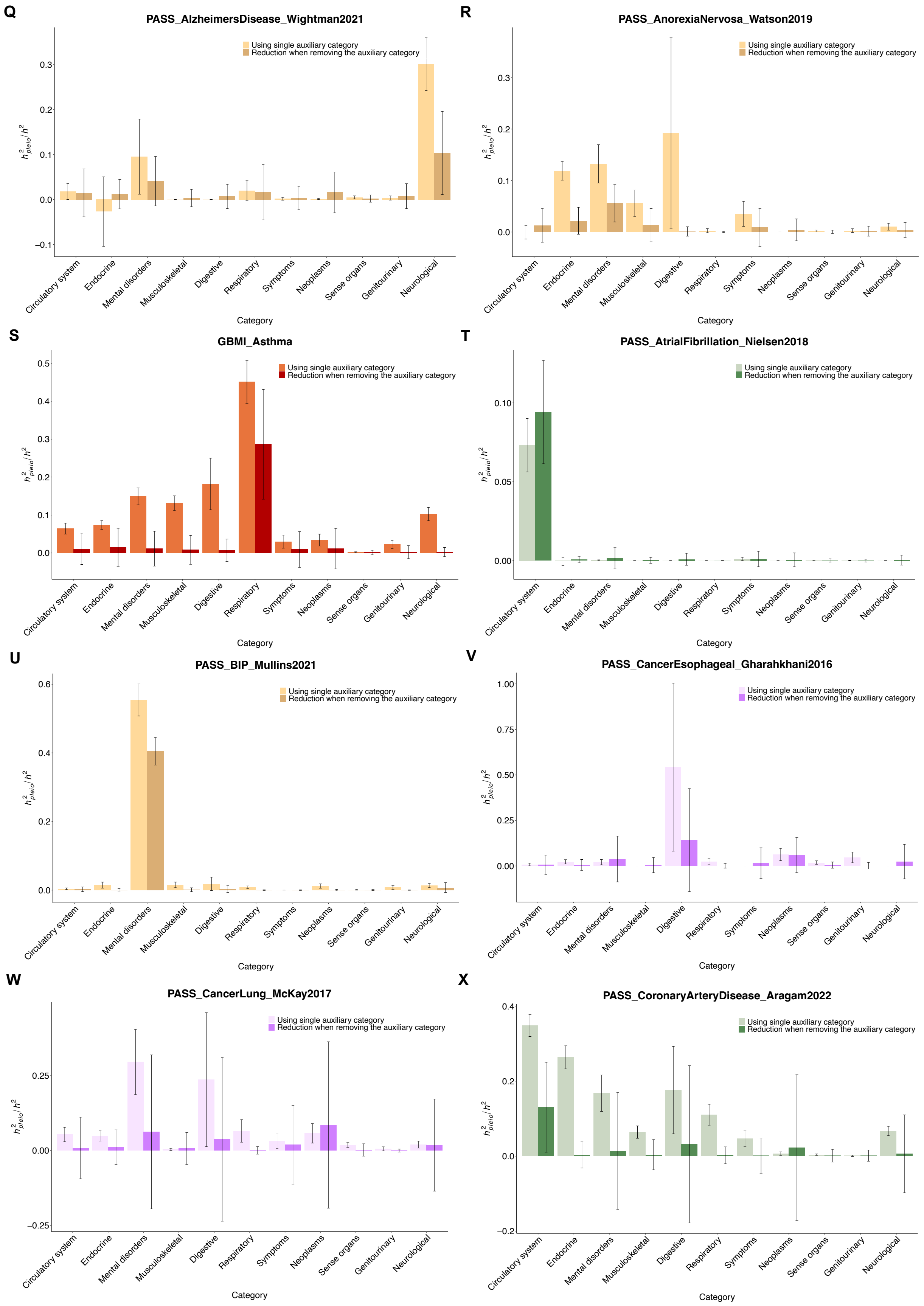
Q
PASS_AlzheimersDisease_Wightman2021
R
PASS_AnorexiaNervosa_Watson2019
S
GBMI_Asthma
T
PASS_AtrialFibrillation_Nielsen2018
U
PASS_BIP_Mullins2021
V
PASS_CancerEsophageal_Gharahkhani2016
W
PASS_CancerLung_McKay2017
X
PASS_CoronaryArteryDisease_Aragam2022
Using single auxiliary category
Reduction when removing the auxiliary category
$h^2_{pleio}/h^2$
Category
Circulatory system
Endocrine
Mental disorders
Musculoskeletal
Digestive
Respiratory
Symptoms
Neoplasms
Sense organs
Genitourinary
Neurological

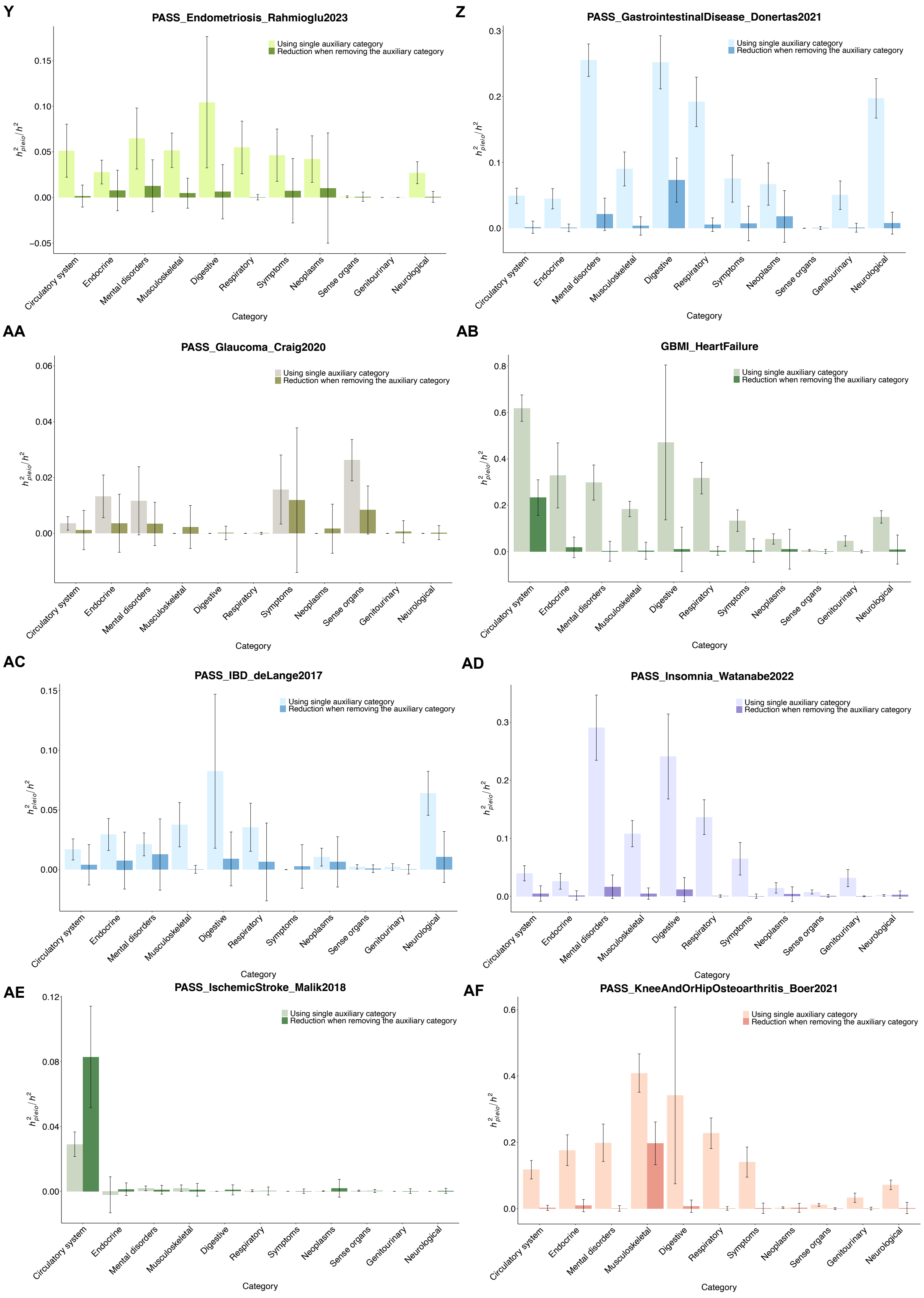
Y
PASS_Endometriosis_Rahmioglu2023
Using single auxiliary category
Reduction when removing the auxiliary category
h2pleio/h2
Category
Circulatory system
Endocrine
Mental disorders
Musculoskeletal
Digestive
Respiratory
Symptoms
Neoplasms
Sense organs
Genitourinary
Neurological
Z
PASS_GastrointestinalDisease_Donertas2021
Using single auxiliary category
Reduction when removing the auxiliary category
AA
PASS_Glaucoma_Craig2020
Using single auxiliary category
Reduction when removing the auxiliary category
AB
GBMI_HeartFailure
Using single auxiliary category
Reduction when removing the auxiliary category
AC
PASS_IBD_deLange2017
Using single auxiliary category
Reduction when removing the auxiliary category
AD
PASS_Insomnia_Watanabe2022
Using single auxiliary category
Reduction when removing the auxiliary category
AE
PASS_IschemicStroke_Malik2018
Using single auxiliary category
Reduction when removing the auxiliary category
AF
PASS_KneeAndOrHipOsteoarthritis_Boer2021
Using single auxiliary category
Reduction when removing the auxiliary category

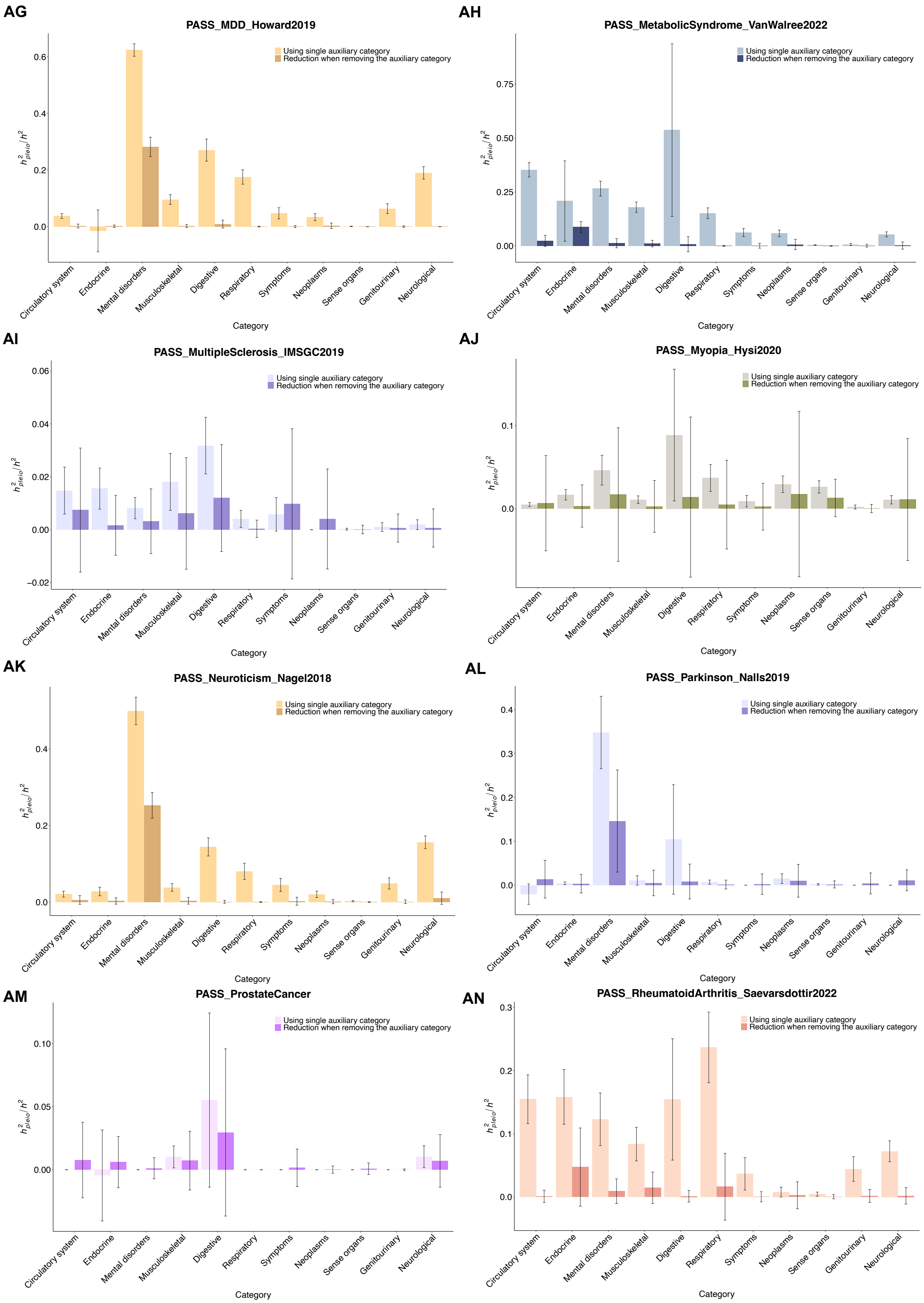
AG
PASS_MDD_Howard2019
Using single auxiliary category
Reduction when removing the auxiliary category
AH
PASS_MetabolicSyndrome_VanWalree2022
Using single auxiliary category
Reduction when removing the auxiliary category
AI
PASS_MultipleSclerosis_IMSGC2019
Using single auxiliary category
Reduction when removing the auxiliary category
AJ
PASS_Myopia_Hysi2020
Using single auxiliary category
Reduction when removing the auxiliary category
AK
PASS_Neuroticism_Nagel2018
Using single auxiliary category
Reduction when removing the auxiliary category
AL
PASS_Parkinson_Nalls2019
Using single auxiliary category
Reduction when removing the auxiliary category
AM
PASS_ProstateCancer
Using single auxiliary category
Reduction when removing the auxiliary category
AN
PASS_RheumatoidArthritis_Saevarsdottir2022
Using single auxiliary category
Reduction when removing the auxiliary category
$h^2_{pleio}/h^2$
Circulatory system
Endocrine
Mental disorders
Musculoskeletal
Digestive
Respiratory
Symptoms
Neoplasms
Sense organs
Genitourinary
Neurological
Category

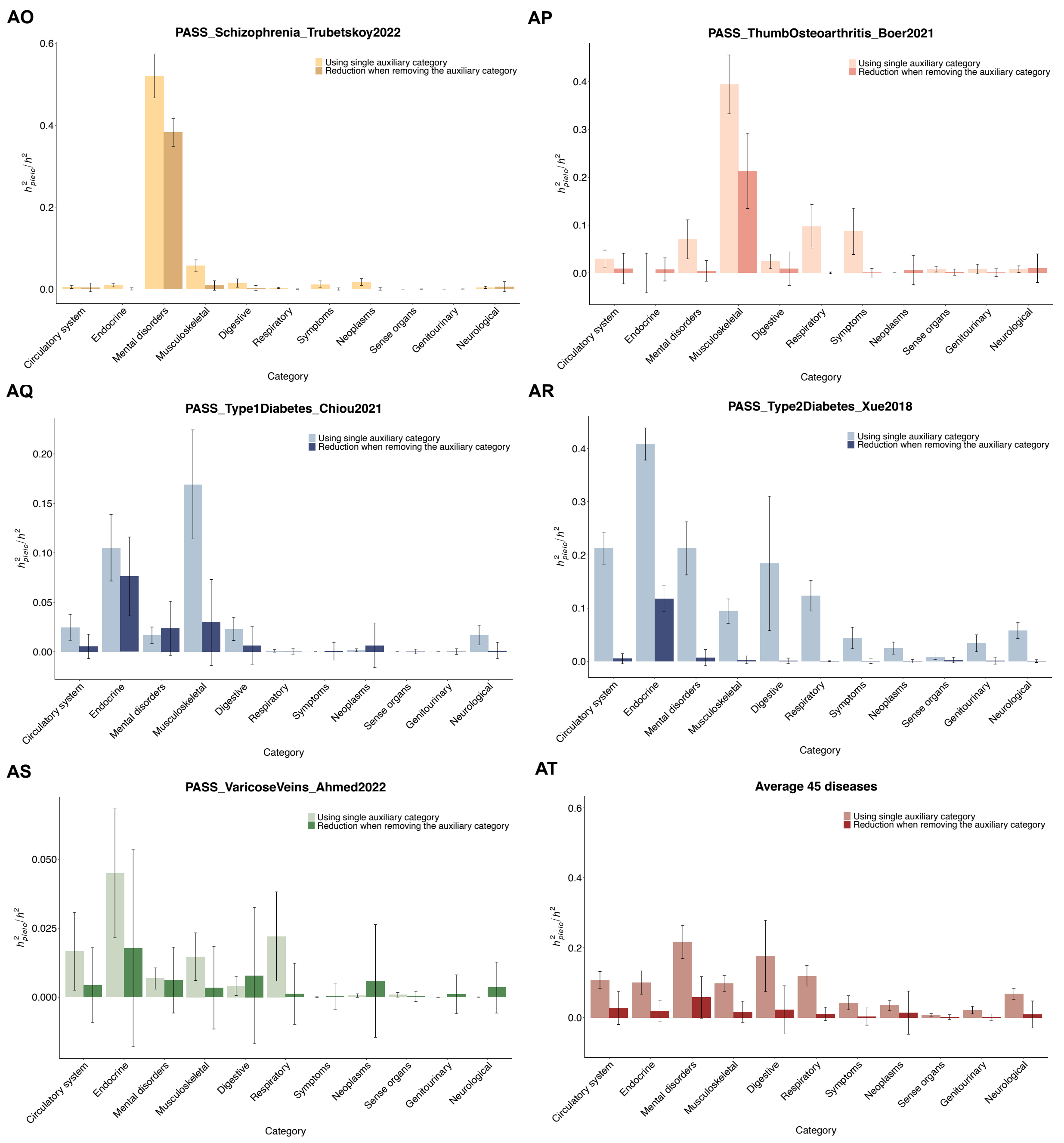


**Supplementary Figure 26. Distribution of $h^2{}_{pleio}/h^2$ across Phecode disease categories using all 45 auxiliary diseases.** (A-O) Comparison of $h^2{}_{pleio}/h^2$ between single-auxiliary-category estimate and reduction when removing the auxiliary category for all 15 UK Biobank target diseases, (P-AS) for all 30 non-UK Biobank target diseases, and (AT) the average across all 45 UK Biobank + non-UK Biobank diseases. Light color bar is the single-auxiliary-category estimate of $h^2{}_{pleio}/h^2$, dark color bar is the reduction of $h^2{}_{pleio}/h^2$ when removing the auxiliary category. The zero $h^2{}_{pleio}/h^2$ of single-auxiliary-category estimate indicates that this auxiliary category only has the target disease. The zero reduction of $h^2{}_{pleio}/h^2$ when removing the auxiliary category indicates that the estimation of $h^2{}_{pleio}/h^2$ for all auxiliary diseases has already excluded those auxiliary diseases in the removed auxiliary category after the pruning procedure. Therefore, further removal of this auxiliary category makes no difference. Different colors of different disease panels represent their target disease category. Error bar shows the jackknife standard error. When the target disease category serves as the auxiliary category, auxiliary diseases within this category exclude the target disease. Detailed results are provided in **Supplementary Table 24**. Abbreviation: T2D: type 2 diabetes; MDD: depression; HTN: hypertension; GERD: gastroesophageal reflux disease; AF: Atrial Fibrillation.

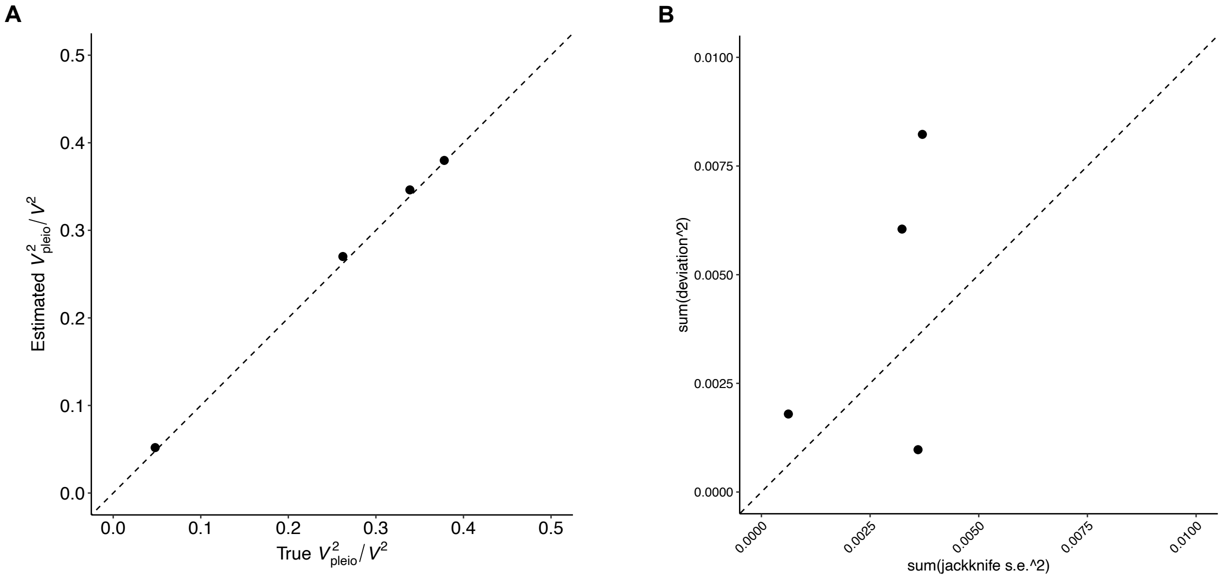


**Supplementary Figure 27. Simulations of $V^2_{pleio}/V^2$ and its standard error.** True $r_o$ is set to 0.5 within diseases categories and 0.1 between diseases categories for the 15 diseases (based on the 7 Phecode disease categories), which implies 4 different values of true $V^2_{pleio}/V^2$ for each target disease (ranging from 0.05 to 0.38). We simulated liabilities for 228,258 individuals. We computed true $r_l$ and true $V^2_{pleio}$ based on the simulated liabilities. Then, we used liability threshold model to generate binary phenotypes based on the empirical prevalence for these 15 UKB diseases. We applied our method on the simulated binary phenotypes to estimate $V^2_{pleio}$ and compared it to the true value. (A) Estimated $V^2_{pleio}/V^2$ is unbiased without the need for bias correction, in which error bars are smaller than dot size in some cases. (B) Deviation is the difference between true $V^2_{pleio}/V^2$ and the average estimated $V^2_{pleio}/V^2$ across diseases that have the same true $V^2_{pleio}/V^2$. We estimate the standard error by jackknifing over blocks of individuals across all diseases. The estimated standard errors of $V^2_{pleio}/V^2$ are anti-conservative, as the ratio of the average estimated squared jackknife standard error for $V^2_{pleio}/V^2$ to the average squared deviation across simulations was equal to 0.65 (jackknife s.e. 0.04); we determined that this does not impact our results, as the standard errors of $V^2_{pleio}/V^2$ estimates are small given large the sample size of UK Biobank data. Detailed results are provided in **Supplementary Table 25**.

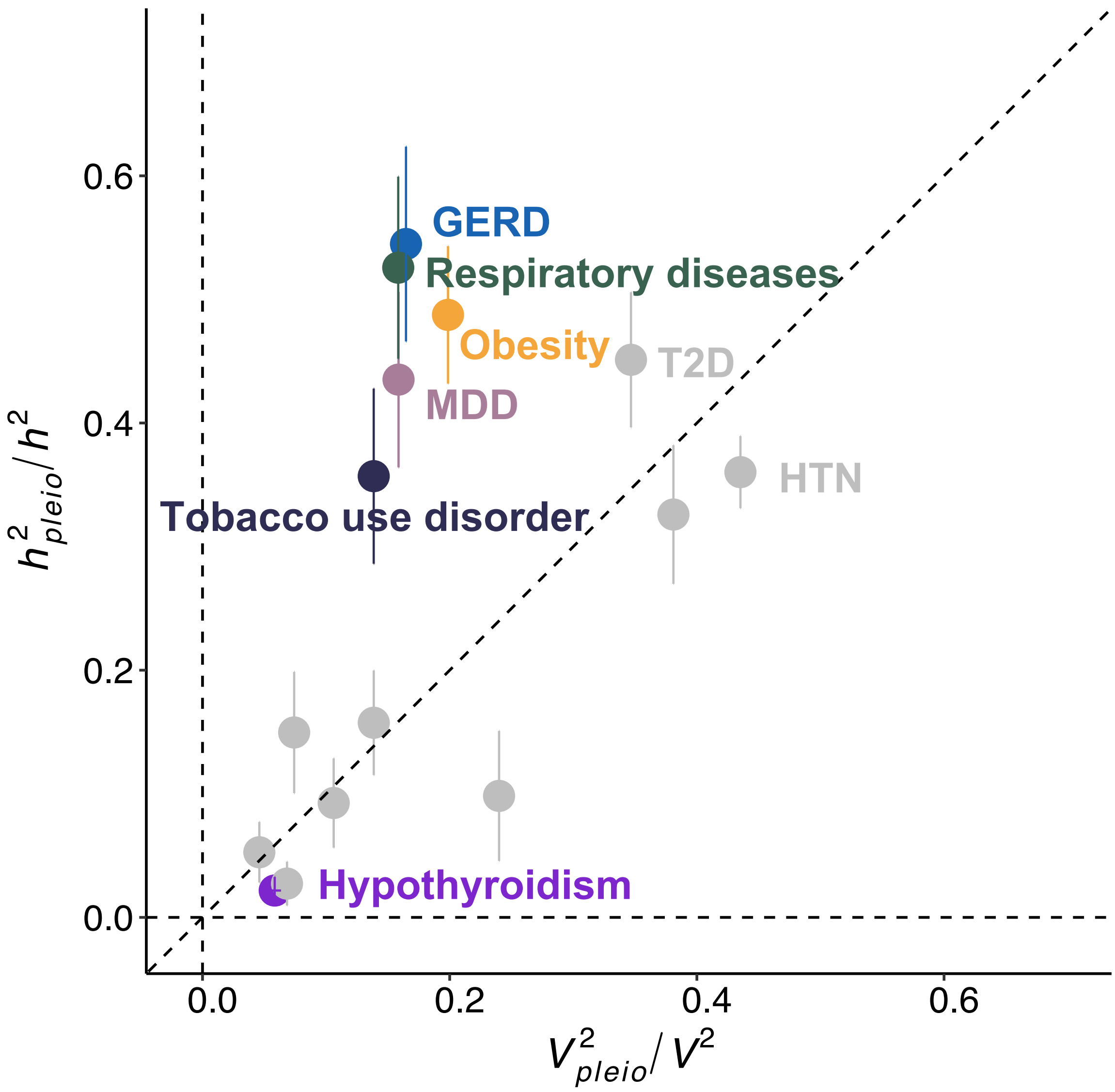


**Supplementary Figure 28. Scatter plot of $h^2{}_{pleio}/h^2$ vs. $V^2{}_{pleio}/V^2$ for the 15 UK Biobank diseases.** We use the same set of auxiliary diseases after pruning in comparing $h^2{}_{pleio}/h^2$ and $V^2{}_{pleio}/V^2$. Four dots with color have $h^2{}_{pleio}/h^2$ estimation significantly different from $V^2{}_{pleio}/V^2$ ($P < 0.05/15$). This figure is the same as **Figure 7** but adding error bar. Error bar shows the jackknife standard error. The estimates of $h^2{}_{pleio}/h^2$ were generally larger than estimates of $V^2{}_{pleio}/V^2$ (ratio of averages = 1.51x (s.e. 0.14)). We compute the ratio of averages using $\frac{average\ h^2{}_{pleio}/h^2\ \text{across 15 diseases}}{average\ V^2{}_{pleio}/V^2\ across\ 15\ diseases}$ and s.e. using $\frac{s.e. of\ average\ h^2{}_{pleio}/h^2\ \text{across 15 diseases}}{average\ V^2{}_{pleio}/V^2\ across\ 15\ diseases}$ which we assume s.e. of average $V^2{}_{pleio}/V^2$ across 15 diseases is 0. Detailed results are provided in **Supplementary Table 26**. Abbreviation: MDD: depression; T2D: type 2 diabetes; HTN: hypertension; GERD: gastroesophageal reflux disease; AF: Atrial Fibrillation.

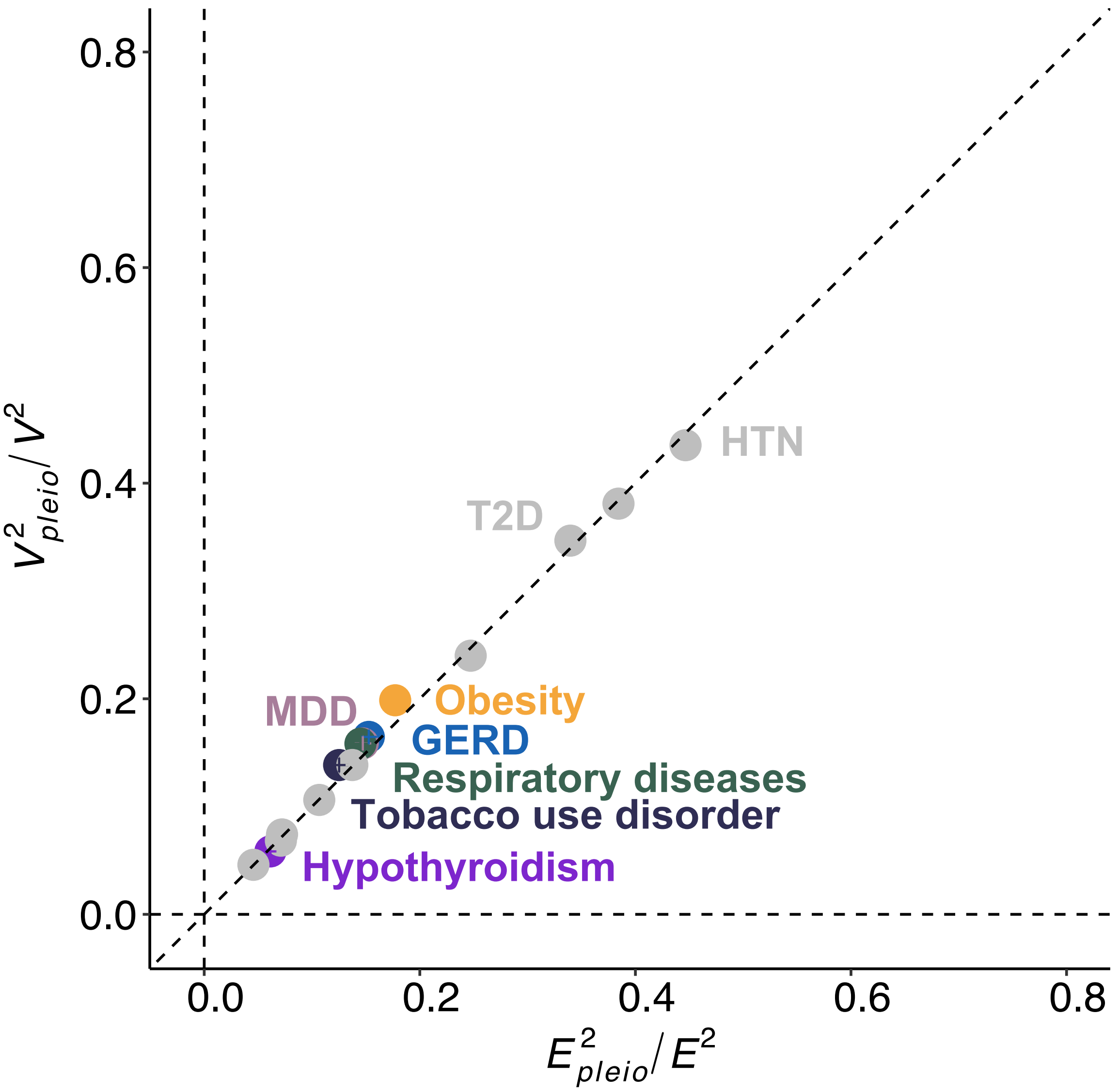


**Supplementary Figure 29. Scatter plot of $V^2{}_{pleio}/V^2$ vs. $E^2{}_{pleio}/E^2$ for the 15 diseases.** We use the same set of auxiliary diseases after pruning in comparing $V^2{}_{pleio}/V^2$ and $E^2{}_{pleio}/E^2$. The colors of dots are the same with **Figure 7**. Error bar shows the jackknife standard error. Detailed results are provided in **Supplementary Table 26**. $E^2{}_{pleio}/E^2$ were slightly smaller than estimates of $V^2{}_{pleio}/V^2$ (ratio of averages = 0.98x). Abbreviation: MDD: depression; T2D: type 2 diabetes; HTN: hypertension; GERD: gastroesophageal reflux disease; AF: Atrial Fibrillation.

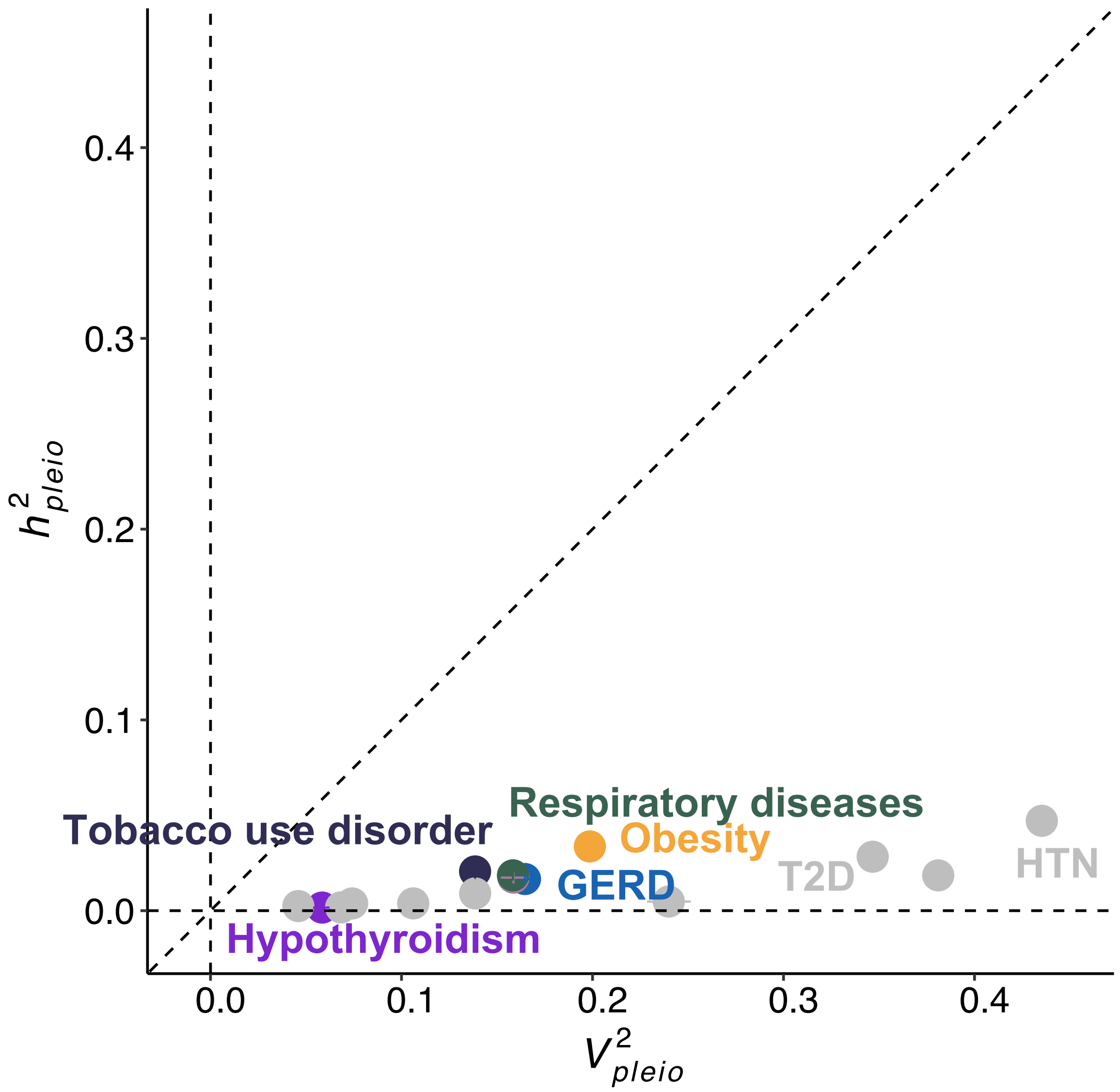


**Supplementary Figure 30. Scatter plot of $h^2{}_{pleio}$ vs. $V^2{}_{pleio}$ for the 15 diseases.** We use the same set of auxiliary diseases after pruning in comparing $h^2{}_{pleio}$ and $V^2{}_{pleio}$ (the ratio of averages = 0.084 (s.e. 0.007). The colors of dots are the same with **Figure 7.** Error bar shows the jackknife standard error. Detailed results are provided in **Supplementary Table 26**. Abbreviation: MDD: depression; T2D: type 2 diabetes; HTN: hypertension; GERD: gastroesophageal reflux disease.

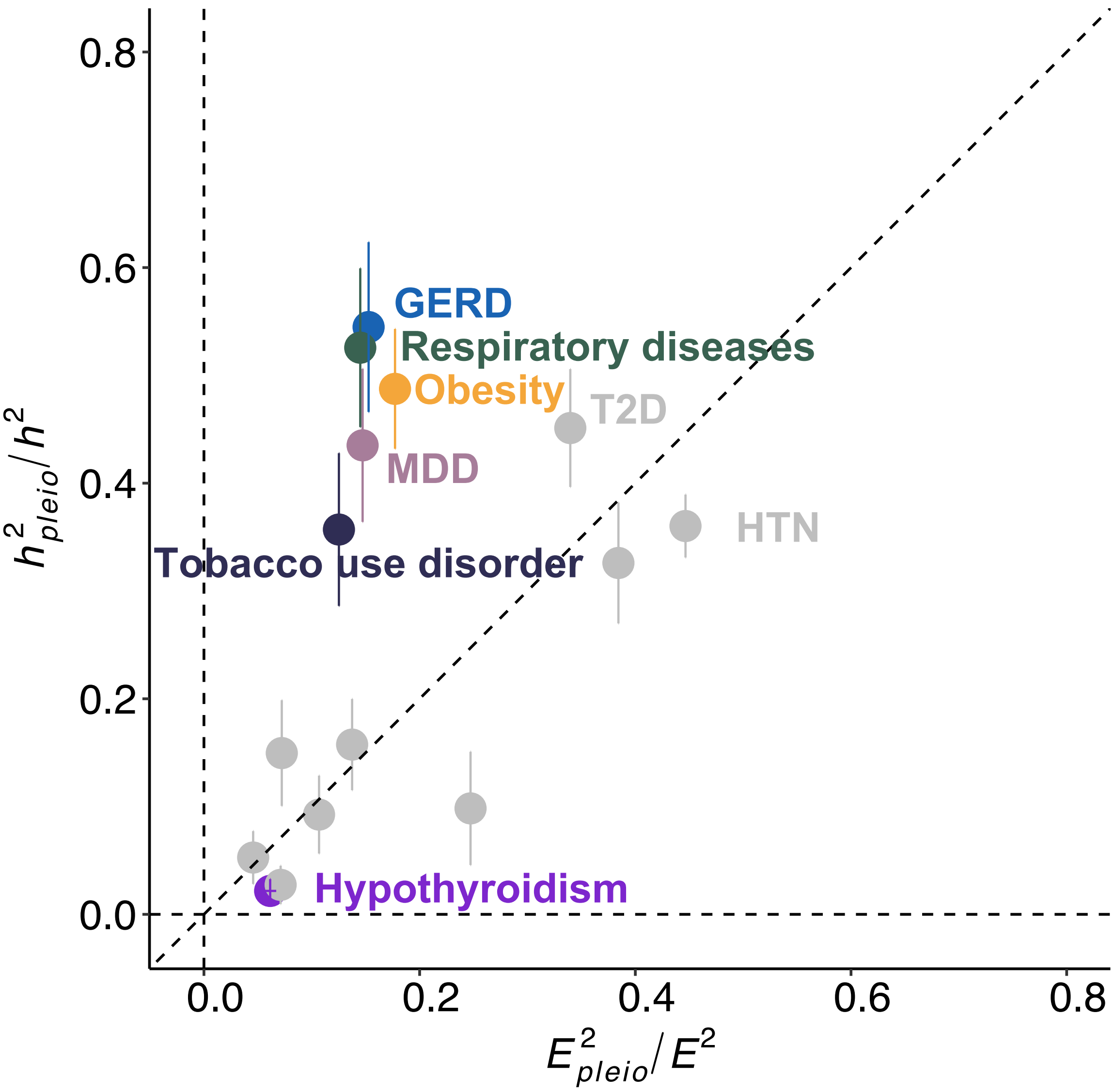


**Supplementary Figure 31. Scatter plot of $h^2{}_{pleio}/h^2$ vs. $E^2{}_{pleio}/E^2$ for the 15 diseases.** We use the same set of auxiliary diseases after pruning in comparing $h^2{}_{pleio}/h^2$ and $E^2{}_{pleio}/E^2$. The colors of dots are the same with **Figure 7**. The estimates of $h^2{}_{pleio}/h^2$ were generally larger than estimates of $E^2{}_{pleio}/E^2$ (ratio of averages = 1.54x). Error bar shows the jackknife standard error. Detailed results are provided in **Supplementary Table 26**. Abbreviation: MDD: depression; T2D: type 2 diabetes; HTN: hypertension; GERD: gastroesophageal reflux disease; AF: Atrial Fibrillation.

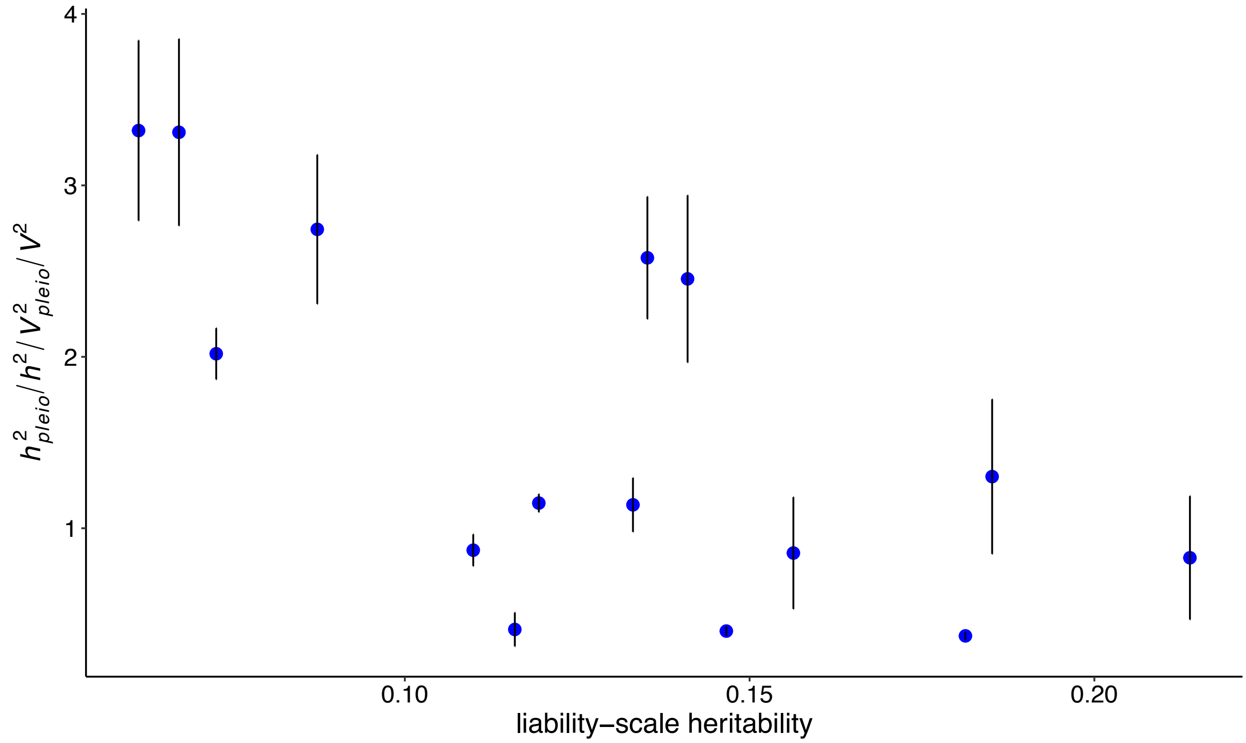


**Supplementary Figure 32. Scatter plot of liability-scale heritability and the ratio of $h^2{}_{pleio}/h^2$ to $V^2{}_{pleio}/V^2$ across the 15 UK Biobank diseases.** We observed a negative correlation (correlation = -0.65, $P$ = 0.008). The p-value of 0.008 is anti-conservative, as it treats the 15 diseases as independent when they are in fact correlated, and it is thus unclear whether the correlation is statistically significant.
